# Interactions between respiratory viruses in US adults and children: a prospective cohort study

**DOI:** 10.64898/2026.10.15.26365672

**Authors:** Katherine L. Hoffman, Ana A. Weil, Erica A. Wetzler, Bethany O. Akinola, Ayisha Beauge, Caroline Andy, Annalyse Adams, Collrane Frivold, Sarah N. Cox, Brenna Ehmen, Tara Hatchie, Erica L. Clark, Mark Drummond, Janet A. Englund, Peter D. Han, Jennifer L. Kuntz, Nathan C. Lo, Christina M. Lockwood, Lea Starita, Richard A. Mularski, Allison L. Naleway, Marco Carone, Helen Y. Chu

**Author notes:** **Corresponding author:** Katherine L. Hoffman. Authors jointly supervised this work (Marco Carone, Helen Y. Chu).

## Abstract

**Background:** The relationship between recent viral infection and subsequent susceptibility to, or protection against, other viruses remains unclear, particularly across age groups and symptom status.

**Methods:** A prospective cohort study enrolled participants aged 6 months to 49 years in Washington and Oregon, USA, 2022–2024. Weekly nasal swabs were tested for SARS-CoV-2, respiratory syncytial virus (RSV), and influenza A/B; additional swabs were collected during symptomatic illness and were also tested for parainfluenza (PIV) 1–4, endemic human coronaviruses (hCOV), adenovirus, human metapneumovirus, human rhinovirus (hRV), and enteroviruses. Proportional hazards regression models were used to characterize associations between incident symptomatic infection (illness) and history of recent illness due to other viruses (within 1–7, 8–14, 15–30, 31–60, 61–90, 91–180 days), adjusted for geographic region, age, sex, daycare/school/work attendance, household size, and number of children in the household. For SARS-CoV-2, influenza, and RSV, these analyses were repeated to study incidence of infection (including asymptomatic cases).

**Findings:** Of 3,560 participants, 2,969 (83%) had ≥1 illness over 4,299 person-years. Adjusted hazard ratios (aHRs) in the short term (≤2 weeks) after an illness were predominantly below 1, indicating that recent viral illness was often associated with lower risk of subsequent illness with other viruses. Across age groups, recent hRV illness was associated with lower risk of hCOV, PIV, and RSV illness (aHRs 0.07–0.27). SARS-CoV-2 illness was associated with a higher short-term risk of adenovirus and PIV illness (aHRs 3.20–3.33). In children aged 6 months–4 years, recent influenza illness was associated with higher short-term risk of SARS-CoV-2 illness. Including asymptomatic infections generally attenuated these associations.

**Interpretation:** Some viral illnesses may alter short-term susceptibility to subsequent viral illnesses. Interactions may differ by age and symptom status.

**Funding:** US Centers for Disease Control and Prevention; Washington State Department of Health.

## INTRODUCTION

Respiratory viruses cause substantial morbidity and mortality worldwide, and multiple viruses co-circulate each season.^1^ A virus–virus interaction occurs when infection with one virus alters infection or replication of another; viral interference denotes the specific case in which the innate immune response to a first virus transiently reduces susceptibility to a second.^2^ Despite their potential impact on the control of epidemics, these interactions remain poorly characterized.^3^

Virus–virus interactions emerged as a scientific hypothesis after live attenuated enterovirus (EV) vaccines, administered to children to prevent poliomyelitis, were found to reduce detection of unrelated respiratory viruses.^4^ Interest was renewed after the 2009 influenza A pandemic appeared to spread later in regions with high human rhinovirus (hRV) circulation^5^, and subsequent surveillance analyses reported negative hRV–influenza A associations at both population and individual levels.^6^ Experimental studies later showed that hRV activates an interferon response in the human airway epithelium sufficient to block replication of influenza^7^ and SARS-CoV-2^8^; influenza A (H3N2) infection may likewise suppress SARS-CoV-2 replication.^9^ Conversely, prior influenza A infection can upregulate ACE2 expression and enhance SARS-CoV-2 infectivity.^10,11^ Together, evidence from cell culture^2,7,12,13^, animal models^10,14^, and ecological observations^15,16^, suggests that a preceding respiratory viral infection may alter susceptibility to other viruses, with effects that may vary by time, pathogen, host, and environment.

Associations consistent with both reduced and increased susceptibility to a second virus have been reported in observational data, typically using retrospective medical records, surveillance data, or registries.^7,17–26^ The reliance of these studies on clinically indicated testing frequently limits interpretation. First, mild and asymptomatic infections, which may alter susceptibility to a subsequent infection, are often missed. Second, testing is not performed systematically during symptom-free periods, making it difficult to characterize virus-negative person-time at risk and to compare the incidence of a second infection after a recent infection with incidence during periods without a recent infection. Additionally, much prior work has relied on codetection of two pathogens in a single specimen, which cannot establish the order or interval between infections; doing so requires repeated longitudinal sampling.

In the present study, we leverage a large, prospective, community-based cohort of adults and children with weekly respiratory sampling and multipathogen testing to assess interactions between nine common respiratory viruses: SARS-CoV-2, influenza, respiratory syncytial virus (RSV), hRV, EV, adenovirus (AdV), human parainfluenza (PIV), human metapneumovirus (hMPV), and endemic human coronavirus (hCOV). We estimate the risk of symptomatic infection (illness) among individuals with and without a different recent illness; for SARS-CoV-2, influenza, and RSV, we additionally study this risk using test results that were collected weekly, regardless of whether an individual reported symptoms.

## METHODS

### Data collection

CASCADIA is a community-based prospective household cohort study characterizing respiratory infections among participants aged 6 months to 49 years from households in Washington State and Oregon, USA.^27^ Between June 2022 and March 2024, enrolled participants completed questionnaires capturing sociodemographic characteristics and vaccination history. Participants also completed weekly symptom surveys and could report new or worsening symptoms at any time.

This study was a secondary analysis of CASCADIA data.^27^ We included all participants who completed an enrollment survey and provided at least one nasal swab specimen during the study period (<u>Supplemental Figure 1</u>).

Irrespective of symptoms, participants self-collected anterior nasal swabs weekly; these were tested for SARS-CoV-2, RSV A/B, and influenza A/B by RT-qPCR. Specimens were additionally tested on a multipathogen (SARS-CoV-2, RSV A/B, influenza A/B/C/H1N1/H3N2, PIV 1–4, EV, AdV, hRV, hCOV, and hMPV) panel if positive for any of these three pathogens, or if the participant reported respiratory symptoms within 72 hours of swabbing. Participants also self-collected an additional swab whenever new or worsening symptoms arose more than 72 hours after their most recent swab; all symptom-triggered specimens were tested on the multipathogen panel. Because a secondary objective of CASCADIA was quantifying SARS-CoV-2 shedding, participants positive for SARS-CoV-2 were additionally asked to swab every 3 days for 21 days. The swab collection protocol, respiratory symptom list, and RT-qPCR methods are detailed in <u>Supplemental Table 1</u> and <u>Supplemental Methods</u>.

### Illness/infection episode definitions

Asymptomatic detections were routinely captured only for SARS-CoV-2, RSV, and influenza. We therefore distinguish between viral illnesses (symptomatic detections only), defined for all viruses, and viral infections (symptomatic or asymptomatic detections), defined only for SARS-CoV-2, RSV, and influenza.

Viral illness episodes were defined using only multipathogen test results associated with symptoms. An illness began on the date of the first positive swab result for a particular virus target and was considered complete after 14 consecutive days without a positive detection from the same target.

For SARS-CoV-2, influenza, and RSV, infection episodes were defined using the same criteria as illness episodes, but incorporating all positive detections from the multipathogen panel, regardless of symptom status. Because participants positive for SARS-CoV-2 were additionally asked to swab every 3 days for 21 days to assess viral shedding, some RSV and influenza detections were found only because of this extra swabbing. If an asymptomatic detection fell within 21 days of the start of a SARS-CoV-2 infection, and was not part of the weekly swab collection irrespective of symptoms, we excluded it from infection episodes so that RSV and influenza detection rates would not be artificially inflated during this period.

Episodes for different viruses, and for different subtypes of the same virus, were allowed to overlap.

### Descriptive characteristics

Descriptive statistics summarized demographic, household, and illness characteristics of the cohort. The weekly proportion of symptomatic positive detections attributable to each virus was computed. The frequency of a secondary illness episode within 1–180 days of a primary illness episode was descriptively reported for all virus pairs.

### Multivariable regression analysis

Associations between recent respiratory viral illness history and subsequent virus-specific illness risk were studied using Cox proportional hazards regression models. Separate models were used for each of nine incident illness outcomes: SARS-CoV-2, RSV, influenza, EV, hRV, AdV, hCOV, PIV, and hMPV.

Calendar time was used as the analysis time scale so that the unspecified baseline hazard function accounts for time-varying infection risk, which is particularly important given respiratory virus seasonality. Methods for left-truncated data were used, with participants entering the risk set at their study enrollment time and contributing person-time until their first illness with the outcome virus, withdrawal from the study, or the end of the study (March 31, 2024). Thus, for each outcome virus, the analysis estimated associations with the hazard of the first detected illness with that virus during study follow-up. Use of a first-event analysis avoided the need for additional assumptions about within-person dependence among recurrent outcome episodes.

For each outcome model, recent illness histories for all eight other respiratory viruses were included as time-varying exposures, categorized according to the time since the most recent illness with that virus: 1–7, 8–14, 15–30, 31–60, 61–90, and 91–180 days, with no illness in the preceding 180 days as the reference category. These definitions permitted recurrent illnesses of the same virus over follow-up, meaning participants could contribute exposure history from multiple temporally distinct illnesses occurring during the study period.

Models included adjustment for prespecified confounders: age category (6 months–1 year, and 2–4, 5–11, 12–17, and 18–49 years), sex at birth, household size, number of children in the household, geographic region, and daycare, school, or work attendance. We report adjusted hazard ratio (aHR) estimates. Each aHR compares the hazard among participants with an illness due to the exposure virus in a given window (e.g., 1–7 days) with that among participants without such an illness in the preceding 180 days, holding the adjustment covariates and recent illness history for other viruses fixed.

We report 95% Wald confidence intervals (CIs) using robust standard error estimates accounting for correlation within households. We report p-values before and after applying the Benjamini-Hochberg (BH) procedure separately within each outcome model to control the false discovery rate at 5%. In the Results, “significant” refers to associations that remain significant after this adjustment.

### Secondary and sensitivity analyses

The primary analysis was repeated with SARS-CoV-2, RSV, and influenza infections (rather than illnesses) defining both the exposures and outcomes for these three viruses. A secondary, exploratory analysis used interaction terms between age category and each time-varying illness exposure to examine potential age-specific differences across pathogens. Age category-specific coefficients and 95% CIs for recent illness were reported. Sparse person-time required collapsing the two youngest age groups into a single category.

In our primary analysis, we considered an illness episode to be distinct if it was followed by 14 days without another symptomatic positive detection. We conducted sensitivity analyses in which we extended this period to 21 and 28 days to account for potential prolonged illness duration. A sensitivity analysis enforcing a delay in study entry until 180 days after enrollment, so that each participant’s full 180-day illness history was observed, was also performed.

## RESULTS

### Cohort characteristics

The cohort comprised 3,560 participants, including 428 children aged 6 months to 4 years at enrollment. The majority were female (58%), non-Hispanic White (70%), from households with a combined income of at least $100,000 (76%), and attending daycare, school, or work (86%). Table 1 shows characteristics overall and by age category.

**Table 1.**
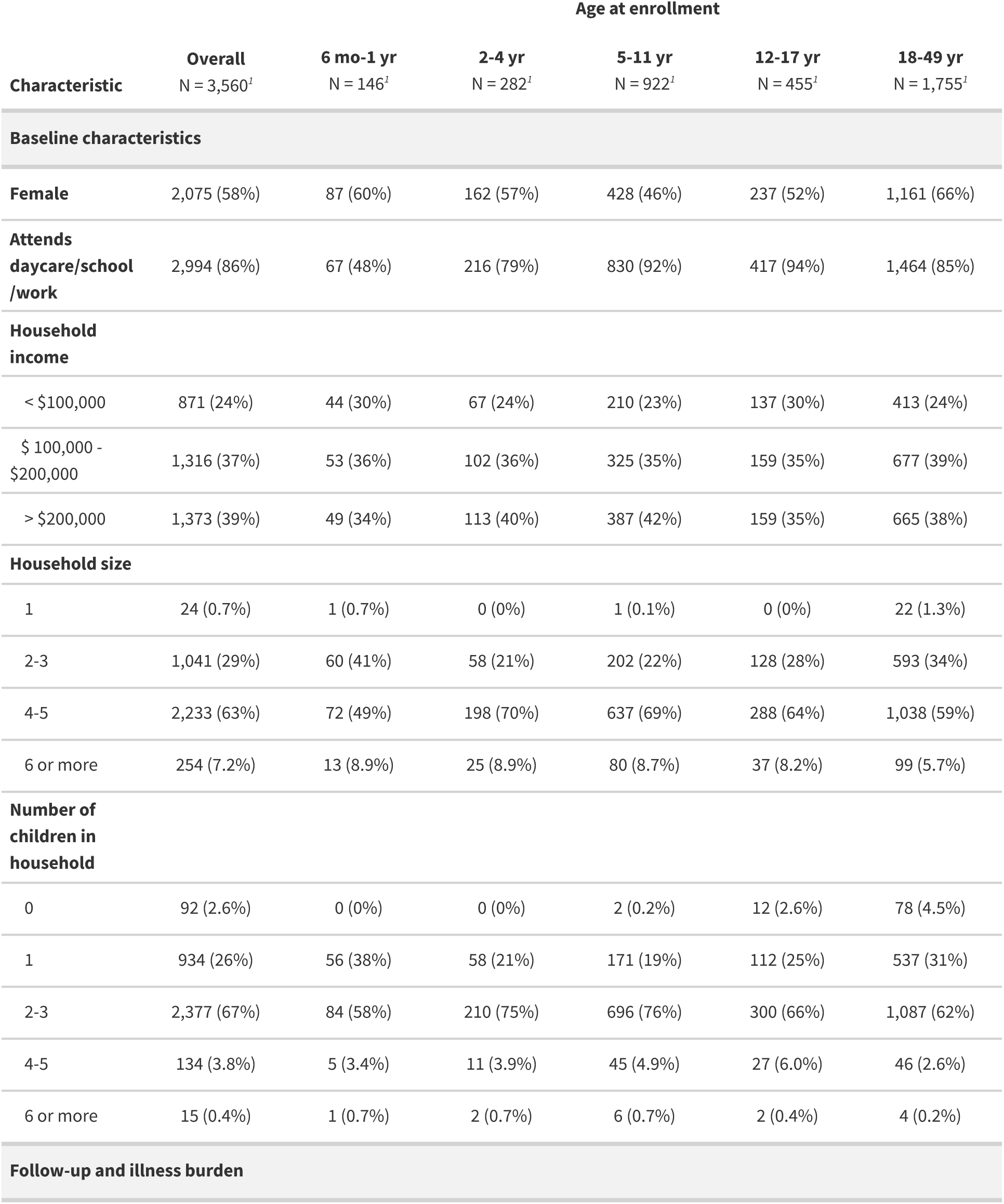
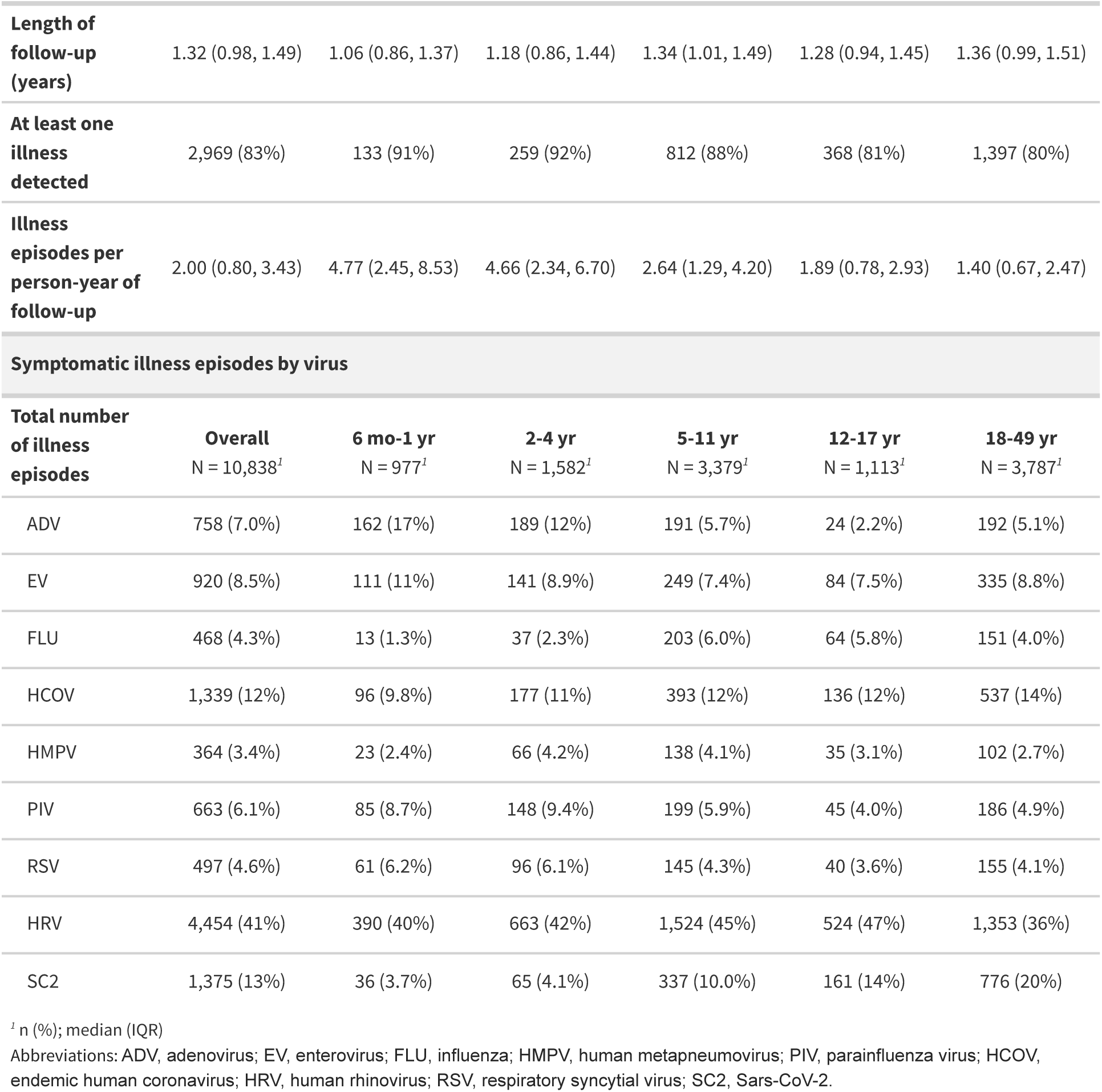
Cohort characteristics and illness burden, overall and stratified by age at enrollment.

Enrollment began June 10, 2022 and concluded December 3, 2023. Active enrollment numbers over time and the number of participants in each age category who submitted at least one swab per week are shown in Figure 1A. The cumulative follow-up for participants in this study was 4,299 person-years, and the median length of follow-up was 1.3 (IQR 1.0–1.5) years. Overall, study protocol adherence was high, with a median weekly swab adherence rate of 85% (IQR 69%–93%).

**Figure 1.**
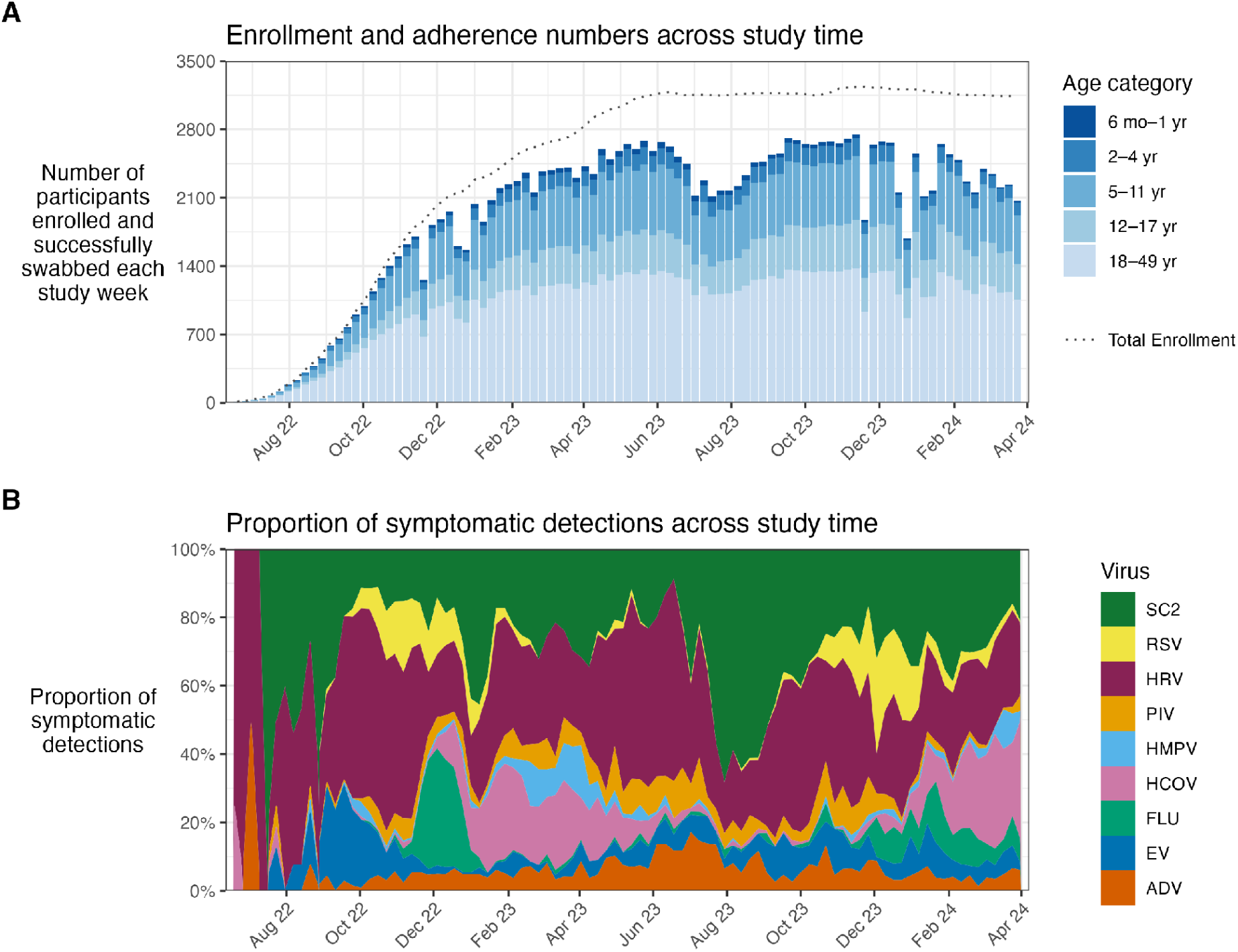
Seasonality of illnesses, swab adherence, and age distribution. Weekly swab collection and respiratory virus positivity across study follow-up, June 2022 to March 2024. **(A)** Number of participants contributing at least one successful swab in each study week, stacked by age category at each swab. The dotted line above each bar indicates the number of participants actively enrolled and under observation that week; the gap between the line and the bars reflects participants enrolled but not successfully swabbed in that week. Bars are counts of participants, not of swabs, i.e., a participant contributing multiple swabs in a week is only counted once. **(B)** Weekly proportion of positive detections for each virus. The x-axis represents study time, and the y-axis shows the percentage of all symptomatic positive detection results are of each virus type. Colors represent viruses: green, SC2; yellow, RSV; maroon, HRV; orange, PIV; light blue, HMPV; pink, HCOV; teal, FLU; blue, EV; dark orange, ADV. ADV, adenovirus; EV, enterovirus; FLU, influenza; HMPV, human metapneumovirus; PIV, parainfluenza virus; HCOV, endemic human coronavirus; HRV, human rhinovirus; RSV, respiratory syncytial virus; SC2, Sars-CoV-2.

The weekly proportion of symptomatic positive detections attributable to each virus is shown in Figure 1B. Overall, 2,969 participants (83%) recorded at least one illness during study follow-up. Of the children 6 months to 4 years old at enrollment, 92% recorded at least one illness, and they contributed a median of 4.7 (IQR 2.4–7.3) illnesses per person-year of follow-up — three times the corresponding rate for adults (1.4; IQR 0.7–2.5). Children often tested positive for many different viruses within overlapping intervals; timelines of three young participants are shown in Figure 2.

**Figure 2.**
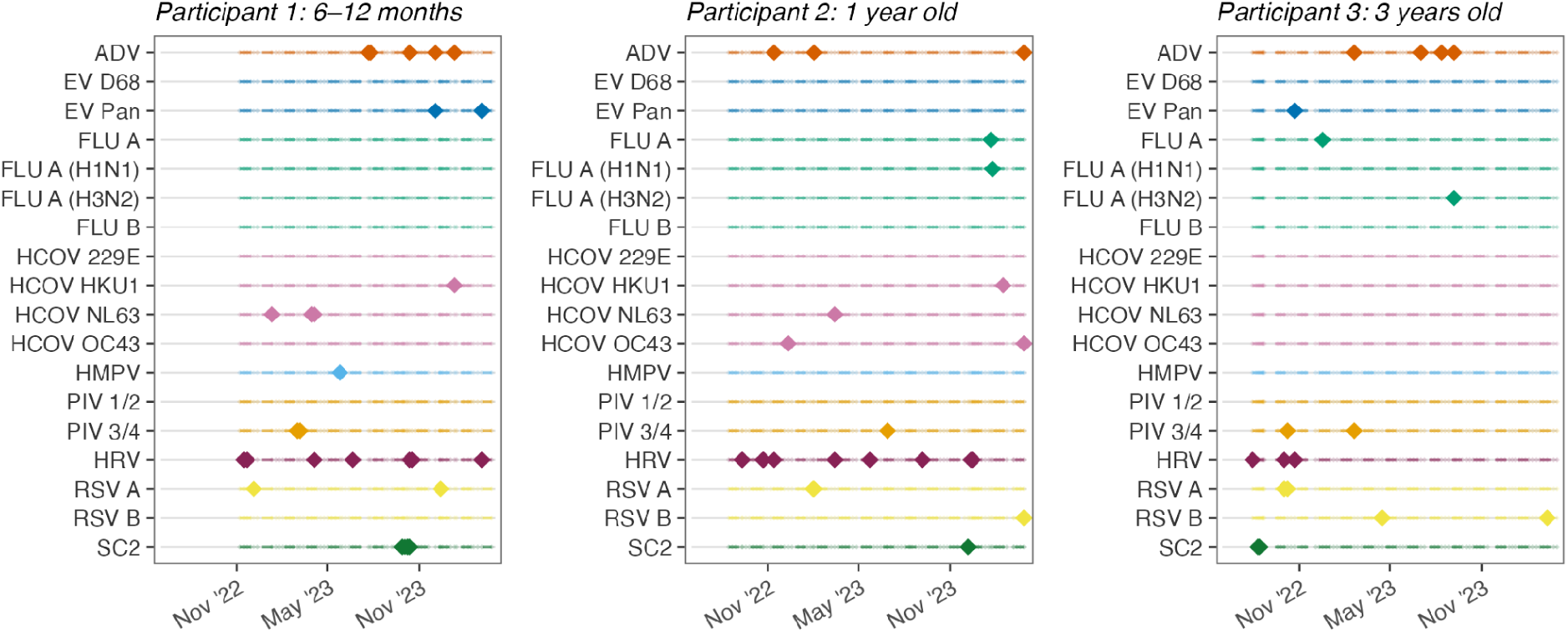
Three pediatric participants’ viral positivity timelines. Timelines for three selected participants under 5 years of age at the time of enrollment. These participants were chosen as examples to illustrate the range of detection frequency and pathogen diversity observed in young children. Each subpanel shows one participant, with age at study enrollment given in the panel header. All panels share a common calendar time axis. Rows correspond to individual viral type or subtype targets on the multipathogen respiratory panel. Large, filled diamonds denote positive detections of a certain target. Marker color identifies the virus group (dark orange, ADV; blue, EV; teal, FLU; pink, HCOV; light blue, HMPV; orange, PIV; maroon, HRV; yellow, RSV; green, SC2), with subtypes distinguished by row. Participants differ in enrollment date and follow-up duration; dashed line spans the participant’s observation period (first to last swab). Closely spaced positive detections on the same row may represent repeated positives within a single illness or infection episode, as detection can persist for multiple weeks after onset. ADV, adenovirus; EV, enterovirus; FLU, influenza; HCOV, human coronavirus; HMPV, human metapneumovirus; PIV, parainfluenza virus; HCOV, endemic human coronavirus; HRV, human rhinovirus; RSV, respiratory syncytial virus; SC2, Sars-CoV-2.

### Secondary illnesses following a primary illness

Figure 3 shows the number and percentage of illnesses detected within 180 days following a primary illness episode. For SARS-CoV-2, RSV, and influenza, these numbers are additionally shown for secondary infections following a primary infection. The most frequently detected secondary illness following every primary illness/infection was hRV, ranging from 30.7% after SARS-CoV-2 illness to 47.4% after AdV illness; hCOV was generally the second most detected secondary illness (8.5%–28.8%). This ordering closely tracks the overall frequency of these pathogens in the cohort — hRV alone accounted for 4,454 (41.1%) of illnesses. These proportions are descriptive and conditional on having experienced a primary episode: the analysis includes no comparator group of participants without a primary episode, and does not incorporate calendar or person-time, so the values cannot be interpreted as the effect of an illness/infection on subsequent episode risk.

**Figure 3.**
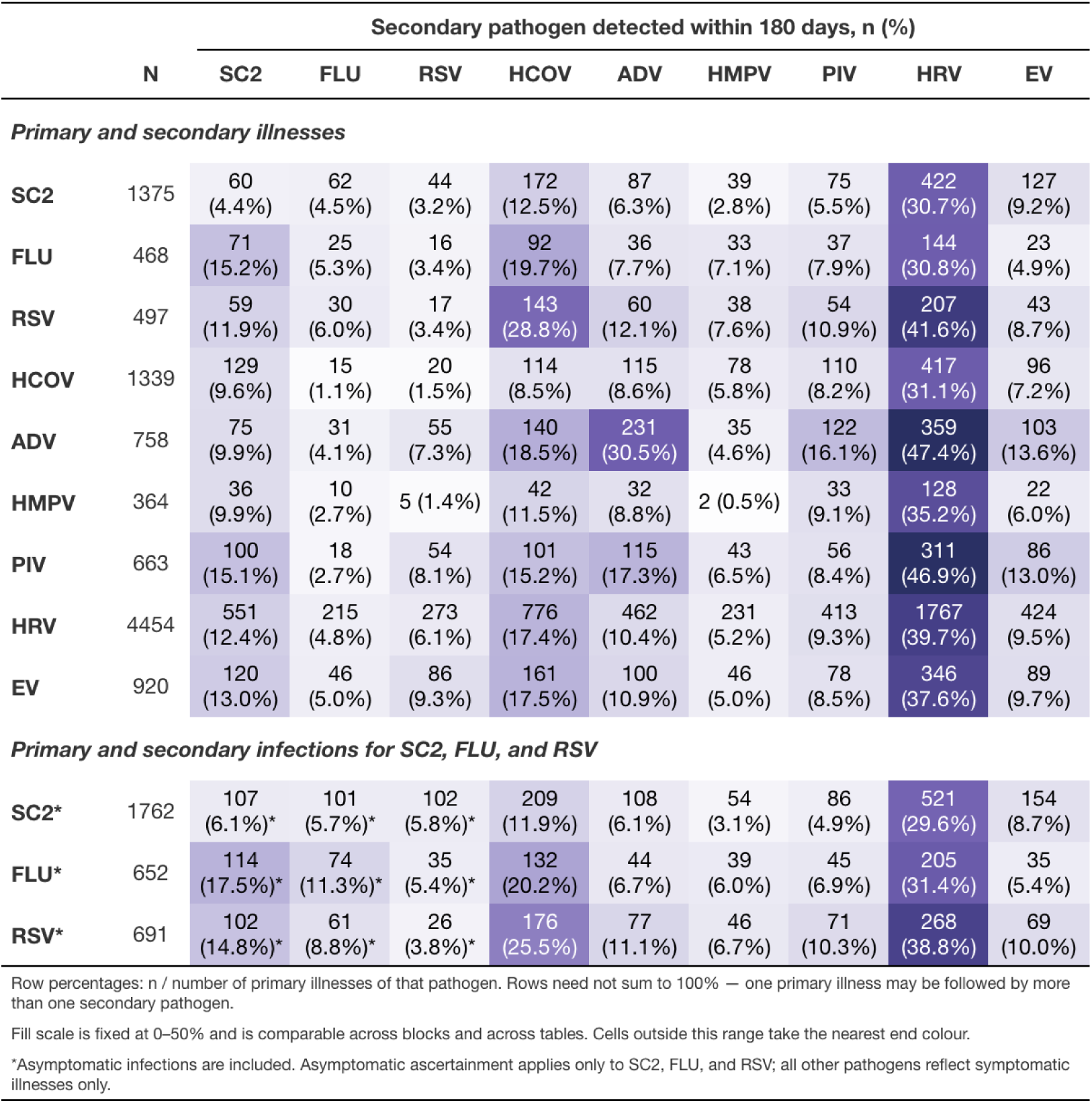
Secondary illness/infection episodes occurring 1-180 days after a primary illness/infection episode, stratified by pathogen type. Cells report the number and percentage of primary episodes of the row pathogen that were followed by at least one subsequent episode in which the column pathogen was detected. Denominators (N) are illness episodes, not participants: a participant may contribute multiple primary episodes and may appear in more than one row. Rows need not sum to 100%, as a single primary episode may be followed by subsequent episodes involving more than one pathogen. Diagonal cells represent re-detection of the same pathogen. The upper block restricts to illnesses; the lower block shows infections, which were systematically ascertained only for SC2, FLU, and RSV (marked with an asterisk). All participants included had a primary episode by definition; no comparator group of uninfected participants or unexposed person-time is represented, and the figure supports no inference about the effect of a primary episode on subsequent risk. Cell shading is proportional to the row percentage and is comparable across both blocks. RSV, respiratory syncytial virus; SC2, SARS-CoV-2; FLU, influenza; ADV, adenovirus; EV, enterovirus; HRV, human rhinovirus; PIV, parainfluenza virus; HCOV, endemic human coronavirus; HMPV, human metapneumovirus.

### Primary analyses

#### Illnesses as exposures and outcomes

The aHRs for each time-varying viral illness exposure, for each time-to-incident illness outcome, are shown in Figure 4A and <u>Supplemental Table 2</u>. In the two shortest windows after a prior illness, aHRs were predominantly below 1; this pattern was largely consistent regardless of the sequence or type of illness. Particularly strong associations were seen for a recent hRV illness and a lower short-term (1–7 and/or 8–14 days) risk of RSV, hCOV, and PIV illnesses (aHRs 0.07–0.27). The exception to this lower short-term risk pattern was that a SARS-CoV-2 illness within the past 1–7 days was associated with a higher risk of AdV and PIV illnesses (aHRs 3.20–3.33). Of note, a higher number of tests were ascertained due to new and worsening symptoms after SARS-CoV-2 illnesses compared to other pathogens (<u>Supplemental Figure 2</u>). For several short-term exposure windows, aHRs were not estimable because no incident illnesses were observed among exposed person-time, leading to complete separation of the data.

**Figure 4:**
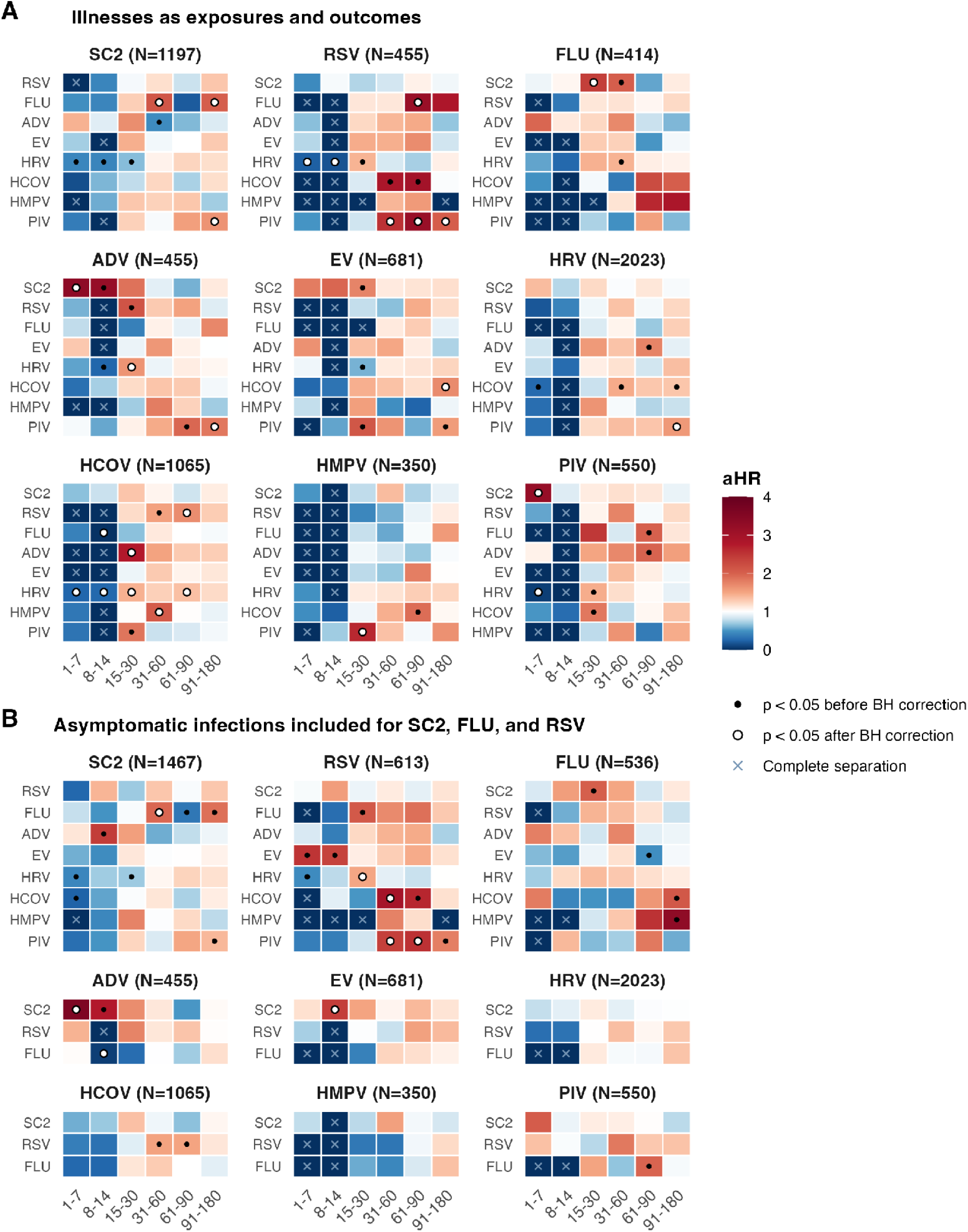
Adjusted HRs for illness/infection with each outcome pathogen following recent illness/infection with a different pathogen, by time since the prior illness/infection. Each panel corresponds to one outcome virus; N denotes the number of incident outcome episodes contributing to the regression model. Within a panel, rows index the prior (exposure) viral illness/infection and columns index the interval since first detection of that prior infection. Cell fill gives the adjusted hazard ratio (aHR) from a single Cox proportional hazards model per outcome pathogen, in which all exposure pathogens and all time windows are entered simultaneously as time-varying indicators; the referent is person-time with no illness/infection within the prior 180 days. Blue indicates aHR < 1, red, aHR > 1, and white, aHR = 1, with a scale from 0 to 4. Black circles, p < 0.05 before but not after Benjamini–Hochberg correction; white circles with black outline, significant after correction; light blue X, no variation (no incident outcomes among exposed person-time; aHR not estimable). Models were adjusted for age category, sex assigned at birth, number of children in the household, household size, daycare/school/work attendance, and geographic region, with robust variance estimation accounting for clustering within households. **(A)** Illness episodes only, for both exposure and outcome. **(B)** Additionally includes asymptomatic infections for SARS-CoV-2, influenza, and RSV; outcome counts therefore differ from panel A only for those three panels, while in the remaining six panels the SARS-CoV-2, influenza, and RSV exposure rows are redefined and the other exposure rows are unchanged apart from shifts induced by the altered adjustment set. SC2, SARS-CoV-2; RSV, respiratory syncytial virus; FLU, influenza; ADV, adenovirus; EV, enterovirus; HRV, human rhinovirus; PIV, parainfluenza virus; HCOV, endemic human coronavirus; HMPV, human metapneumovirus.

For exposures more than two weeks after illness detection, estimates generally attenuated toward the null and, for some virus pairs, were significantly above 1. Higher hazard of RSV illness were observed following influenza illness 61-90 days earlier and PIV illness 31-180 days earlier (aHRs 2.54–3.17). Significant aHRs above 1 were also observed for influenza illness following SARS-CoV-2 illness 15-30 days earlier; hCOV illness following hRV illness 15-30 and 61-90 days earlier, hMPV illness 31-60 days earlier, and RSV illness 61-90 days earlier; hMPV illness following PIV illness 15-30 days earlier; and hRV, AdV, and SARS-CoV-2 illness following PIV illness 91-180 days earlier (aHRs 1.59–1.95).

#### Infections included for SARS-CoV-2, influenza, and RSV

The aHRs for SARS-CoV-2, influenza, and RSV infections as both exposures and outcomes are shown in Figure 4B. For the outcomes of hRV, EV, AdV, hMPV, hCOV, and PIV illness, the short-term risks for those with a recent SARS-CoV-2, influenza, or RSV infection compared with those without were similar to those in the primary analysis, with slightly attenuated estimates overall. An exception was that SARS-CoV-2 infection within 8–14 days was associated with a higher risk of EV illness (aHR 2.42 [1.36–4.28]; the corresponding estimate for SARS-CoV-2 illness was not estimable). When SARS-CoV-2, RSV, and influenza infections were the outcomes, the short-term lower risks seen in the illness analysis were less consistent. Many aHRs for an illness/infection within 1–14 days were attenuated, and recent hRV illness was associated with a higher, rather than lower, risk of RSV infection, although this finding was not significant after BH correction. Associations for exposure windows more than two weeks after the antecedent infection remained qualitatively similar.

### Age-specific analyses

The aHRs specific to each age category are shown in <u>Supplemental Tables 3–6</u>. In this exploratory analysis, age-stratified estimates suggested potential heterogeneity for some virus pairs, although confidence intervals were wide and non-estimable aHRs were more frequent than in the primary analysis. In children 6 months to 4 years, recent (1–7 and 31–60 days) influenza illness/infection was associated with a higher risk of SARS-CoV-2 illness/infection (aHRs 5.50–15.92); a recent (<30 days) AdV illness was associated with a higher risk of SARS-CoV-2 infection and influenza illness/infection (aHRs 4.52–10.35); and a recent SARS-CoV-2 illness/infection was associated with a higher risk of PIV illness (aHRs 3.43–9.54). In 5–11-year-olds, a recent AdV illness was associated with a higher risk of SARS-CoV-2 infection, while in 12–17-year-olds a recent AdV illness was associated with higher risk of influenza illness (and infection), and a recent hMPV illness was associated with higher risk of EV illness (aHRs 3.63–18.26). The estimated short-term inverse associations between recent hRV illness and several other viruses generally held across age strata, although recent hRV illness was associated with a higher risk of EV illness in adults. Also in adults, a recent PIV or SARS-CoV-2 illness was associated with a higher risk of AdV illness, and a recent EV illness was associated with a higher risk of RSV illness (aHRs 4.69–8.67).

### Sensitivity analyses

Sensitivity analyses using an illness definition of 21 and 28 detection-free days yielded results nearly identical to the primary analyses (<u>Supplemental Figure 3</u>). Results enforcing a full 180 days of viral history are displayed in <u>Supplemental Figure 4</u>; estimates were directionally similar.

## DISCUSSION

Using data collected from children and adults with intensive weekly surveillance for respiratory pathogens in a two-year prospective cohort, this study found evidence that recent illness with many common respiratory viruses was associated with a short-term lower risk of illness with several other respiratory viruses. These findings were strongest for the association between hRV illness and the risk of subsequent hCOV, PIV, and RSV illnesses across age groups. An exception was SARS-CoV-2 illness, which was associated with elevated short-term risks of AdV and PIV illness. In children aged 6 months to 4 years, several virus pairs showed higher short-term risk. These findings align with the limited existing clinical^28,29^ and experimental^30^ evidence of enhanced susceptibility to sequential respiratory virus infections in young children and suggest that age-dependent differences in host antiviral responses may contribute to the heterogeneity observed across age groups. Estimates of lower risk were attenuated once asymptomatic infections were included for SARS-CoV-2, influenza, and RSV, which could reflect stronger host responses following symptomatic infection, differences in exposure or outcome ascertainment, or both. Beyond two weeks, risk estimates were less directionally consistent, and, when significant, pointed toward elevated risk. This raises the possibility of shared unmeasured exposure or susceptibility within households or social networks.

Although results suggest there may be some short-term viral interference in the two weeks following a respiratory illness for many common respiratory pathogen pairs, this finding could be partially influenced by behavior and test ascertainment. Non-pharmaceutical interventions (NPIs) following an infection, such as reduced social contact, increased hand hygiene, or masking, may lower the risk of acquiring a second pathogen, and this may also contribute to our findings of lower risk immediately following many infections. However, the strongest inverse associations in our analysis were for a recent hRV illness against secondary illnesses. This finding argues against NPIs as the only mechanism underlying the evidence of a short-term lower risk, because hRV is one of the mildest and most frequently subclinical pathogens in the multipathogen panel. In addition, participants were tested on the multipathogen panel for new and worsening symptoms following SARS-CoV-2 illness at higher frequency than following other viral illnesses. The elevated short-term risks observed after SARS-CoV-2 illness may therefore reflect a combination of differential test ascertainment, driven by this heightened symptom burden and therefore more intensive testing, and a possible SARS-CoV-2–induced increase in susceptibility to secondary pathogens, which may itself contribute to the new or worsening symptoms observed.

Our study’s strengths include high adherence to serial nasal swab collection and testing, allowing us to characterize illnesses with greater temporal resolution than is typically possible in observational studies of virus–virus interactions. The inclusion of young children, who are highly likely to experience frequent viral illnesses but difficult to follow longitudinally, addresses a population that has been understudied prospectively. Examining both very short- and longer-term intervals helps separate transient effects consistent with innate immunity from longer-lasting effects, such as those mediated by adaptive immunity. Because recent illness with every virus category was included in the model simultaneously, each association was estimated conditional on recent illness history. This reduces a source of confounding with measured co-circulating viruses that pairwise analyses cannot address. Use of calendar time as the analysis time scale additionally accounted flexibly for shared temporal variation in infection risk. Together, these findings advance our understanding of the conditions under which virus–virus interactions may occur and highlight the importance of longitudinal studies with frequent pathogen testing, regardless of symptoms, for disentangling the complex interactions among respiratory viruses.

Our study has limitations. First, although we adjusted for measured confounders, residual confounding could operate in either direction. Second, both the time-varying exposure and the outcome were based on detected illnesses/infections rather than true infection-onset or clearance times, so the observed order of detection may not always correspond to the true order of infection acquisition; however, sensitivity analyses using different detection-free windows to define illness episodes yielded nearly identical results. Third, asymptomatic testing results were unavailable for pathogens other than SARS-CoV-2, influenza, and RSV, so true infection rates remain unknown for these other pathogens. Accordingly, for these viruses, our illness outcome reflects symptomatic viral detection rather than acquisition of infection itself, as secondary infections that did not trigger new or worsening symptoms were not detected. Fourth, incomplete symptom reporting, imperfect adherence to specimen collection, and laboratory measurement error may have produced missed or misclassified infections despite intensive surveillance. Fifth, several aHRs — particularly in the shortest risk windows — were non-estimable because of complete separation between the outcome and exposed person-time. Although this pattern is consistent with a protective association, non-estimability may also result from limited exposed person-time, particularly when periods of circulation of the two viruses have limited overlap. Sixth, because outcome models considered only the first detected illness with each virus during follow-up, the analysis may not capture interactions occurring later in the participants with repeated illnesses. Finally, CASCADIA enrolled predominantly households of relatively high socioeconomic status in the US Pacific Northwest and did not include adults aged 50 years or older, limiting generalizability to populations with different demographic characteristics, healthcare access, or exposure patterns.

Nonetheless, our study provides important insights into the potential for viral interference in a large prospective cohort of children and adults undergoing serial respiratory pathogen testing, during a period when patterns of viral circulation and population immunity were undergoing substantial changes after the onset of the COVID-19 pandemic. These findings can inform models of respiratory virus circulation and motivate mechanistic and interventional studies concerning the timing of vaccination, antiviral treatment, and other interventions.

## Human Subjects

This study was reviewed and approved by the Kaiser Permanente Northwest Region Institutional Review Board (See 45 C.F.R. part 46.114; 21 C.F.R. part 56.114). Informed consent was obtained for all adult participants. For pediatric participants, informed consent was obtained from the minor’s parent or legal guardian. We also obtained assent from children aged 7-17. If a participant turned 18 while participating in the study, we then consented these individuals as adults.

## Code availability

All analysis code is publicly available on GitHub: https://github.com/kathoffman/viral-int.

## Data availability

The raw, but deidentified, data (including data dictionaries, study protocols, the statistical analysis plan, and the informed consent form) are available under restricted access due to ethical restrictions and to protect participant confidentiality. Access requests may be submitted to the corresponding author. Access requests will be subject to review and approval by the CDC. The identified data are protected and are not available due to data privacy laws.

