## Supplemental Materials for "Interactions between respiratory viruses in US adults and children: a prospective cohort study"

### Supplemental material

**Supplemental Tables** **2**
Supplemental Table 1: Summary of testing protocol in the CASCADIA study. 2
Supplemental Table 2: Estimates and 95% CIs corresponding to Figure 4. 4
Supplemental Table 3: Age-specific HRs and 95% CI estimates for children 6 months to 4 years old. 7
Supplemental Table 4: Age-specific HRs and 95% CI estimates for children 5–11 years old. 10
Supplemental Table 5: Age-specific HRs and 95% CI estimates for children 12-17 years old. 13
Supplemental Table 6: Age-specific HRs and 95% CI estimates for adults 18–50 years old. 16
Supplemental Figures 19
Supplemental Figure 1: Consort diagram 19
Supplemental Figure 2: Distribution of symptomatic swabs collected after each illness episode. 20
Supplemental Figure 3: Sensitivity analyses with longer case-free windows. 21
Supplemental Figure 4: Sensitivity analyses with a 180 day washout period. 22
Supplemental Methods 23
Laboratory methods 23

### Supplemental Tables

#### Supplemental Table 1: Summary of testing protocol in the CASCADIA study.

| **Timing of nasal swab specimen** | **Testing of specimen** | **Criteria for additional testing** |
|---|---|---|
| Once per week, regardless of symptoms | SARS-CoV-2, RSV A/B, and Influenza A/B on CRI/CRIMP RT-qPCR assay | If nasal specimen was positive or inconclusive for SARS-CoV-2, RSV A, or Influenza A on CRI/CRIMP RT-qPCR assay, then the nasal specimen was sent for multipathogen* Open Array testing<br><br>If a new or worsening respiratory symptoms† was reported at the time of, or within 72 hours following, the nasal specimen collection, then the specimen was sent for multipathogen* Open Array testing |
| Anytime new or worsening symptoms† were reported 72 or more hours since the most recent nasal swab was collected | Multipathogen* Open Array testing | |
| Every 3 days, for 21 days following first positive SARS-CoV-2 test | SARS-CoV-2, RSV, and Influenza on CRI/CRIMP RT-qPCR assay | If nasal specimen was positive or inconclusive for SARS-CoV-2, RSV A, or Influenza A on CRI/CRIMP RT-qPCR assay, then the nasal specimen was |

| | | |
|---|---|---|
| | | sent for multipathogen* Open Array testing<br><br>If a new or worsening respiratory symptoms† was reported at the time of, or within 72 hours following, the nasal specimen collection, then the specimen was sent for multipathogen* Open Array testing |

*Multipathogen testing provided detection results for a broader array of pathogens including influenza A/H3N2, influenza A/H1N1, influenza B, parainfluenza, enterovirus, rhinovirus, adenovirus, human coronaviruses, SARS-CoV-2, RSV A/B, and human metapneumovirus.
† New or worsening symptoms included two or more of the following: fever, chills, cough, shortness of breath, sore throat, muscle or body aches, headache, change in smell or taste, nasal congestion or runny nose, diarrhea, fatigue/being run-down, persistent pain or pressure in chest, or pale, gray, or blue colored skin, lips, or nail beds.

#### Supplemental Table 2: Estimates and 95% CIs corresponding to Figure 4.

Adjusted hazard ratios (aHRs) and 95% confidence intervals are from Cox proportional hazards models with time-varying indicator covariates for each pathogen exposure window (1–7, 8–14, 15–30, 31–60, 61–90, and 91–180 days after first positive swab). Each row represents a distinct exposure pathogen and each outcome pathogen is shown in a separate block. The left panel includes symptomatic infections only; the right panel additionally includes asymptomatic infections for SC2, FLU, and RSV. Cell shading reflects the magnitude of the aHR: navy indicates values below 1, white indicates 1, and red indicates values above 1 (clamped at 4). Bold indicates $p<0.05$; em dashes denote cells with complete separation of the exposure and outcome data, where the aHR was not estimable. Abbreviations: SC2, SARS-CoV-2; FLU, influenza; RSV, respiratory syncytial virus; ADV, adenovirus; EV, enterovirus; RV, rhinovirus; HCOV, seasonal coronavirus; HMPV, human metapneumovirus; HPIV, human parainfluenza virus.

#### Overall hazard ratios (1 of 2)

Hazard of symptomatic infection with a second pathogen

| Exposure | Symptomatic infections only | | | | | | Asymptomatic infections included (SC2, FLU, RSV) | | | | | |
|---|---|---|---|---|---|---|---|---|---|---|---|---|
| | 1–7 | 8–14 | 15–30 | 31–60 | 61–90 | 91–180 | 1–7 | 8–14 | 15–30 | 31–60 | 61–90 | 91–180 |
| **Outcome: SC2** | | | | | | | | | | | | |
| RSV | — | 0.42 (0.06, 2.93) | 0.96 (0.42, 2.18) | 1.17 (0.68, 2.03) | 0.82 (0.38, 1.74) | 1.32 (0.82, 2.12) | 0.26 (0.04, 1.86) | 1.41 (0.63, 3.16) | 0.70 (0.33, 1.46) | 1.32 (0.84, 2.06) | 0.97 (0.58, 1.65) | 1.31 (0.86, 2.00) |
| FLU | 0.47 (0.07, 3.33) | 0.43 (0.07, 2.50) | 1.27 (0.51, 3.16) | **2.12 (1.30, 3.48)** | 0.20 (0.04, 1.08) | **2.03 (1.26, 3.27)** | 0.80 (0.26, 2.46) | 0.50 (0.13, 1.95) | 0.98 (0.45, 2.10) | **1.95 (1.30, 2.92)** | **0.35 (0.13, 0.95)** | **1.89 (1.23, 2.90)** |
| ADV | 1.43 (0.45, 4.52) | 0.88 (0.22, 3.50) | 1.67 (0.93, 3.02) | **0.46 (0.23, 0.90)** | 0.63 (0.33, 1.20) | 0.87 (0.55, 1.36) | 1.14 (0.36, 3.58) | **2.42 (1.21, 4.82)** | 1.54 (0.90, 2.64) | 0.59 (0.33, 1.03) | 0.79 (0.46, 1.34) | 0.94 (0.61, 1.43) |
| EV | 0.71 (0.16, 3.11) | — | 1.45 (0.80, 2.62) | 0.95 (0.53, 1.72) | 1.00 (0.53, 1.88) | 1.29 (0.94, 1.78) | 0.60 (0.14, 2.55) | 0.46 (0.11, 1.89) | 1.13 (0.62, 2.06) | 0.99 (0.58, 1.69) | 1.00 (0.58, 1.73) | 1.07 (0.79, 1.46) |
| RV | **0.43 (0.22, 0.86)** | **0.42 (0.21, 0.85)** | **0.64 (0.45, 0.91)** | 1.07 (0.81, 1.42) | 1.24 (0.94, 1.62) | 1.18 (0.99, 1.41) | **0.43 (0.23, 0.80)** | 0.70 (0.45, 1.11) | **0.70 (0.51, 0.94)** | 1.05 (0.81, 1.36) | 1.24 (0.98, 1.57) | 1.17 (1.00, 1.36) |
| HCOV | 0.15 (0.02, 1.05) | 0.62 (0.24, 1.61) | 0.79 (0.42, 1.48) | 1.22 (0.80, 1.87) | 0.73 (0.37, 1.44) | 1.18 (0.87, 1.60) | **0.24 (0.06, 0.95)** | 0.56 (0.23, 1.33) | 0.98 (0.60, 1.62) | 1.14 (0.75, 1.74) | 0.73 (0.40, 1.33) | 1.10 (0.84, 1.44) |
| HMPV | — | 0.59 (0.10, 3.55) | 1.31 (0.36, 4.80) | 1.25 (0.59, 2.66) | 1.31 (0.52, 3.34) | 0.71 (0.38, 1.32) | — | 0.44 (0.07, 2.89) | 1.75 (0.71, 4.34) | 0.99 (0.46, 2.10) | 1.20 (0.52, 2.81) | 0.78 (0.46, 1.32) |
| HPIV | 0.38 (0.05, 2.61) | — | 1.25 (0.42, 3.72) | 0.97 (0.54, 1.74) | 1.55 (0.90, 2.65) | **1.63 (1.16, 2.30)** | 0.30 (0.04, 2.09) | 0.50 (0.13, 1.99) | 1.23 (0.50, 3.04) | 0.86 (0.51, 1.47) | 1.50 (0.95, 2.36) | **1.39 (1.00, 1.94)** |
| **Outcome: RSV** | | | | | | | | | | | | |
| SC2 | 0.45 (0.06, 3.25) | 0.97 (0.24, 3.95) | 0.75 (0.28, 1.97) | 0.84 (0.37, 1.87) | 1.08 (0.54, 2.14) | 0.95 (0.47, 1.92) | 1.47 (0.65, 3.30) | 1.57 (0.70, 3.56) | 1.06 (0.55, 2.04) | 1.12 (0.64, 1.96) | 0.90 (0.52, 1.58) | 1.14 (0.74, 1.78) |
| FLU | — | — | 1.14 (0.42, 3.10) | 1.13 (0.40, 3.21) | **3.15 (1.23, 8.06)** | 2.83 (0.86, 9.34) | — | 0.30 (0.04, 2.01) | **2.08 (1.11, 3.89)** | 1.68 (0.82, 3.42) | 1.86 (0.75, 4.59) | 1.26 (0.37, 4.30) |
| ADV | 0.65 (0.09, 4.68) | — | 1.21 (0.54, 2.74) | 1.40 (0.73, 2.69) | 1.47 (0.69, 3.14) | 0.68 (0.23, 2.00) | 0.86 (0.22, 3.33) | 0.39 (0.05, 2.80) | 1.20 (0.55, 2.60) | 1.42 (0.80, 2.53) | 1.48 (0.69, 3.18) | 0.78 (0.36, 1.69) |
| EV | 0.76 (0.11, 5.05) | — | 1.46 (0.85, 2.50) | 1.45 (0.85, 2.46) | 1.68 (0.95, 3.00) | 0.83 (0.41, 1.70) | **2.50 (1.08, 5.80)** | **2.32 (1.04, 5.16)** | 1.20 (0.68, 2.12) | 1.27 (0.78, 2.06) | 1.47 (0.88, 2.45) | 1.16 (0.67, 2.01) |
| RV | **0.22 (0.07, 0.66)** | **0.17 (0.04, 0.68)** | **1.42 (1.02, 1.97)** | 0.85 (0.61, 1.20) | 0.72 (0.46, 1.14) | 1.09 (0.76, 1.55) | **0.45 (0.24, 0.87)** | 0.78 (0.46, 1.33) | **1.47 (1.09, 1.98)** | 0.91 (0.68, 1.23) | 0.81 (0.54, 1.22) | 1.01 (0.75, 1.37) |
| HCOV | — | — | 0.86 (0.27, 2.73) | **2.86 (1.09, 7.53)** | **3.00 (1.06, 8.50)** | 0.98 (0.39, 2.50) | — | 0.89 (0.22, 3.72) | 0.50 (0.14, 1.77) | **2.92 (1.27, 6.69)** | **2.62 (1.02, 6.74)** | 1.34 (0.65, 2.77) |
| HMPV | — | — | — | 1.40 (0.40, 4.88) | 1.58 (0.40, 6.28) | — | — | — | — | 1.74 (0.65, 4.65) | 1.15 (0.30, 4.43) | — |
| HPIV | 0.50 (0.07, 3.64) | — | 0.83 (0.30, 2.26) | **2.54 (1.50, 4.31)** | **3.17 (1.78, 5.67)** | **2.03 (1.15, 3.60)** | 0.38 (0.05, 2.74) | 0.38 (0.05, 2.77) | 0.83 (0.35, 1.95) | **2.36 (1.51, 3.70)** | **2.53 (1.44, 4.44)** | **1.75 (1.07, 2.87)** |
| **Outcome: FLU** | | | | | | | | | | | | |
| SC2 | 0.94 (0.23, 3.81) | 1.05 (0.26, 4.25) | **2.40 (1.25, 4.62)** | **2.03 (1.08, 3.80)** | 0.56 (0.13, 2.53) | 1.10 (0.60, 2.03) | 1.15 (0.43, 3.09) | **2.22 (1.05, 4.66)** | **1.89 (1.00, 3.55)** | 1.49 (0.86, 2.58) | 0.84 (0.34, 2.06) | 1.12 (0.73, 1.74) |
| RSV | — | 0.35 (0.06, 2.18) | 1.13 (0.59, 2.20) | 1.15 (0.64, 2.07) | 1.05 (0.33, 3.31) | 1.20 (0.39, 3.75) | — | 0.58 (0.14, 2.45) | 1.42 (0.87, 2.31) | 1.36 (0.86, 2.13) | 1.15 (0.56, 2.36) | 0.80 (0.31, 2.05) |
| ADV | 1.96 (0.51, 7.47) | 1.12 (0.17, 7.36) | 1.22 (0.51, 2.93) | 1.71 (0.83, 3.52) | 0.81 (0.26, 2.54) | 0.62 (0.29, 1.32) | 1.76 (0.58, 5.34) | 1.32 (0.44, 3.95) | 0.84 (0.32, 2.19) | 1.74 (0.87, 3.48) | 0.89 (0.35, 2.29) | 0.94 (0.51, 1.70) |
| EV | — | — | 1.28 (0.57, 2.91) | 1.26 (0.74, 2.14) | 0.51 (0.25, 1.02) | 0.95 (0.52, 1.76) | 0.63 (0.08, 4.74) | 0.50 (0.07, 3.62) | 0.95 (0.41, 2.19) | 1.17 (0.72, 1.90) | **0.47 (0.25, 0.91)** | 0.96 (0.56, 1.66) |
| RV | 0.56 (0.21, 1.46) | 0.27 (0.06, 1.10) | 1.43 (0.95, 2.17) | **1.42 (1.05, 1.93)** | 1.09 (0.74, 1.60) | 1.07 (0.78, 1.46) | 0.71 (0.34, 1.48) | 1.29 (0.72, 2.31) | 1.34 (0.92, 1.94) | 1.30 (0.98, 1.71) | 1.17 (0.83, 1.65) | 1.10 (0.82, 1.47) |
| HCOV | 0.49 (0.07, 3.31) | — | 0.96 (0.30, 3.06) | 0.36 (0.07, 1.74) | 2.25 (0.68, 7.46) | 2.02 (0.85, 4.80) | 1.80 (0.63, 5.10) | 0.42 (0.06, 3.02) | 0.49 (0.13, 1.94) | 0.45 (0.12, 1.73) | 1.59 (0.50, 5.09) | **2.09 (1.04, 4.21)** |
| HMPV | — | — | — | 1.15 (0.30, 4.42) | 2.64 (0.73, 9.51) | 2.86 (0.93, 8.76) | — | — | 0.84 (0.12, 5.78) | 1.29 (0.41, 4.02) | 2.54 (0.89, 7.28) | **3.31 (1.33, 8.25)** |
| HPIV | — | — | 0.73 (0.23, 2.31) | 0.49 (0.16, 1.52) | 1.53 (0.66, 3.54) | 0.63 (0.22, 1.83) | — | 1.37 (0.44, 4.29) | 0.64 (0.21, 1.94) | 0.71 (0.32, 1.60) | 1.82 (0.88, 3.76) | 0.58 (0.26, 1.33) |
| **Outcome: ADV** | | | | | | | | | | | | |
| SC2 | **3.33 (1.45, 7.64)** | **3.20 (1.27, 8.07)** | 1.87 (0.96, 3.66) | 0.90 (0.41, 1.95) | 0.60 (0.26, 1.40) | 1.12 (0.71, 1.76) | **3.62 (1.93, 6.78)** | **2.75 (1.19, 6.40)** | 1.69 (0.85, 3.34) | 1.13 (0.62, 2.05) | 0.51 (0.24, 1.11) | 1.03 (0.66, 1.59) |
| RSV | 0.61 (0.09, 4.34) | — | **2.15 (1.10, 4.21)** | 1.29 (0.65, 2.59) | 1.53 (0.73, 3.18) | 0.88 (0.42, 1.84) | 1.42 (0.44, 4.52) | — | 1.76 (0.90, 3.45) | 1.27 (0.67, 2.41) | 1.34 (0.67, 2.66) | 1.01 (0.53, 1.96) |
| FLU | 0.79 (0.11, 5.46) | — | 0.40 (0.05, 2.91) | 1.08 (0.42, 2.77) | 0.91 (0.24, 3.47) | 1.72 (0.92, 3.21) | 1.00 (0.23, 4.32) | **0.07 (0.02, 0.28)** | 0.29 (0.04, 2.31) | 0.99 (0.41, 2.39) | 0.70 (0.18, 2.72) | 1.16 (0.61, 2.23) |
| EV | 1.30 (0.31, 5.49) | — | 0.92 (0.42, 2.01) | 1.60 (0.90, 2.86) | 1.08 (0.58, 2.00) | 1.00 (0.60, 1.65) | 1.30 (0.31, 5.46) | — | 0.93 (0.42, 2.02) | 1.60 (0.89, 2.85) | 1.09 (0.58, 2.04) | 0.99 (0.60, 1.64) |
| RV | 0.55 (0.26, 1.20) | **0.25 (0.08, 0.81)** | **1.63 (1.15, 2.30)** | 0.93 (0.63, 1.37) | 1.10 (0.75, 1.60) | 1.05 (0.77, 1.43) | 0.54 (0.25, 1.18) | **0.26 (0.08, 0.82)** | **1.65 (1.16, 2.33)** | 0.93 (0.63, 1.37) | 1.11 (0.76, 1.61) | 1.06 (0.77, 1.44) |
| HCOV | 0.31 (0.04, 2.18) | 0.72 (0.18, 2.92) | 1.20 (0.59, 2.44) | 1.32 (0.76, 2.32) | 1.29 (0.69, 2.42) | 0.96 (0.60, 1.53) | 0.31 (0.04, 2.19) | 0.72 (0.18, 2.93) | 1.19 (0.58, 2.42) | 1.31 (0.74, 2.29) | 1.28 (0.68, 2.39) | 0.95 (0.60, 1.53) |
| HMPV | — | — | 0.71 (0.17, 2.87) | 1.79 (0.88, 3.67) | 1.25 (0.54, 2.87) | 0.71 (0.33, 1.53) | — | — | 0.72 (0.18, 2.95) | 1.82 (0.89, 3.72) | 1.25 (0.54, 2.87) | 0.71 (0.33, 1.54) |
| HPIV | 0.97 (0.24, 3.94) | 0.57 (0.08, 4.09) | 1.08 (0.45, 2.55) | 1.47 (0.74, 2.91) | **1.95 (1.07, 3.58)** | **1.84 (1.18, 2.85)** | 0.97 (0.24, 3.95) | 0.58 (0.08, 4.12) | 1.09 (0.46, 2.59) | 1.46 (0.74, 2.90) | **1.98 (1.09, 3.60)** | **1.84 (1.18, 2.87)** |

Columns within each block give exposure windows in days. Cell shading reflects the hazard ratio: navy below 1, white at 1, red above 1, clamped at 4. Bold indicates $p < 0.05$ ; em dashes denote cells where the hazard ratio was not estimable. Blank cells were not fitted in that model.

#### Overall hazard ratios (2 of 2)

Hazard of symptomatic infection with a second pathogen

| Exposure | Symptomatic infections only: 1–7 | 8–14 | 15–30 | 31–60 | 61–90 | 91–180 | Asymptomatic infections included (SC2, FLU, RSV): 1–7 | 8–14 | 15–30 | 31–60 | 61–90 | 91–180 |
|---|---|---|---|---|---|---|---|---|---|---|---|---|
| **Outcome: EV** | | | | | | | | | | | | |
| SC2 | 1.80 (0.78, 4.19) | 1.96 (0.93, 4.13) | **1.74 (1.05, 2.88)** | 1.11 (0.69, 1.79) | 1.15 (0.66, 2.00) | 1.18 (0.82, 1.68) | 1.17 (0.46, 2.95) | **2.42 (1.36, 4.28)** | 1.51 (0.94, 2.42) | 1.06 (0.69, 1.62) | 1.39 (0.90, 2.14) | 1.15 (0.84, 1.58) |
| RSV | — | — | 0.80 (0.26, 2.42) | 0.65 (0.24, 1.78) | 1.46 (0.71, 3.02) | 1.19 (0.68, 2.09) | 0.72 (0.18, 2.85) | — | 0.98 (0.41, 2.37) | 0.81 (0.38, 1.73) | 1.52 (0.84, 2.76) | 1.37 (0.82, 2.30) |
| FLU | — | — | — | 1.12 (0.47, 2.64) | 1.28 (0.53, 3.12) | 0.90 (0.44, 1.87) | — | — | 0.40 (0.06, 2.89) | 1.23 (0.58, 2.57) | 1.17 (0.52, 2.62) | 1.07 (0.59, 1.95) |
| ADV | 1.64 (0.60, 4.51) | — | 1.35 (0.70, 2.60) | 1.49 (0.86, 2.56) | 1.10 (0.57, 2.14) | 0.74 (0.41, 1.32) | 1.60 (0.58, 4.39) | — | 1.33 (0.68, 2.59) | 1.47 (0.86, 2.54) | 1.08 (0.56, 2.09) | 0.73 (0.41, 1.31) |
| RV | 0.94 (0.57, 1.55) | — | **0.61 (0.38, 0.95)** | 1.11 (0.80, 1.55) | 1.15 (0.82, 1.60) | 1.23 (0.98, 1.55) | 0.94 (0.57, 1.54) | — | **0.61 (0.39, 0.96)** | 1.11 (0.80, 1.54) | 1.14 (0.82, 1.60) | 1.22 (0.97, 1.54) |
| HCOV | 0.27 (0.04, 1.92) | 0.33 (0.05, 2.31) | 1.44 (0.77, 2.72) | 1.34 (0.78, 2.30) | 1.18 (0.64, 2.16) | **1.69 (1.17, 2.44)** | 0.26 (0.04, 1.92) | 0.33 (0.05, 2.30) | 1.44 (0.76, 2.72) | 1.32 (0.77, 2.26) | 1.17 (0.64, 2.13) | **1.68 (1.16, 2.43)** |
| HMPV | 0.79 (0.11, 5.71) | — | 1.54 (0.56, 4.24) | 0.47 (0.12, 1.94) | 0.25 (0.04, 1.65) | 0.94 (0.50, 1.76) | 0.77 (0.11, 5.53) | — | 1.53 (0.56, 4.23) | 0.47 (0.12, 1.93) | 0.25 (0.04, 1.67) | 0.94 (0.50, 1.77) |
| HPIV | — | 0.56 (0.08, 3.86) | **2.12 (1.15, 3.92)** | 1.63 (0.92, 2.86) | 0.59 (0.27, 1.29) | **1.57 (1.09, 2.26)** | — | 0.56 (0.08, 3.94) | **2.13 (1.15, 3.95)** | 1.62 (0.92, 2.86) | 0.59 (0.27, 1.28) | **1.56 (1.08, 2.24)** |
| **Outcome: RV** | | | | | | | | | | | | |
| SC2 | 1.37 (0.81, 2.32) | 0.76 (0.38, 1.52) | 1.20 (0.84, 1.73) | 0.98 (0.72, 1.35) | 1.13 (0.81, 1.58) | 0.97 (0.74, 1.26) | 0.80 (0.44, 1.45) | 0.90 (0.52, 1.55) | 1.11 (0.80, 1.55) | 0.90 (0.67, 1.21) | 0.98 (0.73, 1.33) | 0.99 (0.79, 1.24) |
| RSV | 0.18 (0.03, 1.11) | 0.41 (0.10, 1.70) | 0.94 (0.55, 1.63) | 1.34 (0.85, 2.10) | 0.96 (0.52, 1.77) | 1.22 (0.82, 1.80) | 0.30 (0.08, 1.08) | 0.33 (0.08, 1.33) | 1.00 (0.61, 1.63) | 1.36 (0.87, 2.02) | 1.04 (0.60, 1.81) | 1.30 (0.92, 1.84) |
| FLU | — | — | 0.87 (0.36, 2.09) | 1.06 (0.65, 1.73) | 0.69 (0.31, 1.54) | 1.27 (0.85, 1.89) | — | — | 1.10 (0.56, 2.18) | 0.96 (0.59, 1.57) | 0.95 (0.54, 1.67) | 1.35 (0.95, 1.92) |
| ADV | 0.83 (0.27, 2.59) | — | 1.58 (0.96, 2.61) | 1.33 (0.79, 2.25) | **1.75 (1.09, 2.80)** | 0.90 (0.52, 1.57) | 0.82 (0.26, 2.56) | — | 1.58 (0.96, 2.60) | 1.34 (0.80, 2.27) | **1.76 (1.10, 2.82)** | 0.90 (0.52, 1.56) |
| EV | 0.79 (0.25, 2.49) | 0.29 (0.04, 2.05) | 1.09 (0.57, 2.09) | 1.16 (0.66, 2.05) | 0.83 (0.40, 1.75) | 1.36 (0.86, 2.14) | 0.80 (0.25, 2.53) | 0.29 (0.04, 2.08) | 1.10 (0.58, 2.11) | 1.18 (0.67, 2.06) | 0.84 (0.40, 1.77) | 1.34 (0.85, 2.11) |
| HCOV | **0.28 (0.09, 0.85)** | — | 0.81 (0.51, 1.29) | **1.46 (1.08, 1.98)** | 1.31 (0.89, 1.92) | **1.32 (1.01, 1.73)** | **0.28 (0.09, 0.86)** | — | 0.81 (0.51, 1.29) | **1.46 (1.08, 1.97)** | 1.31 (0.89, 1.92) | **1.32 (1.01, 1.73)** |
| HMPV | 0.30 (0.04, 2.11) | — | 1.63 (0.95, 2.80) | 0.97 (0.54, 1.74) | 0.75 (0.37, 1.51) | 0.81 (0.48, 1.36) | 0.30 (0.04, 2.11) | — | 1.62 (0.94, 2.79) | 0.97 (0.54, 1.74) | 0.75 (0.37, 1.51) | 0.81 (0.48, 1.36) |
| HPIV | 0.31 (0.08, 1.24) | — | 1.20 (0.71, 2.03) | 1.37 (0.89, 2.10) | 1.32 (0.85, 2.05) | **1.59 (1.14, 2.21)** | 0.31 (0.08, 1.24) | — | 1.19 (0.70, 2.02) | 1.37 (0.89, 2.10) | 1.33 (0.86, 2.05) | **1.59 (1.14, 2.21)** |
| **Outcome: HCOV** | | | | | | | | | | | | |
| SC2 | 0.66 (0.25, 1.74) | 0.81 (0.29, 2.28) | 1.32 (0.83, 2.10) | 1.06 (0.73, 1.54) | 0.82 (0.50, 1.36) | 1.10 (0.82, 1.49) | 0.59 (0.25, 1.42) | 0.69 (0.27, 1.75) | 1.35 (0.91, 2.00) | 0.94 (0.66, 1.33) | 0.66 (0.41, 1.05) | 1.05 (0.80, 1.38) |
| RSV | — | — | 1.19 (0.64, 2.23) | **1.53 (1.05, 2.23)** | **1.67 (1.19, 2.36)** | 1.28 (0.92, 1.78) | 0.39 (0.05, 2.74) | 0.32 (0.05, 2.17) | 0.89 (0.46, 1.69) | **1.51 (1.07, 2.13)** | **1.50 (1.08, 2.09)** | 1.13 (0.83, 1.54) |
| FLU | 0.40 (0.05, 2.97) | **0.01 (0.00, 0.06)** | 0.66 (0.26, 1.66) | 1.12 (0.75, 1.68) | 1.00 (0.64, 1.57) | 0.97 (0.61, 1.53) | 0.53 (0.13, 2.15) | 0.26 (0.04, 1.68) | 1.05 (0.58, 1.90) | 1.32 (0.94, 1.84) | 1.00 (0.66, 1.51) | 0.90 (0.58, 1.38) |
| ADV | — | — | **2.75 (1.80, 4.21)** | 1.52 (0.91, 2.52) | 1.29 (0.79, 2.11) | 1.27 (0.83, 1.96) | — | — | **2.80 (1.82, 4.29)** | 1.53 (0.93, 2.54) | 1.31 (0.80, 2.16) | 1.28 (0.83, 1.97) |
| EV | — | — | 0.93 (0.46, 1.87) | 1.29 (0.81, 2.08) | 1.07 (0.64, 1.77) | 1.13 (0.85, 1.52) | — | — | 0.93 (0.46, 1.87) | 1.28 (0.80, 2.06) | 1.05 (0.63, 1.74) | 1.13 (0.84, 1.51) |
| RV | **0.25 (0.10, 0.60)** | **0.27 (0.09, 0.75)** | **1.44 (1.08, 1.91)** | 1.26 (0.97, 1.64) | **1.35 (1.07, 1.70)** | 1.20 (0.99, 1.45) | **0.25 (0.10, 0.60)** | **0.27 (0.09, 0.75)** | **1.45 (1.08, 1.93)** | 1.26 (0.97, 1.64) | **1.35 (1.07, 1.70)** | 1.20 (1.00, 1.46) |
| HMPV | 0.35 (0.05, 2.44) | — | 1.01 (0.49, 2.10) | **2.05 (1.21, 3.46)** | 1.01 (0.40, 2.52) | 0.92 (0.36, 2.39) | 0.34 (0.05, 2.46) | — | 1.02 (0.49, 2.13) | **2.06 (1.22, 3.48)** | 1.01 (0.40, 2.52) | 0.93 (0.35, 2.46) |
| HPIV | 0.33 (0.05, 2.35) | — | **1.85 (1.04, 3.31)** | 0.81 (0.44, 1.48) | 0.86 (0.48, 1.52) | 0.95 (0.62, 1.47) | 0.32 (0.04, 2.26) | — | **1.89 (1.06, 3.37)** | 0.82 (0.45, 1.50) | 0.88 (0.50, 1.55) | 0.96 (0.62, 1.48) |
| **Outcome: HMPV** | | | | | | | | | | | | |
| SC2 | 0.51 (0.07, 3.58) | — | 0.79 (0.29, 2.11) | 1.37 (0.79, 2.39) | 0.74 (0.33, 1.63) | 0.91 (0.53, 1.57) | 0.79 (0.20, 3.20) | — | 0.78 (0.32, 1.87) | 1.54 (0.95, 2.50) | 0.96 (0.49, 1.86) | 0.84 (0.51, 1.39) |
| RSV | — | — | 0.43 (0.06, 3.23) | 0.60 (0.21, 1.74) | 0.92 (0.42, 2.02) | 1.10 (0.63, 1.91) | — | — | 0.36 (0.05, 2.64) | 0.51 (0.17, 1.49) | 0.96 (0.48, 1.91) | 1.20 (0.75, 1.94) |
| FLU | — | — | 0.80 (0.11, 5.99) | 0.57 (0.15, 2.27) | 0.97 (0.46, 2.06) | 1.58 (0.98, 2.56) | — | — | 0.57 (0.08, 4.06) | 0.43 (0.06, 2.89) | 0.85 (0.42, 1.75) | 1.34 (0.84, 2.13) |
| ADV | — | — | 0.95 (0.34, 2.60) | 0.84 (0.33, 2.16) | 0.69 (0.30, 1.60) | 0.97 (0.55, 1.73) | — | — | 0.96 (0.35, 2.64) | 0.84 (0.33, 2.15) | 0.67 (0.29, 1.57) | 0.96 (0.54, 1.70) |
| EV | — | — | 0.78 (0.24, 2.49) | 0.68 (0.22, 2.12) | 1.79 (0.88, 3.62) | 1.01 (0.63, 1.62) | — | — | 0.78 (0.24, 2.51) | 0.69 (0.22, 2.13) | 1.82 (0.90, 3.67) | 1.00 (0.62, 1.61) |
| RV | 0.48 (0.14, 1.62) | — | 1.11 (0.69, 1.78) | 1.16 (0.77, 1.72) | 1.27 (0.85, 1.91) | 1.23 (0.90, 1.69) | 0.48 (0.14, 1.61) | — | 1.11 (0.69, 1.78) | 1.15 (0.77, 1.72) | 1.26 (0.84, 1.90) | 1.23 (0.90, 1.69) |
| HCOV | 0.47 (0.12, 1.88) | 0.22 (0.03, 1.60) | 0.94 (0.51, 1.75) | 1.44 (0.92, 2.25) | **1.91 (1.11, 3.28)** | 1.04 (0.44, 2.48) | 0.47 (0.12, 1.88) | 0.22 (0.03, 1.59) | 0.95 (0.51, 1.76) | 1.44 (0.92, 2.25) | **1.87 (1.09, 3.22)** | 1.03 (0.43, 2.44) |
| HPIV | — | 0.70 (0.10, 5.09) | **2.63 (1.34, 5.17)** | 1.34 (0.64, 2.84) | 0.96 (0.31, 2.99) | 1.65 (0.92, 2.96) | — | 0.70 (0.10, 5.04) | **2.59 (1.31, 5.11)** | 1.35 (0.64, 2.85) | 0.97 (0.31, 2.98) | 1.61 (0.90, 2.88) |
| **Outcome: HPIV** | | | | | | | | | | | | |
| SC2 | **3.21 (1.61, 6.42)** | 0.88 (0.22, 3.50) | 1.11 (0.54, 2.28) | 1.17 (0.66, 2.07) | 1.13 (0.64, 2.01) | 0.92 (0.58, 1.46) | 2.05 (0.98, 4.29) | 0.93 (0.31, 2.84) | 1.10 (0.57, 2.09) | 1.09 (0.65, 1.83) | 1.01 (0.60, 1.71) | 0.77 (0.50, 1.19) |
| RSV | 0.56 (0.08, 3.99) | — | 1.06 (0.36, 3.11) | 1.75 (0.93, 3.28) | 0.97 (0.42, 2.25) | 1.20 (0.70, 2.08) | 1.33 (0.42, 4.24) | 0.97 (0.24, 3.90) | 0.75 (0.27, 2.07) | 1.81 (0.99, 3.32) | 1.29 (0.65, 2.58) | 1.33 (0.81, 2.17) |
| FLU | — | — | 2.48 (0.86, 7.11) | 0.85 (0.33, 2.24) | **2.00 (1.04, 3.86)** | 1.14 (0.61, 2.13) | — | — | 1.91 (0.67, 5.43) | 0.68 (0.26, 1.76) | **1.97 (1.05, 3.71)** | 0.97 (0.53, 1.77) |
| ADV | 1.07 (0.26, 4.46) | — | 1.50 (0.78, 2.87) | 1.71 (0.96, 3.05) | **1.98 (1.14, 3.44)** | 1.54 (0.95, 2.49) | 1.07 (0.25, 4.49) | — | 1.53 (0.81, 2.92) | 1.73 (0.98, 3.07) | **1.97 (1.13, 3.43)** | 1.53 (0.95, 2.47) |
| EV | — | — | 0.82 (0.38, 1.76) | 1.37 (0.79, 2.38) | 1.10 (0.59, 2.05) | 0.95 (0.55, 1.63) | — | — | 0.81 (0.38, 1.73) | 1.35 (0.78, 2.33) | 1.09 (0.58, 2.04) | 0.94 (0.55, 1.62) |
| RV | **0.07 (0.01, 0.48)** | — | **1.45 (1.04, 2.03)** | 1.18 (0.87, 1.60) | 1.03 (0.72, 1.47) | 0.91 (0.68, 1.21) | **0.07 (0.01, 0.48)** | — | **1.46 (1.04, 2.03)** | 1.19 (0.88, 1.60) | 1.03 (0.72, 1.47) | 0.91 (0.68, 1.21) |
| HCOV | 0.51 (0.12, 2.07) | 0.29 (0.04, 2.02) | **1.83 (1.01, 3.30)** | 0.77 (0.43, 1.41) | 1.05 (0.63, 1.72) | 1.33 (0.91, 1.95) | 0.50 (0.12, 2.06) | 0.29 (0.04, 2.00) | 1.77 (0.98, 3.21) | 0.77 (0.42, 1.39) | 1.04 (0.63, 1.72) | 1.34 (0.91, 1.95) |
| HMPV | — | — | 0.78 (0.25, 2.46) | 1.63 (0.80, 3.34) | 0.37 (0.09, 1.53) | 1.46 (0.85, 2.52) | — | — | 0.79 (0.25, 2.46) | 1.64 (0.81, 3.34) | 0.38 (0.09, 1.55) | 1.48 (0.86, 2.54) |

Columns within each block give exposure windows in days. Cell shading reflects the hazard ratio: navy below 1, white at 1, red above 1, clamped at 4. Bold indicates $p < 0.05$ ; em dashes denote cells where the hazard ratio was not estimable. Blank cells were not fitted in that model.

#### Supplemental Table 3: Age-specific HRs and 95% CI estimates for children 6 months to 4 years old.

Adjusted hazard ratios (aHRs) and 95% confidence intervals are from Cox proportional hazards models with time-varying indicator covariates for each pathogen exposure window (1–7, 8–14, 15–30, 31–60, 61–90, and 91–180 days after first positive swab) and an interaction term between exposure and age category, where the reference category is children aged 6 months–4 years old. Each row represents a distinct exposure pathogen and each outcome pathogen is shown in a separate block. The left panel includes symptomatic infections only; the right panel additionally includes asymptomatic infections for SC2, FLU, and RSV. Cell shading reflects the magnitude of the aHR: navy indicates values below 1, white indicates 1, and red indicates values above 1 (clamped at 4). Bold indicates p<0.05; em dashes denote cells where the aHR was not estimable; blank cells indicate the model was not fitted for that exposure–outcome combination. Abbreviations: SC2, SARS-CoV-2; FLU, influenza; RSV, respiratory syncytial virus; ADV, adenovirus; EV, enterovirus; RV, rhinovirus; HCOV, seasonal coronavirus; HMPV, human metapneumovirus; HPIV, human parainfluenza virus.

#### Age-specific hazard ratios: < 5 yr (1 of 2)

Hazard of symptomatic infection with a second pathogen

| | Symptomatic infections only | | | | | | Asymptomatic infections included (SC2, FLU, RSV) | | | | | |
|---|---|---|---|---|---|---|---|---|---|---|---|---|
| **Exposure** | **1–7** | **8–14** | **15–30** | **31–60** | **61–90** | **91–180** | **1–7** | **8–14** | **15–30** | **31–60** | **61–90** | **91–180** |
| **Outcome: RSV** | | | | | | | | | | | | |
| SC2 | — | — | 1.51 (0.29, 7.80) | 0.40 (0.03, 5.39) | 0.67 (0.08, 5.35) | 1.03 (0.26, 4.09) | — | 1.75 (0.21, 14.53) | 1.15 (0.22, 5.85) | 0.69 (0.12, 4.07) | 0.47 (0.10, 2.31) | 1.50 (0.60, 3.75) |
| FLU | — | — | — | 2.01 (0.30, 13.46) | **21.28 (5.11, 88.61)** | 5.04 (0.42, 60.59) | — | — | — | 1.69 (0.22, 13.26) | **12.05 (2.96, 49.10)** | — |
| ADV | 1.23 (0.15, 10.01) | — | 0.63 (0.17, 2.38) | 1.74 (0.75, 4.04) | 1.49 (0.54, 4.09) | 0.68 (0.18, 2.65) | 1.93 (0.51, 7.34) | — | 0.80 (0.25, 2.58) | 1.63 (0.71, 3.71) | 1.61 (0.55, 4.76) | 0.75 (0.26, 2.13) |
| EV | — | — | 1.96 (0.93, 4.14) | 1.34 (0.62, 2.87) | 1.54 (0.72, 3.28) | 1.00 (0.33, 3.02) | 2.67 (0.68, 10.38) | 2.08 (0.75, 5.75) | 1.60 (0.76, 3.34) | 1.19 (0.57, 2.46) | 1.20 (0.55, 2.59) | 0.95 (0.39, 2.30) |
| RV | 0.20 (0.03, 1.51) | — | 1.66 (0.94, 2.93) | 1.12 (0.65, 1.96) | 0.61 (0.31, 1.19) | 0.98 (0.48, 2.00) | 0.26 (0.07, 1.02) | 1.35 (0.68, 2.69) | **2.29 (1.39, 3.77)** | 1.37 (0.83, 2.26) | 0.61 (0.30, 1.22) | 1.11 (0.59, 2.06) |
| HCOV | — | — | 0.79 (0.10, 6.00) | **3.81 (1.24, 11.68)** | — | — | — | — | 0.59 (0.07, 4.70) | 3.24 (0.99, 10.56) | **0.02 (0.00, 0.10)** | **0.04 (0.01, 0.22)** |
| HMPV | — | — | — | — | 2.10 (0.24, 18.18) | — | — | — | — | — | 1.65 (0.20, 13.59) | — |
| HPIV | — | — | 0.73 (0.17, 3.18) | **2.22 (1.02, 4.83)** | **2.73 (1.05, 7.12)** | 2.27 (0.97, 5.33) | — | 0.72 (0.09, 5.66) | 0.33 (0.05, 2.39) | **2.31 (1.16, 4.63)** | 1.90 (0.70, 5.14) | 2.17 (0.96, 4.95) |
| **Outcome: SC2** | | | | | | | | | | | | |
| RSV | — | — | — | 0.43 (0.07, 2.67) | 1.03 (0.27, 3.84) | 1.05 (0.37, 2.93) | — | 1.20 (0.15, 9.55) | 0.53 (0.07, 4.00) | 1.21 (0.43, 3.44) | 1.04 (0.35, 3.12) | 1.24 (0.50, 3.08) |
| FLU | **15.92 (2.00, 126.96)** | — | 4.90 (0.72, 33.30) | **5.50 (1.37, 22.03)** | — | 2.56 (0.63, 10.49) | 6.29 (0.80, 49.49) | 0.13 (0.02, 1.04) | 2.57 (0.40, 16.50) | **4.81 (1.53, 15.11)** | — | **3.81 (1.03, 14.13)** |
| ADV | 3.24 (0.57, 18.56) | 1.19 (0.18, 7.78) | **4.52 (1.95, 10.48)** | 0.80 (0.29, 2.26) | 0.72 (0.21, 2.53) | 0.63 (0.26, 1.56) | 2.70 (0.50, 14.53) | 2.49 (0.80, 7.74) | **2.97 (1.23, 7.17)** | 0.85 (0.31, 2.30) | 0.87 (0.31, 2.41) | 0.65 (0.31, 1.36) |
| EV | 1.73 (0.15, 19.76) | — | 0.51 (0.07, 3.50) | 1.12 (0.25, 4.95) | 0.59 (0.06, 5.66) | 2.00 (0.97, 4.12) | 1.89 (0.19, 19.37) | — | 0.95 (0.24, 3.72) | 0.72 (0.17, 3.02) | 0.66 (0.11, 3.96) | 1.47 (0.74, 2.90) |
| RV | 0.55 (0.10, 3.12) | 0.36 (0.04, 2.97) | 0.81 (0.34, 1.90) | 0.66 (0.25, 1.77) | 1.35 (0.58, 3.10) | 1.28 (0.75, 2.19) | 0.22 (0.02, 1.94) | 1.35 (0.49, 3.75) | 0.79 (0.38, 1.64) | 0.94 (0.40, 2.23) | 1.24 (0.55, 2.75) | 1.42 (0.85, 2.35) |
| HCOV | — | 3.41 (0.77, 15.04) | 0.73 (0.19, 2.86) | 1.42 (0.42, 4.83) | 0.68 (0.13, 3.53) | 0.93 (0.31, 2.78) | — | 1.63 (0.35, 7.64) | 0.58 (0.16, 2.18) | 1.44 (0.40, 5.25) | 0.56 (0.13, 2.44) | 1.15 (0.47, 2.80) |
| HMPV | — | — | — | 0.94 (0.11, 7.79) | 2.53 (0.51, 12.59) | 0.98 (0.29, 3.28) | — | — | — | 0.66 (0.08, 5.30) | 1.70 (0.36, 8.11) | 0.71 (0.22, 2.29) |
| HPIV | — | — | 3.02 (0.87, 10.43) | 1.13 (0.29, 4.38) | 2.46 (0.68, 8.89) | 2.02 (0.92, 4.45) | — | — | **3.59 (1.38, 9.36)** | 0.51 (0.14, 1.82) | 2.37 (0.92, 6.09) | 1.77 (0.85, 3.67) |
| **Outcome: FLU** | | | | | | | | | | | | |
| RSV | — | 2.01 (0.41, 9.82) | 2.28 (0.61, 8.52) | 1.74 (0.58, 5.24) | **0.04 (0.01, 0.19)** | **0.28 (0.08, 0.91)** | — | 1.29 (0.23, 7.32) | 1.38 (0.41, 4.68) | 1.50 (0.60, 3.78) | 1.48 (0.39, 5.57) | **0.32 (0.13, 0.79)** |
| SC2 | — | — | — | **6.92 (1.41, 33.95)** | — | 2.81 (0.73, 10.88) | — | — | 1.33 (0.39, 4.48) | 1.48 (0.14, 15.57) | **0.10 (0.02, 0.56)** | 0.79 (0.20, 3.16) |
| ADV | **10.35 (1.76, 60.77)** | **7.41 (1.08, 50.81)** | 0.39 (0.10, 1.51) | 3.37 (0.99, 11.46) | 1.00 (0.34, 2.88) | 0.58 (0.21, 1.57) | **6.45 (1.02, 40.73)** | 2.98 (0.32, 27.41) | 1.22 (0.23, 6.47) | **4.61 (1.50, 14.22)** | 0.85 (0.29, 2.45) | 0.65 (0.16, 2.58) |
| EV | — | — | — | 0.33 (0.09, 1.19) | 0.33 (0.04, 2.58) | **0.05 (0.01, 0.17)** | — | — | — | 0.50 (0.17, 1.42) | 0.57 (0.13, 2.45) | 0.21 (0.04, 1.05) |
| RV | — | — | **4.81 (1.22, 18.91)** | **7.25 (2.47, 21.24)** | 3.30 (0.95, 11.40) | **3.95 (1.41, 11.06)** | — | 0.80 (0.08, 7.91) | 2.88 (0.92, 9.02) | **4.14 (1.86, 9.21)** | 2.03 (0.79, 5.24) | **3.03 (1.31, 7.02)** |
| HCOV | 1.96 (0.25, 15.74) | — | **8.68 (2.05, 36.84)** | — | — | — | 1.28 (0.14, 11.38) | 2.85 (0.31, 26.30) | 2.34 (0.29, 18.95) | **0.08 (0.01, 0.47)** | **0.09 (0.02, 0.53)** | 1.50 (0.12, 19.62) |
| HMPV | — | — | — | — | — | 5.35 (0.81, 35.25) | — | — | — | — | — | **7.56 (1.82, 31.46)** |
| HPIV | — | — | — | — | — | 1.20 (0.24, 5.96) | — | — | — | — | — | 1.31 (0.39, 4.46) |
| **Outcome: ADV** | | | | | | | | | | | | |
| RSV | 1.02 (0.13, 7.83) | — | 1.52 (0.55, 4.20) | 1.01 (0.40, 2.56) | 1.55 (0.55, 4.38) | 0.19 (0.03, 1.13) | 2.42 (0.68, 8.59) | — | 1.29 (0.45, 3.69) | 0.86 (0.32, 2.28) | 1.34 (0.50, 3.58) | 0.50 (0.15, 1.66) |
| SC2 | — | — | 0.92 (0.11, 7.69) | 0.70 (0.09, 5.69) | 0.51 (0.09, 3.07) | 2.03 (0.81, 5.08) | — | — | 1.46 (0.18, 11.66) | 1.03 (0.26, 4.02) | 0.57 (0.15, 2.20) | 1.68 (0.68, 4.14) |
| FLU | 3.18 (0.44, 22.90) | — | — | 1.08 (0.20, 5.78) | 3.23 (0.61, 17.15) | 2.06 (0.63, 6.72) | 2.09 (0.26, 16.82) | — | — | 0.91 (0.14, 5.76) | 2.21 (0.48, 10.12) | 1.37 (0.43, 4.36) |
| EV | — | — | 1.85 (0.76, 4.47) | 1.08 (0.42, 2.78) | 1.43 (0.62, 3.29) | 1.08 (0.48, 2.39) | — | — | 1.85 (0.75, 4.55) | 1.15 (0.45, 2.94) | 1.44 (0.62, 3.36) | 1.06 (0.46, 2.43) |
| RV | 0.37 (0.09, 1.42) | 0.17 (0.02, 1.25) | 1.28 (0.71, 2.32) | 0.67 (0.34, 1.32) | 1.00 (0.52, 1.93) | 0.88 (0.54, 1.43) | 0.34 (0.08, 1.38) | 0.16 (0.02, 1.23) | 1.26 (0.70, 2.26) | 0.66 (0.34, 1.29) | 1.03 (0.54, 1.96) | 0.89 (0.54, 1.46) |
| HCOV | — | 1.03 (0.13, 7.84) | 0.87 (0.24, 3.18) | 1.48 (0.58, 3.80) | 1.88 (0.80, 4.41) | 0.62 (0.25, 1.54) | — | 1.00 (0.13, 7.49) | 0.80 (0.22, 2.95) | 1.36 (0.53, 3.50) | 1.77 (0.74, 4.24) | 0.61 (0.24, 1.54) |
| HMPV | — | — | 0.87 (0.14, 5.22) | 1.16 (0.26, 5.24) | 0.97 (0.21, 4.36) | 0.49 (0.11, 2.13) | — | — | 0.88 (0.13, 5.73) | 1.16 (0.27, 5.05) | 0.94 (0.21, 4.16) | 0.49 (0.11, 2.13) |
| HPIV | — | 0.94 (0.13, 6.90) | 0.67 (0.17, 2.64) | 0.91 (0.33, 2.52) | 1.38 (0.54, 3.52) | 1.25 (0.59, 2.65) | — | 0.92 (0.12, 6.85) | 0.71 (0.18, 2.76) | 0.90 (0.32, 2.51) | 1.41 (0.56, 3.54) | 1.29 (0.62, 2.69) |

Columns within each block give exposure windows in days. Cell shading reflects the hazard ratio: navy below 1, white at 1, red above 1, clamped at 4. Bold indicates $p < 0.05$ ; em dashes denote cells where the hazard ratio was not estimable. Blank cells were not fitted in that model.

#### Age-specific hazard ratios: < 5 yr (2 of 2)

Hazard of symptomatic infection with a second pathogen

| | Symptomatic infections only | | | | | | Asymptomatic infections included (SC2, FLU, RSV) | | | | | |
|---|---|---|---|---|---|---|---|---|---|---|---|---|
| Exposure | 1–7 | 8–14 | 15–30 | 31–60 | 61–90 | 91–180 | 1–7 | 8–14 | 15–30 | 31–60 | 61–90 | 91–180 |
| **Outcome: EV** | | | | | | | | | | | | |
| RSV | — | — | 1.19 (0.34, 4.12) | 0.42 (0.06, 2.88) | 1.40 (0.36, 5.43) | 1.09 (0.45, 2.64) | 1.82 (0.47, 7.01) | — | 1.53 (0.50, 4.62) | 0.30 (0.05, 1.92) | 1.72 (0.61, 4.79) | 1.18 (0.52, 2.66) |
| SC2 | — | 3.23 (0.58, 17.94) | **3.85 (1.49, 9.96)** | 2.22 (0.91, 5.42) | **3.77 (1.32, 10.72)** | 1.72 (0.59, 5.00) | — | 3.94 (0.95, 16.39) | 2.17 (0.73, 6.48) | 1.75 (0.74, 4.13) | 2.43 (0.93, 6.34) | 1.33 (0.52, 3.43) |
| FLU | — | — | — | 2.26 (0.57, 9.02) | 2.27 (0.56, 9.12) | 0.51 (0.08, 3.33) | — | — | — | 2.33 (0.63, 8.69) | 2.22 (0.59, 8.43) | 0.42 (0.06, 2.83) |
| ADV | — | — | **2.30 (1.04, 5.11)** | 1.86 (0.93, 3.74) | 1.00 (0.37, 2.70) | 0.65 (0.24, 1.71) | — | — | **2.27 (1.01, 5.09)** | 1.98 (0.99, 3.99) | 0.98 (0.36, 2.68) | 0.61 (0.23, 1.61) |
| RV | 0.39 (0.10, 1.53) | — | 0.64 (0.31, 1.31) | 1.01 (0.55, 1.85) | 1.22 (0.67, 2.23) | 1.12 (0.69, 1.81) | 0.37 (0.09, 1.46) | — | 0.66 (0.32, 1.36) | 1.06 (0.58, 1.94) | 1.29 (0.72, 2.33) | 1.13 (0.70, 1.83) |
| HCOV | — | 1.59 (0.21, 11.89) | **2.50 (1.04, 5.97)** | 0.61 (0.16, 2.44) | 2.11 (0.80, 5.57) | **2.41 (1.28, 4.55)** | — | 1.64 (0.22, 12.11) | **2.55 (1.03, 6.27)** | 0.61 (0.15, 2.46) | 2.26 (0.87, 5.90) | **2.54 (1.36, 4.74)** |
| HMPV | — | — | — | 0.61 (0.08, 4.56) | 0.57 (0.12, 2.65) | 0.67 (0.22, 2.02) | — | — | — | 0.60 (0.08, 4.55) | 0.59 (0.10, 3.42) | 0.67 (0.22, 2.03) |
| HPIV | — | 1.12 (0.22, 5.56) | **2.66 (1.26, 5.58)** | 2.22 (0.97, 5.10) | 0.78 (0.24, 2.51) | 1.13 (0.58, 2.18) | — | 1.29 (0.20, 8.59) | **2.80 (1.34, 5.85)** | **2.32 (1.02, 5.28)** | 0.79 (0.25, 2.51) | 1.19 (0.61, 2.29) |
| **Outcome: RV** | | | | | | | | | | | | |
| RSV | 0.47 (0.09, 2.54) | — | 1.81 (0.80, 4.05) | 1.01 (0.30, 3.41) | 2.04 (0.67, 6.17) | 1.13 (0.35, 3.62) | 0.44 (0.08, 2.48) | — | 1.55 (0.69, 3.49) | 1.35 (0.51, 3.57) | 1.72 (0.59, 5.01) | 1.63 (0.63, 4.22) |
| SC2 | — | 1.84 (0.27, 12.58) | **5.66 (1.87, 17.19)** | 2.12 (0.63, 7.07) | — | 1.27 (0.25, 6.56) | — | 0.94 (0.13, 6.58) | **3.14 (1.19, 8.27)** | 1.29 (0.44, 3.79) | — | 0.62 (0.14, 2.88) |
| FLU | — | — | 0.73 (0.07, 7.58) | 0.48 (0.10, 2.21) | 1.29 (0.28, 5.94) | 3.66 (0.48, 28.05) | — | — | 0.97 (0.12, 7.61) | 0.43 (0.08, 2.38) | 0.79 (0.15, 4.05) | 2.33 (0.68, 7.97) |
| ADV | 1.15 (0.24, 5.52) | — | 1.92 (0.88, 4.16) | 0.92 (0.32, 2.63) | 2.26 (0.88, 5.75) | — | 1.08 (0.22, 5.24) | — | 2.05 (0.97, 4.34) | 1.01 (0.36, 2.78) | 2.47 (0.99, 6.21) | — |
| EV | — | — | 0.93 (0.29, 2.92) | 1.39 (0.56, 3.46) | 1.31 (0.35, 4.94) | 1.69 (0.60, 4.78) | — | — | 1.06 (0.35, 3.22) | 1.39 (0.56, 3.43) | 1.37 (0.38, 4.95) | 1.73 (0.59, 5.10) |
| HCOV | 0.43 (0.06, 2.87) | — | 0.87 (0.35, 2.17) | 1.26 (0.60, 2.65) | 0.73 (0.23, 2.33) | 0.85 (0.29, 2.45) | 0.44 (0.07, 2.95) | — | 0.91 (0.37, 2.26) | 1.32 (0.65, 2.70) | 0.72 (0.22, 2.36) | 0.98 (0.36, 2.62) |
| HMPV | 1.31 (0.19, 9.27) | — | 1.09 (0.29, 4.07) | 1.09 (0.37, 3.27) | 1.16 (0.29, 4.55) | 0.59 (0.09, 3.89) | 1.31 (0.19, 9.21) | — | 1.07 (0.29, 4.02) | 1.06 (0.36, 3.14) | 1.27 (0.33, 4.88) | 0.59 (0.08, 4.14) |
| HPIV | 0.38 (0.08, 1.87) | — | 1.05 (0.41, 2.69) | 1.42 (0.64, 3.19) | 2.04 (0.79, 5.26) | 0.57 (0.21, 1.60) | 0.37 (0.06, 2.17) | — | 1.06 (0.41, 2.73) | 1.47 (0.68, 3.19) | 2.02 (0.80, 5.07) | 0.63 (0.22, 1.76) |
| **Outcome: HCOV** | | | | | | | | | | | | |
| RSV | — | — | 0.99 (0.33, 2.99) | 1.51 (0.76, 2.98) | 1.74 (0.95, 3.19) | 1.43 (0.80, 2.56) | — | 1.09 (0.21, 5.71) | 0.86 (0.29, 2.58) | 1.55 (0.82, 2.95) | 1.33 (0.71, 2.48) | 1.16 (0.66, 2.02) |
| SC2 | 1.77 (0.23, 13.75) | 2.17 (0.32, 14.69) | 0.54 (0.08, 3.56) | 1.76 (0.68, 4.59) | 0.54 (0.08, 3.64) | 1.39 (0.47, 4.07) | 1.08 (0.14, 8.52) | 2.94 (0.71, 12.16) | 0.56 (0.10, 3.23) | 1.91 (0.82, 4.41) | 0.44 (0.07, 2.72) | 1.71 (0.75, 3.91) |
| FLU | — | — | 0.49 (0.06, 4.16) | 0.86 (0.25, 2.98) | 0.57 (0.16, 2.04) | 1.09 (0.39, 3.04) | — | 1.43 (0.18, 11.49) | 1.44 (0.40, 5.19) | 0.97 (0.32, 2.96) | 0.50 (0.13, 1.89) | 0.94 (0.33, 2.65) |
| ADV | — | — | **4.02 (2.23, 7.24)** | 1.35 (0.60, 3.02) | 1.46 (0.69, 3.07) | 1.14 (0.54, 2.41) | — | — | **4.04 (2.22, 7.32)** | 1.36 (0.63, 2.95) | 1.46 (0.68, 3.16) | 1.17 (0.56, 2.43) |
| EV | — | — | 1.32 (0.53, 3.33) | 0.69 (0.24, 1.99) | 1.48 (0.67, 3.27) | 1.32 (0.77, 2.25) | — | — | 1.30 (0.53, 3.19) | 0.67 (0.24, 1.88) | 1.38 (0.62, 3.09) | 1.31 (0.76, 2.26) |
| RV | 0.16 (0.02, 1.20) | **0.16 (0.03, 0.99)** | 1.63 (0.99, 2.68) | 1.46 (0.85, 2.53) | 1.59 (0.92, 2.73) | 1.04 (0.65, 1.68) | 0.17 (0.02, 1.26) | 0.16 (0.02, 1.07) | **1.66 (1.00, 2.74)** | 1.45 (0.84, 2.51) | 1.60 (0.93, 2.76) | 1.07 (0.67, 1.73) |
| HMPV | — | — | — | 1.47 (0.44, 4.92) | — | 0.49 (0.08, 2.92) | — | — | — | 1.40 (0.41, 4.82) | — | 0.49 (0.08, 2.82) |
| HPIV | — | — | 1.48 (0.61, 3.59) | 0.98 (0.34, 2.79) | 0.57 (0.19, 1.69) | 0.99 (0.54, 1.81) | — | — | 1.51 (0.62, 3.69) | 1.01 (0.35, 2.90) | 0.57 (0.19, 1.72) | 0.96 (0.53, 1.73) |
| **Outcome: HMPV** | | | | | | | | | | | | |
| RSV | — | — | — | 1.38 (0.43, 4.43) | 0.68 (0.15, 3.10) | 0.67 (0.23, 1.92) | — | — | — | 0.86 (0.20, 3.71) | 0.84 (0.25, 2.88) | 1.09 (0.48, 2.48) |
| SC2 | — | — | — | 1.16 (0.27, 5.06) | 0.61 (0.08, 4.69) | 0.36 (0.06, 2.31) | — | — | — | 1.59 (0.50, 5.08) | 0.54 (0.07, 4.01) | 0.32 (0.05, 2.21) |
| FLU | — | — | — | — | — | 1.82 (0.60, 5.54) | — | — | — | — | — | 1.28 (0.40, 4.02) |
| ADV | — | — | 1.27 (0.35, 4.66) | 0.70 (0.22, 2.28) | 0.52 (0.16, 1.62) | 1.65 (0.70, 3.86) | — | — | 1.32 (0.36, 4.79) | 0.68 (0.21, 2.23) | 0.48 (0.15, 1.51) | 1.64 (0.69, 3.89) |
| EV | — | — | 0.87 (0.21, 3.60) | 0.50 (0.04, 7.18) | 1.51 (0.45, 5.00) | 1.00 (0.44, 2.27) | — | — | 0.91 (0.22, 3.70) | 0.51 (0.04, 6.78) | 1.64 (0.51, 5.25) | 0.98 (0.42, 2.30) |
| RV | — | — | 1.95 (0.98, 3.85) | 1.24 (0.67, 2.27) | 1.28 (0.59, 2.81) | 1.10 (0.55, 2.19) | — | — | 1.87 (0.95, 3.68) | 1.20 (0.66, 2.18) | 1.17 (0.52, 2.63) | 1.01 (0.51, 1.99) |
| HCOV | — | — | 0.47 (0.11, 2.03) | 1.55 (0.66, 3.67) | 1.62 (0.50, 5.25) | 1.46 (0.40, 5.31) | — | — | 0.45 (0.10, 1.94) | 1.43 (0.60, 3.41) | 1.59 (0.48, 5.25) | 1.43 (0.39, 5.26) |
| HPIV | — | 1.35 (0.17, 10.36) | 2.59 (0.98, 6.87) | 0.64 (0.19, 2.15) | 0.52 (0.04, 6.21) | 1.15 (0.48, 2.76) | — | 1.28 (0.17, 9.95) | 2.48 (0.93, 6.64) | 0.67 (0.19, 2.37) | 0.52 (0.03, 7.93) | 1.12 (0.48, 2.61) |
| **Outcome: HPIV** | | | | | | | | | | | | |
| RSV | — | — | 0.35 (0.05, 2.54) | 2.06 (0.93, 4.58) | 0.55 (0.12, 2.57) | 0.79 (0.35, 1.80) | — | 0.85 (0.12, 6.02) | 0.31 (0.04, 2.27) | 2.13 (0.93, 4.84) | 0.80 (0.23, 2.83) | 0.95 (0.46, 1.96) |
| SC2 | **9.54 (3.07, 29.62)** | — | **3.43 (1.05, 11.22)** | 1.82 (0.52, 6.33) | — | 0.32 (0.04, 2.51) | **4.14 (1.15, 14.87)** | — | **3.15 (1.16, 8.52)** | 1.40 (0.43, 4.62) | 0.37 (0.06, 2.30) | 0.23 (0.03, 1.76) |
| FLU | — | — | 1.69 (0.23, 12.30) | 1.24 (0.34, 4.59) | **4.52 (1.70, 12.04)** | 2.34 (0.84, 6.50) | — | — | 1.43 (0.19, 10.60) | 0.99 (0.26, 3.86) | **4.18 (1.75, 9.97)** | 1.79 (0.64, 4.95) |
| ADV | — | — | 1.14 (0.45, 2.87) | 1.09 (0.37, 3.16) | 1.67 (0.68, 4.10) | 1.69 (0.97, 2.94) | — | — | 1.29 (0.54, 3.09) | 1.14 (0.40, 3.23) | 1.61 (0.67, 3.86) | 1.62 (0.94, 2.77) |
| EV | — | — | 0.99 (0.32, 3.01) | 1.70 (0.77, 3.75) | 1.01 (0.42, 2.45) | 1.17 (0.55, 2.48) | — | — | 1.02 (0.35, 2.97) | 1.60 (0.73, 3.53) | 0.97 (0.39, 2.42) | 1.14 (0.53, 2.44) |
| RV | — | — | **1.68 (1.01, 2.79)** | 1.23 (0.73, 2.07) | 0.90 (0.49, 1.65) | 0.88 (0.53, 1.46) | — | — | **1.66 (1.00, 2.76)** | 1.21 (0.72, 2.05) | 0.89 (0.48, 1.65) | 0.88 (0.53, 1.45) |
| HCOV | 1.36 (0.40, 4.58) | — | 1.99 (0.79, 5.00) | 0.31 (0.09, 1.16) | 0.81 (0.35, 1.90) | 0.85 (0.38, 1.89) | 1.44 (0.38, 5.40) | — | 1.86 (0.76, 4.52) | 0.33 (0.09, 1.17) | 0.78 (0.32, 1.87) | 0.87 (0.40, 1.89) |
| HMPV | — | — | 1.11 (0.25, 4.88) | 2.33 (0.96, 5.68) | 0.44 (0.06, 3.33) | 1.34 (0.55, 3.27) | — | — | 1.13 (0.27, 4.79) | 2.31 (0.96, 5.54) | 0.47 (0.06, 3.45) | 1.38 (0.56, 3.40) |

Columns within each block give exposure windows in days. Cell shading reflects the hazard ratio: navy below 1, white at 1, red above 1, clamped at 4. Bold indicates $p < 0.05$ ; em dashes denote cells where the hazard ratio was not estimable. Blank cells were not fitted in that model.

#### Supplemental Table 4: Age-specific HRs and 95% CI estimates for children 5–11 years old.

Adjusted hazard ratios (aHRs) and 95% confidence intervals are from Cox proportional hazards models with time-varying indicator covariates for each pathogen exposure window (1–7, 8–14, 15–30, 31–60, 61–90, and 91–180 days after first positive swab) and an interaction term between exposure and age category, where the reference category is children aged 5–11 years old. Each row represents a distinct exposure pathogen and each outcome pathogen is shown in a separate block. The left panel includes symptomatic infections only; the right panel additionally includes asymptomatic infections for SC2, FLU, and RSV. Cell shading reflects the magnitude of the aHR: navy indicates values below 1, white indicates 1, and red indicates values above 1 (clamped at 4). Bold indicates $p<0.05$; em dashes denote cells with complete separation of the exposure and outcome data, where the aHR was not estimable. Abbreviations: SC2, SARS-CoV-2; FLU, influenza; RSV, respiratory syncytial virus; ADV, adenovirus; EV, enterovirus; RV, rhinovirus; HCOV, seasonal coronavirus; HMPV, human metapneumovirus; HPIV, human parainfluenza virus.

#### Age-specific hazard ratios: 5-11 yr (1 of 2)

Hazard of symptomatic infection with a second pathogen

| | Symptomatic infections only | | | | | | Asymptomatic infections included (SC2, FLU, RSV) | | | | | |
|---|---|---|---|---|---|---|---|---|---|---|---|---|
| **Exposure** | **1–7** | **8–14** | **15–30** | **31–60** | **61–90** | **91–180** | **1–7** | **8–14** | **15–30** | **31–60** | **61–90** | **91–180** |
| **Outcome: RSV** | | | | | | | | | | | | |
| SC2 | — | — | — | 1.07 (0.35, 3.29) | 1.63 (0.67, 3.96) | 0.60 (0.14, 2.58) | 2.43 (0.77, 7.73) | 0.78 (0.11, 5.71) | 0.75 (0.19, 3.05) | 0.91 (0.38, 2.15) | 1.67 (0.89, 3.12) | 0.84 (0.40, 1.76) |
| FLU | — | — | 0.67 (0.10, 4.78) | 2.05 (0.60, 6.99) | — | — | — | 0.64 (0.09, 4.41) | 0.79 (0.19, 3.28) | **2.42 (1.02, 5.73)** | **0.07 (0.01, 0.35)** | 1.20 (0.14, 10.40) |
| ADV | — | — | 1.43 (0.41, 5.02) | **0.06 (0.01, 0.28)** | 0.67 (0.10, 4.55) | 0.40 (0.01, 21.89) | — | — | 1.69 (0.62, 4.63) | 0.42 (0.06, 2.91) | 0.55 (0.09, 3.48) | 0.63 (0.10, 4.13) |
| EV | — | — | 1.23 (0.40, 3.78) | 1.47 (0.51, 4.26) | 2.10 (0.79, 5.56) | 0.29 (0.06, 1.35) | 3.00 (0.83, 10.91) | 2.10 (0.45, 9.92) | 1.52 (0.54, 4.31) | 1.22 (0.49, 3.04) | 1.50 (0.61, 3.70) | 0.92 (0.33, 2.58) |
| RV | — | 0.47 (0.11, 1.92) | 1.35 (0.80, 2.28) | 0.53 (0.26, 1.07) | 0.51 (0.25, 1.07) | 1.04 (0.61, 1.77) | 0.55 (0.22, 1.37) | 1.00 (0.43, 2.31) | 1.11 (0.67, 1.85) | 0.62 (0.35, 1.11) | 0.59 (0.32, 1.10) | 0.89 (0.55, 1.43) |
| HCOV | — | — | 1.06 (0.15, 7.80) | 1.59 (0.25, 10.03) | **6.11 (1.40, 26.71)** | 1.18 (0.22, 6.25) | — | 1.44 (0.20, 10.39) | **0.04 (0.01, 0.27)** | 1.07 (0.14, 8.26) | **4.60 (1.07, 19.67)** | 2.12 (0.67, 6.73) |
| HMPV | — | — | | 1.01 (0.11, 8.89) | — | — | — | — | — | 0.66 (0.08, 5.43) | — | — |
| HPIV | — | — | 0.66 (0.09, 4.78) | **2.70 (1.14, 6.39)** | **4.60 (2.02, 10.48)** | **2.98 (1.19, 7.44)** | — | — | 1.52 (0.50, 4.67) | **2.98 (1.50, 5.90)** | **4.06 (1.87, 8.84)** | **2.41 (1.13, 5.14)** |
| **Outcome: SC2** | | | | | | | | | | | | |
| RSV | — | — | 1.06 (0.26, 4.29) | 1.05 (0.38, 2.94) | 0.42 (0.12, 1.50) | **2.43 (1.36, 4.32)** | — | 1.22 (0.29, 5.03) | 0.57 (0.15, 2.22) | 1.01 (0.48, 2.14) | 0.84 (0.34, 2.05) | **1.87 (1.08, 3.22)** |
| FLU | — | 0.69 (0.30, 1.61) | 0.89 (0.22, 3.62) | **2.38 (1.03, 5.51)** | **0.05 (0.01, 0.27)** | 0.75 (0.27, 2.10) | 0.71 (0.10, 4.96) | 1.12 (0.32, 3.87) | 0.80 (0.27, 2.43) | **1.96 (1.04, 3.69)** | 0.25 (0.04, 1.78) | 0.68 (0.31, 1.51) |
| ADV | — | — | — | 0.47 (0.11, 1.93) | 0.27 (0.05, 1.38) | 0.73 (0.35, 1.51) | — | 2.69 (0.89, 8.07) | 0.78 (0.23, 2.70) | 0.59 (0.21, 1.68) | 0.72 (0.27, 1.91) | 0.84 (0.45, 1.56) |
| EV | — | — | 0.92 (0.27, 3.18) | 1.11 (0.46, 2.67) | 0.14 (0.01, 1.72) | 1.07 (0.60, 1.89) | — | 0.81 (0.10, 6.31) | 0.63 (0.17, 2.33) | 1.13 (0.50, 2.53) | 0.24 (0.04, 1.60) | 0.79 (0.46, 1.36) |
| RV | 0.73 (0.27, 1.96) | 0.52 (0.16, 1.65) | 0.83 (0.46, 1.51) | 1.27 (0.82, 1.99) | 1.12 (0.63, 1.98) | 1.21 (0.87, 1.68) | 1.00 (0.48, 2.12) | 0.53 (0.21, 1.29) | 1.12 (0.71, 1.78) | 1.09 (0.72, 1.65) | 1.34 (0.87, 2.08) | 1.21 (0.91, 1.62) |
| HCOV | — | 0.50 (0.07, 3.55) | 0.77 (0.24, 2.48) | 1.19 (0.56, 2.55) | 0.54 (0.14, 2.07) | 1.38 (0.82, 2.32) | — | 0.68 (0.17, 2.71) | 1.26 (0.59, 2.71) | 0.88 (0.42, 1.85) | 0.54 (0.18, 1.62) | 0.97 (0.58, 1.62) |
| HMPV | — | 1.14 (0.44, 2.93) | 1.46 (0.36, 5.87) | 1.41 (0.44, 4.59) | 1.21 (0.31, 4.66) | 0.89 (0.36, 2.21) | — | 1.13 (0.23, 5.44) | 1.79 (0.57, 5.59) | 0.77 (0.19, 3.17) | 0.94 (0.24, 3.65) | 0.98 (0.47, 2.06) |
| HPIV | — | — | — | 0.54 (0.13, 2.20) | **2.14 (1.08, 4.22)** | 1.52 (0.90, 2.58) | — | 1.29 (0.31, 5.47) | **0.07 (0.02, 0.22)** | 0.51 (0.16, 1.66) | 1.55 (0.82, 2.93) | 1.35 (0.82, 2.22) |
| **Outcome: FLU** | | | | | | | | | | | | |
| RSV | — | — | 0.91 (0.34, 2.43) | 1.38 (0.60, 3.20) | 2.61 (0.79, 8.66) | 1.73 (0.40, 7.57) | — | 0.40 (0.05, 2.95) | 1.67 (0.90, 3.09) | 1.72 (0.98, 3.02) | 2.05 (0.83, 5.05) | 1.32 (0.42, 4.12) |
| SC2 | 1.25 (0.17, 9.36) | 2.98 (0.71, 12.47) | 2.44 (0.89, 6.68) | 1.60 (0.59, 4.31) | 1.07 (0.24, 4.72) | 0.76 (0.31, 1.87) | 2.22 (0.68, 7.27) | **3.44 (1.26, 9.44)** | 2.12 (0.93, 4.83) | 1.45 (0.67, 3.15) | 0.84 (0.26, 2.63) | 1.12 (0.62, 2.02) |
| ADV | — | — | 0.98 (0.31, 3.08) | 0.42 (0.06, 2.74) | 0.70 (0.11, 4.62) | 0.78 (0.24, 2.54) | 0.88 (0.12, 6.39) | 0.60 (0.15, 2.45) | 0.13 (0.01, 2.96) | 0.76 (0.25, 2.31) | 0.54 (0.10, 2.93) | 0.98 (0.40, 2.43) |
| EV | — | — | 0.80 (0.17, 3.70) | 1.61 (0.69, 3.79) | 0.51 (0.20, 1.26) | 1.22 (0.52, 2.84) | 1.31 (0.17, 10.20) | — | 0.33 (0.04, 2.45) | 1.58 (0.77, 3.25) | 0.48 (0.20, 1.17) | 1.07 (0.52, 2.20) |
| RV | 1.06 (0.35, 3.25) | — | 1.62 (0.93, 2.83) | 1.19 (0.74, 1.92) | 0.89 (0.55, 1.45) | 1.00 (0.59, 1.69) | 1.33 (0.57, 3.11) | 0.96 (0.42, 2.19) | 1.42 (0.86, 2.35) | 1.08 (0.70, 1.66) | 0.95 (0.61, 1.48) | 1.19 (0.76, 1.86) |
| HCOV | — | — | — | 0.72 (0.10, 5.35) | 2.16 (0.32, 14.81) | 1.92 (0.48, 7.71) | 0.73 (0.10, 5.44) | — | **0.02 (0.00, 0.12)** | 0.41 (0.05, 3.26) | 1.29 (0.19, 8.73) | 1.68 (0.54, 5.22) |
| HMPV | — | — | — | 1.71 (0.41, 7.05) | **3.98 (1.13, 14.09)** | 1.89 (0.24, 14.82) | — | — | 1.80 (0.28, 11.55) | 1.94 (0.60, 6.24) | **4.57 (1.62, 12.89)** | 2.63 (0.63, 10.98) |
| HPIV | — | — | 0.49 (0.07, 3.21) | 0.63 (0.14, 2.77) | 1.42 (0.41, 4.87) | 0.40 (0.11, 1.54) | — | 0.96 (0.15, 6.36) | 0.43 (0.06, 3.07) | 1.12 (0.44, 2.83) | 1.67 (0.63, 4.44) | 0.52 (0.21, 1.33) |
| **Outcome: ADV** | | | | | | | | | | | | |
| RSV | — | — | 2.51 (0.78, 8.05) | — | 1.39 (0.34, 5.62) | 1.59 (0.70, 3.58) | — | — | 2.15 (0.68, 6.82) | 0.89 (0.22, 3.63) | 1.62 (0.53, 4.91) | 1.60 (0.72, 3.56) |
| SC2 | 2.78 (0.67, 11.46) | 3.33 (0.82, 13.45) | 1.41 (0.45, 4.45) | 1.85 (0.71, 4.82) | 0.51 (0.14, 1.90) | 1.41 (0.73, 2.73) | **5.12 (2.04, 12.84)** | 2.39 (0.59, 9.67) | 1.07 (0.33, 3.41) | 2.09 (0.97, 4.48) | 0.42 (0.13, 1.32) | 1.37 (0.77, 2.44) |
| FLU | — | — | — | 0.50 (0.07, 3.66) | **0.04 (0.00, 0.38)** | 1.50 (0.62, 3.63) | — | — | — | 0.71 (0.17, 3.02) | **0.03 (0.00, 0.21)** | 0.90 (0.32, 2.49) |
| EV | 1.62 (0.18, 14.62) | — | — | **2.60 (1.17, 5.80)** | 0.63 (0.18, 2.22) | 0.98 (0.43, 2.21) | 1.58 (0.18, 14.17) | — | — | **2.57 (1.16, 5.69)** | 0.64 (0.19, 2.21) | 1.00 (0.44, 2.23) |
| RV | 1.23 (0.44, 3.41) | 0.53 (0.12, 2.25) | **1.92 (1.11, 3.31)** | 1.14 (0.62, 2.11) | 1.04 (0.56, 1.91) | 1.07 (0.66, 1.74) | 1.21 (0.44, 3.33) | 0.54 (0.13, 2.29) | **1.97 (1.15, 3.39)** | 1.15 (0.63, 2.12) | 1.03 (0.56, 1.92) | 1.08 (0.66, 1.76) |
| HCOV | 0.84 (0.11, 6.27) | — | 1.13 (0.34, 3.75) | 1.10 (0.45, 2.73) | 0.83 (0.30, 2.25) | 1.08 (0.58, 2.02) | 0.82 (0.11, 6.12) | — | 1.10 (0.33, 3.65) | 1.11 (0.45, 2.73) | 0.83 (0.31, 2.26) | 1.10 (0.59, 2.04) |
| HMPV | — | — | 0.95 (0.13, 6.97) | **2.87 (1.17, 7.06)** | 1.14 (0.28, 4.60) | 0.70 (0.22, 2.22) | — | — | 1.04 (0.14, 7.52) | **3.08 (1.26, 7.54)** | 1.15 (0.28, 4.62) | 0.73 (0.23, 2.31) |
| HPIV | — | — | 1.40 (0.37, 5.28) | **2.74 (1.19, 6.30)** | 2.05 (0.76, 5.57) | **2.29 (1.28, 4.09)** | — | — | 1.47 (0.38, 5.62) | **2.75 (1.20, 6.31)** | 2.07 (0.77, 5.57) | **2.29 (1.27, 4.14)** |

Columns within each block give exposure windows in days. Cell shading reflects the hazard ratio: navy below 1, white at 1, red above 1, clamped at 4. Bold indicates $p < 0.05$ ; em dashes denote cells where the hazard ratio was not estimable. Blank cells were not fitted in that model.

#### Age-specific hazard ratios: 5-11 yr (2 of 2)

Hazard of symptomatic infection with a second pathogen

| | Symptomatic infections only | | | | | | Asymptomatic infections included (SC2, FLU, RSV) | | | | | |
|---|---|---|---|---|---|---|---|---|---|---|---|---|
| **Exposure** | **1–7** | **8–14** | **15–30** | **31–60** | **61–90** | **91–180** | **1–7** | **8–14** | **15–30** | **31–60** | **61–90** | **91–180** |
| **Outcome: EV** | | | | | | | | | | | | |
| RSV | — | — | — | 0.52 (0.07, 3.89) | 1.17 (0.28, 4.92) | 1.47 (0.59, 3.67) | — | — | — | 0.72 (0.18, 2.96) | 1.26 (0.39, 4.06) | 1.90 (0.82, 4.41) |
| SC2 | 1.60 (0.40, 6.49) | 0.98 (0.13, 7.13) | 2.11 (0.89, 5.02) | 1.00 (0.37, 2.71) | 1.53 (0.65, 3.61) | 0.92 (0.40, 2.10) | 0.60 (0.08, 4.25) | 1.20 (0.29, 4.94) | 2.03 (0.97, 4.26) | 1.20 (0.56, 2.55) | **2.12 (1.15, 3.90)** | 0.81 (0.40, 1.65) |
| FLU | — | — | — | 1.68 (0.53, 5.33) | **0.04 (0.01, 0.15)** | 0.56 (0.13, 2.37) | — | — | — | 1.99 (0.81, 4.90) | 0.45 (0.06, 3.12) | 0.87 (0.30, 2.50) |
| ADV | 3.11 (0.74, 13.06) | — | 0.81 (0.16, 4.07) | 1.24 (0.40, 3.87) | 1.50 (0.59, 3.81) | 0.63 (0.30, 1.32) | 3.23 (0.77, 13.49) | — | 0.80 (0.16, 4.04) | 1.17 (0.37, 3.63) | 1.45 (0.59, 3.58) | 0.63 (0.30, 1.32) |
| RV | 0.94 (0.41, 2.15) | — | 0.52 (0.23, 1.17) | 1.30 (0.77, 2.22) | 1.22 (0.67, 2.24) | 1.41 (0.95, 2.10) | 0.94 (0.41, 2.13) | — | 0.51 (0.22, 1.15) | 1.27 (0.75, 2.17) | 1.18 (0.65, 2.14) | 1.37 (0.92, 2.05) |
| HCOV | — | — | 0.42 (0.06, 3.17) | 0.83 (0.26, 2.70) | 1.00 (0.33, 3.00) | 1.22 (0.61, 2.46) | — | — | 0.43 (0.06, 3.17) | 0.79 (0.25, 2.52) | 0.95 (0.31, 2.87) | 1.20 (0.60, 2.40) |
| HMPV | — | — | **3.73 (1.14, 12.15)** | 0.70 (0.09, 5.11) | — | 0.50 (0.12, 2.09) | — | — | **3.73 (1.14, 12.16)** | 0.67 (0.09, 4.98) | — | 0.51 (0.12, 2.14) |
| HPIV | — | — | 1.46 (0.35, 6.08) | 1.28 (0.41, 3.99) | 0.42 (0.08, 2.30) | **2.18 (1.25, 3.81)** | — | — | 1.49 (0.36, 6.17) | 1.32 (0.42, 4.12) | 0.43 (0.08, 2.36) | **2.15 (1.24, 3.75)** |
| **Outcome: RV** | | | | | | | | | | | | |
| RSV | — | 0.67 (0.09, 4.93) | 0.91 (0.32, 2.62) | 1.53 (0.75, 3.13) | 0.73 (0.27, 1.97) | 1.08 (0.59, 1.99) | 0.45 (0.06, 3.15) | 0.50 (0.07, 3.63) | 0.69 (0.24, 2.03) | 1.42 (0.76, 2.64) | 0.87 (0.38, 1.98) | 1.21 (0.71, 2.09) |
| SC2 | 2.16 (0.94, 4.94) | 0.68 (0.16, 2.81) | 0.83 (0.38, 1.80) | 1.30 (0.70, 2.44) | 1.05 (0.52, 2.13) | 0.67 (0.36, 1.26) | 1.25 (0.51, 3.07) | 0.69 (0.22, 2.18) | 0.71 (0.35, 1.46) | 1.33 (0.81, 2.20) | 0.86 (0.47, 1.60) | 0.80 (0.50, 1.26) |
| FLU | — | — | 1.23 (0.38, 4.02) | 1.16 (0.55, 2.45) | 0.52 (0.13, 2.08) | 1.55 (0.84, 2.85) | — | — | 1.69 (0.70, 4.10) | 1.35 (0.72, 2.54) | 1.31 (0.58, 2.95) | 1.69 (0.99, 2.90) |
| ADV | — | — | 0.74 (0.19, 2.89) | 0.69 (0.22, 2.12) | 1.44 (0.65, 3.18) | 1.11 (0.52, 2.36) | — | — | 0.77 (0.19, 3.06) | 0.72 (0.23, 2.19) | 1.45 (0.65, 3.22) | 1.10 (0.52, 2.34) |
| EV | 0.70 (0.10, 5.17) | 0.82 (0.11, 6.02) | 1.62 (0.62, 4.26) | 1.23 (0.45, 3.34) | — | 1.43 (0.72, 2.85) | 0.73 (0.10, 5.36) | 0.83 (0.11, 6.10) | 1.60 (0.61, 4.21) | 1.22 (0.45, 3.29) | — | 1.35 (0.68, 2.66) |
| HCOV | 0.29 (0.04, 2.02) | — | 1.12 (0.56, 2.26) | 1.52 (0.93, 2.49) | 1.81 (0.97, 3.38) | 1.38 (0.86, 2.20) | 0.28 (0.04, 2.01) | — | 1.11 (0.55, 2.23) | 1.49 (0.91, 2.44) | 1.77 (0.95, 3.29) | 1.40 (0.87, 2.24) |
| HMPV | — | — | 1.80 (0.80, 4.04) | 0.85 (0.26, 2.80) | 0.22 (0.03, 1.59) | 0.57 (0.23, 1.42) | — | — | 1.78 (0.80, 3.98) | 0.85 (0.26, 2.80) | 0.21 (0.03, 1.51) | 0.57 (0.23, 1.43) |
| HPIV | — | — | 0.84 (0.31, 2.27) | 1.25 (0.62, 2.50) | 1.75 (0.91, 3.36) | 1.50 (0.88, 2.55) | — | — | 0.82 (0.30, 2.24) | 1.24 (0.62, 2.48) | 1.71 (0.89, 3.29) | 1.51 (0.89, 2.56) |
| **Outcome: HCOV** | | | | | | | | | | | | |
| RSV | — | — | 0.73 (0.19, 2.81) | 1.58 (0.86, 2.87) | 1.45 (0.77, 2.72) | 1.18 (0.67, 2.08) | 1.31 (0.18, 9.32) | — | 0.61 (0.16, 2.29) | 1.43 (0.82, 2.48) | 1.27 (0.72, 2.25) | 1.12 (0.68, 1.85) |
| SC2 | — | 0.62 (0.09, 4.34) | 1.54 (0.60, 3.98) | 1.35 (0.66, 2.78) | 1.37 (0.66, 2.86) | 1.07 (0.61, 1.86) | 0.40 (0.06, 2.85) | — | 1.60 (0.79, 3.22) | 0.91 (0.46, 1.80) | 0.85 (0.41, 1.76) | 0.91 (0.56, 1.49) |
| FLU | — | — | 0.77 (0.24, 2.45) | 1.03 (0.56, 1.89) | 0.75 (0.33, 1.74) | 1.03 (0.54, 1.97) | — | — | 1.17 (0.53, 2.57) | 1.23 (0.75, 2.01) | 0.93 (0.47, 1.86) | 0.82 (0.43, 1.56) |
| ADV | — | — | 1.75 (0.66, 4.62) | 1.72 (0.77, 3.87) | 0.87 (0.33, 2.26) | 1.25 (0.66, 2.36) | — | — | 1.79 (0.68, 4.75) | 1.73 (0.76, 3.91) | 0.88 (0.34, 2.30) | 1.23 (0.66, 2.32) |
| EV | — | — | 0.77 (0.19, 3.06) | 1.43 (0.57, 3.55) | 0.51 (0.10, 2.56) | 1.01 (0.61, 1.69) | — | — | 0.76 (0.19, 3.05) | 1.44 (0.58, 3.56) | 0.49 (0.09, 2.62) | 1.01 (0.61, 1.68) |
| RV | 0.15 (0.02, 1.07) | 0.16 (0.02, 1.13) | 1.54 (0.96, 2.47) | 1.03 (0.67, 1.58) | 1.31 (0.84, 2.04) | 1.31 (0.95, 1.82) | 0.15 (0.02, 1.06) | 0.16 (0.02, 1.17) | 1.55 (0.97, 2.48) | 1.05 (0.68, 1.61) | 1.32 (0.85, 2.07) | 1.33 (0.96, 1.84) |
| HMPV | — | — | 1.52 (0.59, 3.88) | 1.70 (0.70, 4.12) | 0.99 (0.24, 4.13) | 1.44 (0.49, 4.21) | — | — | 1.53 (0.60, 3.91) | 1.76 (0.72, 4.27) | 1.02 (0.25, 4.25) | 1.44 (0.48, 4.38) |
| HPIV | — | — | 2.26 (0.93, 5.47) | 0.39 (0.10, 1.50) | 0.75 (0.27, 2.05) | 1.01 (0.54, 1.87) | — | — | 2.27 (0.94, 5.50) | 0.40 (0.10, 1.52) | 0.76 (0.28, 2.11) | 1.02 (0.55, 1.89) |
| **Outcome: HMPV** | | | | | | | | | | | | |
| RSV | — | — | — | — | 0.96 (0.29, 3.14) | 1.25 (0.65, 2.43) | — | — | — | 0.43 (0.06, 3.20) | 1.10 (0.41, 2.97) | 1.21 (0.66, 2.22) |
| SC2 | — | — | — | 0.97 (0.31, 3.05) | 1.24 (0.40, 3.87) | 0.99 (0.39, 2.52) | — | — | 0.38 (0.05, 2.71) | 1.48 (0.68, 3.21) | 1.73 (0.69, 4.36) | 1.02 (0.47, 2.25) |
| FLU | — | — | 1.59 (0.21, 11.86) | 0.54 (0.08, 3.72) | 1.27 (0.51, 3.14) | 0.86 (0.36, 2.05) | — | — | 1.10 (0.16, 7.46) | 0.38 (0.02, 9.22) | 1.20 (0.52, 2.79) | 0.90 (0.42, 1.93) |
| ADV | — | — | 0.91 (0.14, 5.89) | 1.50 (0.36, 6.32) | 0.89 (0.21, 3.74) | 0.57 (0.20, 1.62) | — | — | 0.92 (0.14, 5.92) | 1.51 (0.36, 6.23) | 0.90 (0.21, 3.78) | 0.58 (0.20, 1.66) |
| EV | — | — | — | 0.92 (0.22, 3.89) | 2.32 (0.74, 7.30) | 1.09 (0.53, 2.24) | — | — | — | 0.93 (0.22, 4.00) | 2.34 (0.75, 7.28) | 1.12 (0.54, 2.29) |
| RV | 0.56 (0.13, 2.31) | — | 0.69 (0.27, 1.73) | 1.47 (0.80, 2.69) | 1.20 (0.66, 2.16) | 1.43 (0.92, 2.23) | 0.54 (0.13, 2.27) | — | 0.68 (0.27, 1.71) | 1.44 (0.78, 2.65) | 1.19 (0.67, 2.12) | 1.40 (0.90, 2.19) |
| HCOV | 1.28 (0.32, 5.12) | 0.57 (0.08, 4.09) | 0.91 (0.34, 2.47) | 1.30 (0.69, 2.44) | **2.31 (1.16, 4.60)** | 0.49 (0.05, 5.02) | 1.27 (0.32, 5.08) | 0.57 (0.08, 4.08) | 0.92 (0.34, 2.50) | 1.31 (0.70, 2.47) | **2.30 (1.16, 4.58)** | 0.48 (0.05, 5.00) |
| HPIV | — | — | 1.46 (0.35, 6.08) | 2.07 (0.77, 5.54) | 1.33 (0.35, 5.01) | 1.67 (0.78, 3.56) | — | — | 1.48 (0.35, 6.17) | 2.05 (0.76, 5.51) | 1.37 (0.36, 5.19) | 1.60 (0.74, 3.44) |
| **Outcome: HPIV** | | | | | | | | | | | | |
| RSV | 1.82 (0.24, 13.81) | — | 0.79 (0.11, 5.94) | 1.25 (0.29, 5.28) | 1.15 (0.29, 4.61) | 1.30 (0.59, 2.89) | 2.80 (0.67, 11.68) | 1.70 (0.23, 12.53) | 0.69 (0.10, 4.96) | 1.09 (0.26, 4.53) | 1.99 (0.74, 5.37) | 1.20 (0.54, 2.66) |
| SC2 | **3.63 (1.17, 11.32)** | — | 0.50 (0.08, 3.16) | 0.53 (0.14, 2.05) | 1.77 (0.75, 4.17) | 1.42 (0.69, 2.92) | 2.34 (0.74, 7.35) | 0.86 (0.12, 6.01) | 0.35 (0.05, 2.38) | 0.71 (0.26, 1.93) | 1.45 (0.66, 3.20) | 1.14 (0.59, 2.20) |
| FLU | — | — | 2.66 (0.63, 11.26) | 1.06 (0.26, 4.30) | 0.88 (0.22, 3.53) | 0.16 (0.02, 1.22) | — | — | 1.79 (0.42, 7.56) | 0.79 (0.20, 3.07) | 1.01 (0.33, 3.10) | 0.26 (0.06, 1.07) |
| ADV | 2.03 (0.27, 15.11) | — | 1.28 (0.31, 5.33) | **2.62 (1.11, 6.17)** | **3.51 (1.57, 7.86)** | 1.43 (0.59, 3.46) | 1.98 (0.26, 15.15) | — | 1.26 (0.30, 5.26) | **2.67 (1.13, 6.31)** | **3.48 (1.55, 7.81)** | 1.42 (0.58, 3.47) |
| EV | — | — | 0.82 (0.21, 3.28) | 1.75 (0.78, 3.96) | 0.82 (0.22, 3.04) | 0.97 (0.31, 3.04) | — | — | 0.83 (0.21, 3.27) | 1.71 (0.76, 3.89) | 0.81 (0.22, 3.00) | 1.00 (0.32, 3.15) |
| RV | — | — | 1.58 (0.91, 2.76) | 1.18 (0.72, 1.95) | 1.15 (0.65, 2.02) | 0.78 (0.49, 1.24) | — | — | 1.60 (0.92, 2.78) | 1.22 (0.74, 2.00) | 1.15 (0.65, 2.04) | 0.78 (0.49, 1.24) |
| HCOV | — | — | 1.82 (0.77, 4.27) | 0.57 (0.19, 1.74) | 0.78 (0.30, 1.98) | 1.44 (0.84, 2.48) | — | — | 1.79 (0.76, 4.21) | 0.58 (0.19, 1.75) | 0.81 (0.32, 2.06) | 1.49 (0.86, 2.56) |
| HMPV | — | — | — | 2.29 (0.81, 6.51) | 0.55 (0.08, 3.95) | 1.87 (0.88, 3.96) | — | — | — | 2.31 (0.81, 6.58) | 0.54 (0.07, 4.06) | 1.93 (0.91, 4.12) |

Columns within each block give exposure windows in days. Cell shading reflects the hazard ratio: navy below 1, white at 1, red above 1, clamped at 4. Bold indicates $p < 0.05$ ; em dashes denote cells where the hazard ratio was not estimable. Blank cells were not fitted in that model.

#### Supplemental Table 5: Age-specific HRs and 95% CI estimates for children 12-17 years old.

Adjusted hazard ratios (aHRs) and 95% confidence intervals are from Cox proportional hazards models with time-varying indicator covariates for each pathogen exposure window (1–7, 8–14, 15–30, 31–60, 61–90, and 91–180 days after first positive swab) and an interaction term between exposure and age category, where the reference category is children aged 12–17 years old. Each row represents a distinct exposure pathogen and each outcome pathogen is shown in a separate block. The left panel includes symptomatic infections only; the right panel additionally includes asymptomatic infections for SC2, FLU, and RSV. Cell shading reflects the magnitude of the aHR: navy indicates values below 1, white indicates 1, and red indicates values above 1 (clamped at 4). Bold indicates $p<0.05$;em dashes denote cells with complete separation of the exposure and outcome data, where the aHR was not estimable. Abbreviations: SC2, SARS-CoV-2; FLU, influenza; RSV, respiratory syncytial virus; ADV, adenovirus; EV, enterovirus; RV, rhinovirus; HCOV, seasonal coronavirus; HMPV, human metapneumovirus; HPIV, human parainfluenza virus.

#### Age-specific hazard ratios: 12-17 yr (1 of 2)

Hazard of symptomatic infection with a second pathogen

| Exposure | Symptomatic infections only | | | | | | Asymptomatic infections included (SC2, FLU, RSV) | | | | | |
|---|---|---|---|---|---|---|---|---|---|---|---|---|
| Exposure | 1–7 | 8–14 | 15–30 | 31–60 | 61–90 | 91–180 | 1–7 | 8–14 | 15–30 | 31–60 | 61–90 | 91–180 |
| **Outcome: RSV** | | | | | | | | | | | | |
| SC2 | 3.19 (0.16, 61.73) | — | 1.43 (0.14, 14.15) | — | 2.33 (0.67, 8.08) | 1.50 (0.53, 4.27) | 3.53 (0.82, 15.08) | 2.06 (0.29, 14.79) | 2.09 (0.49, 8.91) | 0.47 (0.06, 3.50) | 1.65 (0.58, 4.70) | 1.83 (0.92, 3.67) |
| FLU | — | — | 1.38 (0.21, 9.09) | — | 3.11 (0.47, 20.47) | **9.48 (2.36, 38.07)** | — | — | 2.21 (0.58, 8.49) | — | 2.03 (0.32, 12.91) | **4.63 (1.26, 17.05)** |
| ADV | — | — | **28.22 (8.38, 95.00)** | — | — | **4.90 (1.52, 15.80)** | — | — | **16.21 (4.32, 60.85)** | 2.87 (0.29, 28.52) | — | 1.56 (0.09, 25.96) |
| EV | — | — | — | 1.37 (0.17, 11.10) | **6.06 (1.10, 33.49)** | 0.82 (0.10, 7.04) | — | — | — | 0.70 (0.09, 5.22) | 2.66 (0.68, 10.44) | 1.38 (0.32, 5.87) |
| RV | 0.52 (0.03, 10.47) | — | 1.02 (0.33, 3.14) | 0.92 (0.27, 3.05) | 0.25 (0.06, 1.10) | 0.88 (0.36, 2.13) | 1.24 (0.26, 5.86) | — | 0.91 (0.30, 2.76) | 1.16 (0.47, 2.87) | 0.97 (0.37, 2.59) | 0.98 (0.43, 2.25) |
| HCOV | — | — | 3.21 (0.34, 30.68) | — | **7.03 (1.55, 31.90)** | **0.11 (0.03, 0.45)** | — | — | 3.58 (0.44, 29.41) | **7.77 (1.47, 41.10)** | **7.52 (1.50, 37.63)** | 2.32 (0.30, 17.66) |
| HMPV | — | — | — | — | 16.25 (0.61, 432.29) | — | — | — | — | — | 4.02 (0.30, 54.27) | — |
| HPIV | — | — | — | **5.52 (1.45, 20.99)** | — | 1.27 (0.30, 5.48) | — | — | — | 2.68 (0.54, 13.36) | — | 1.03 (0.22, 4.82) |
| **Outcome: SC2** | | | | | | | | | | | | |
| RSV | — | — | 1.60 (0.22, 11.82) | 1.90 (0.47, 7.75) | 2.43 (0.60, 9.84) | 0.65 (0.08, 5.52) | — | 2.08 (0.29, 15.23) | 0.91 (0.12, 6.66) | 1.62 (0.53, 4.92) | **3.55 (1.51, 8.37)** | 1.54 (0.42, 5.69) |
| FLU | — | — | 1.73 (0.42, 7.16) | 1.75 (0.66, 4.64) | 0.56 (0.08, 4.03) | **4.14 (2.00, 8.58)** | — | — | 1.53 (0.49, 4.79) | **2.16 (1.02, 4.56)** | 0.77 (0.19, 3.15) | **3.49 (1.81, 6.72)** |
| ADV | — | — | — | — | 1.42 (0.27, 7.52) | 1.11 (0.29, 4.26) | — | — | — | — | 1.09 (0.22, 5.46) | 0.79 (0.19, 3.23) |
| EV | — | — | 1.49 (0.23, 9.50) | 0.95 (0.24, 3.76) | 1.62 (0.51, 5.14) | 1.10 (0.46, 2.65) | — | 2.16 (0.34, 13.70) | — | 1.09 (0.34, 3.53) | 1.57 (0.58, 4.22) | 1.01 (0.46, 2.25) |
| RV | 0.60 (0.15, 2.44) | 0.61 (0.15, 2.51) | **0.28 (0.09, 0.87)** | 1.03 (0.57, 1.86) | 1.16 (0.63, 2.12) | 1.25 (0.83, 1.89) | 0.42 (0.10, 1.69) | 0.50 (0.16, 1.52) | **0.15 (0.04, 0.50)** | 0.93 (0.53, 1.65) | 1.10 (0.66, 1.81) | 1.12 (0.78, 1.61) |
| HCOV | — | — | 0.44 (0.06, 3.22) | 0.87 (0.28, 2.74) | 0.40 (0.07, 2.46) | 0.83 (0.37, 1.88) | — | — | 0.67 (0.17, 2.67) | 0.89 (0.32, 2.43) | 0.59 (0.15, 2.30) | 1.07 (0.59, 1.95) |
| HMPV | — | — | 1.62 (0.21, 12.53) | 1.07 (0.14, 8.20) | — | 0.32 (0.04, 2.56) | — | — | 2.63 (0.62, 11.17) | 1.69 (0.41, 6.95) | — | 0.79 (0.23, 2.64) |
| HPIV | — | — | — | 0.80 (0.11, 5.77) | 2.35 (0.85, 6.50) | 1.72 (0.62, 4.72) | — | — | — | 0.65 (0.09, 4.65) | 1.86 (0.70, 4.91) | 0.76 (0.21, 2.73) |
| **Outcome: FLU** | | | | | | | | | | | | |
| RSV | — | — | 2.55 (0.72, 9.03) | — | — | 4.60 (0.49, 43.47) | — | — | 1.55 (0.45, 5.27) | 2.32 (0.77, 6.99) | — | 2.33 (0.29, 18.74) |
| SC2 | — | — | 2.94 (0.68, 12.74) | 1.52 (0.40, 5.69) | **0.07 (0.01, 0.43)** | 1.61 (0.69, 3.77) | — | 2.04 (0.31, 13.61) | 1.95 (0.48, 7.93) | 1.58 (0.54, 4.63) | 0.51 (0.05, 4.82) | **1.88 (1.05, 3.34)** |
| ADV | **18.26 (3.78, 88.11)** | — | **13.07 (2.90, 59.00)** | — | — | — | **13.07 (2.79, 61.30)** | **22.34 (2.01, 248.62)** | **11.47 (2.57, 51.25)** | — | — | — |
| EV | — | — | 2.84 (0.58, 13.88) | — | 0.67 (0.09, 4.75) | 1.77 (0.59, 5.37) | — | — | 2.63 (0.54, 12.91) | — | 0.45 (0.06, 3.14) | 1.28 (0.42, 3.92) |
| RV | 0.67 (0.21, 2.15) | 0.88 (0.12, 6.49) | 1.46 (0.57, 3.74) | 1.42 (0.69, 2.93) | 1.44 (0.69, 2.98) | 1.25 (0.64, 2.43) | 0.55 (0.16, 1.82) | 1.44 (0.36, 5.79) | 1.23 (0.49, 3.11) | 1.37 (0.67, 2.82) | 1.70 (0.91, 3.19) | 1.20 (0.67, 2.14) |
| HCOV | — | — | 2.35 (0.31, 17.89) | — | — | 3.31 (0.41, 27.07) | **5.59 (1.65, 18.99)** | — | 1.72 (0.22, 13.57) | 2.06 (0.45, 9.46) | **0.04 (0.01, 0.20)** | **0.07 (0.02, 0.31)** |
| HMPV | — | — | — | — | — | — | — | — | — | — | — | — |
| HPIV | — | — | 2.42 (0.29, 20.20) | — | 2.72 (0.34, 21.86) | 1.34 (0.20, 9.12) | — | — | 1.80 (0.22, 14.87) | — | 3.43 (0.95, 12.33) | — |
| **Outcome: ADV** | | | | | | | | | | | | |
| RSV | — | — | — | 7.64 (0.82, 71.39) | **9.94 (1.22, 80.84)** | **8.67 (2.08, 36.05)** | — | — | — | 4.87 (0.56, 42.28) | 7.78 (0.92, 65.68) | **6.46 (1.49, 27.95)** |
| SC2 | — | — | — | — | 1.50 (0.19, 11.75) | 0.67 (0.08, 5.44) | — | — | — | 1.08 (0.14, 8.21) | 1.00 (0.13, 7.76) | 0.91 (0.21, 4.02) |
| FLU | — | — | 7.44 (0.93, 59.52) | — | **9.27 (1.54, 55.74)** | 1.80 (0.26, 12.65) | **10.31 (1.43, 74.17)** | — | 5.18 (0.55, 48.83) | 0.54 (0.09, 3.29) | **6.13 (1.01, 37.35)** | 1.56 (0.24, 10.19) |
| EV | — | — | — | — | — | — | — | — | — | — | — | — |
| RV | — | — | 1.74 (0.49, 6.14) | 0.49 (0.09, 2.64) | 0.74 (0.18, 3.07) | 0.36 (0.13, 1.04) | — | — | 1.69 (0.50, 5.75) | 0.51 (0.10, 2.59) | 0.81 (0.20, 3.36) | 0.39 (0.15, 1.04) |
| HCOV | — | — | 2.20 (0.21, 22.94) | 1.64 (0.16, 16.25) | **0.10 (0.02, 0.59)** | 1.58 (0.34, 7.27) | — | — | 2.70 (0.29, 24.67) | 2.04 (0.24, 17.51) | **0.11 (0.02, 0.58)** | 1.43 (0.32, 6.45) |
| HMPV | — | — | — | — | — | 2.46 (0.29, 20.89) | — | — | — | — | — | 2.65 (0.32, 21.75) |
| HPIV | — | — | **9.32 (1.53, 56.82)** | — | — | **4.60 (1.36, 15.56)** | — | — | **9.32 (1.47, 59.20)** | — | — | **4.24 (1.20, 15.04)** |

Columns within each block give exposure windows in days. Cell shading reflects the hazard ratio: navy below 1, white at 1, red above 1, clamped at 4. Bold indicates $p < 0.05$ ; em dashes denote cells where the hazard ratio was not estimable. Blank cells were not fitted in that model.

#### Age-specific hazard ratios: 12-17 yr (2 of 2)

Hazard of symptomatic infection with a second pathogen

| | Symptomatic infections only | | | | | | Asymptomatic infections included (SC2, FLU, RSV) | | | | | |
|---|---|---|---|---|---|---|---|---|---|---|---|---|
| **Exposure** | **1–7** | **8–14** | **15–30** | **31–60** | **61–90** | **91–180** | **1–7** | **8–14** | **15–30** | **31–60** | **61–90** | **91–180** |
| **Outcome: EV** | | | | | | | | | | | | |
| RSV | — | — | — | 2.05 (0.30, 14.23) | — | 1.26 (0.16, 10.17) | — | — | — | 1.27 (0.18, 9.10) | — | 1.64 (0.35, 7.74) |
| SC2 | — | 4.00 (0.98, 16.33) | 1.65 (0.43, 6.36) | 1.17 (0.36, 3.88) | **0.12 (0.04, 0.36)** | 1.02 (0.40, 2.65) | 1.32 (0.18, 9.60) | **4.28 (1.31, 13.93)** | 1.86 (0.61, 5.72) | 0.90 (0.28, 2.88) | 0.41 (0.07, 2.34) | 1.43 (0.70, 2.89) |
| FLU | — | — | — | — | 3.20 (0.75, 13.67) | 1.28 (0.32, 5.12) | — | — | — | — | 2.40 (0.56, 10.24) | 1.56 (0.51, 4.75) |
| ADV | — | — | — | — | — | 1.04 (0.12, 8.87) | — | — | — | — | — | 1.01 (0.12, 8.57) |
| RV | 0.45 (0.06, 3.29) | — | 0.26 (0.04, 1.81) | 0.93 (0.39, 2.20) | 0.66 (0.22, 1.99) | 0.99 (0.56, 1.75) | 0.44 (0.06, 3.21) | — | 0.26 (0.04, 1.81) | 0.92 (0.39, 2.18) | 0.65 (0.22, 1.96) | 0.98 (0.55, 1.76) |
| HCOV | 2.38 (0.29, 19.21) | — | **0.04 (0.01, 0.29)** | 1.09 (0.14, 8.42) | 1.40 (0.19, 10.46) | 2.20 (0.85, 5.66) | 2.53 (0.32, 20.20) | — | **0.05 (0.01, 0.29)** | 1.12 (0.16, 7.90) | 1.37 (0.18, 10.32) | 2.08 (0.80, 5.43) |
| HMPV | **13.47 (1.98, 91.61)** | — | — | — | — | 3.12 (0.90, 10.82) | **13.58 (2.00, 92.41)** | — | — | — | — | 3.07 (0.88, 10.79) |
| HPIV | — | — | 3.94 (0.57, 27.19) | 1.81 (0.21, 15.40) | — | **0.07 (0.02, 0.29)** | — | — | 3.65 (0.50, 26.47) | 1.73 (0.20, 14.73) | — | **0.07 (0.02, 0.28)** |
| **Outcome: RV** | | | | | | | | | | | | |
| RSV | — | — | — | 1.94 (0.64, 5.89) | 1.85 (0.43, 8.03) | **2.63 (1.20, 5.77)** | — | — | — | 1.71 (0.56, 5.19) | 2.40 (0.73, 7.90) | **2.85 (1.35, 6.01)** |
| SC2 | — | — | 1.31 (0.55, 3.15) | 1.26 (0.66, 2.44) | 1.32 (0.64, 2.74) | 0.84 (0.40, 1.77) | — | 0.37 (0.05, 2.68) | 1.42 (0.71, 2.85) | 0.93 (0.49, 1.76) | 0.90 (0.46, 1.76) | 1.10 (0.66, 1.85) |
| FLU | — | — | 0.86 (0.14, 5.48) | 1.33 (0.48, 3.69) | 0.77 (0.20, 3.05) | 1.43 (0.67, 3.04) | — | — | 1.34 (0.39, 4.60) | 0.83 (0.21, 3.23) | 0.94 (0.32, 2.79) | 1.66 (0.80, 3.46) |
| ADV | — | — | 1.26 (0.28, 5.69) | 0.83 (0.09, 7.42) | — | 0.10 (0.00, 12.95) | — | — | 1.21 (0.29, 5.10) | 0.82 (0.09, 7.45) | — | 0.09 (0.00, 11.73) |
| EV | — | — | 0.99 (0.13, 7.31) | — | 1.78 (0.44, 7.18) | 1.01 (0.28, 3.57) | — | — | 1.03 (0.14, 7.51) | — | 1.78 (0.42, 7.61) | 0.96 (0.27, 3.41) |
| HCOV | — | — | 0.32 (0.04, 2.22) | 1.57 (0.74, 3.32) | 0.47 (0.11, 1.96) | **2.22 (1.37, 3.60)** | — | — | 0.31 (0.04, 2.17) | 1.55 (0.74, 3.28) | 0.46 (0.11, 1.93) | **2.25 (1.39, 3.65)** |
| HMPV | — | — | 1.93 (0.43, 8.67) | 1.74 (0.49, 6.22) | 1.13 (0.31, 4.18) | 2.15 (0.78, 5.92) | — | — | 1.97 (0.44, 8.73) | 1.62 (0.45, 5.90) | 1.10 (0.31, 3.93) | 2.15 (0.77, 5.97) |
| HPIV | — | — | 0.97 (0.12, 7.55) | 2.10 (0.63, 6.94) | — | **2.45 (1.26, 4.79)** | — | — | 0.96 (0.12, 7.47) | 2.09 (0.63, 6.87) | — | **2.37 (1.21, 4.66)** |
| **Outcome: HCOV** | | | | | | | | | | | | |
| RSV | — | — | 1.16 (0.16, 8.16) | 1.18 (0.32, 4.43) | 2.15 (0.71, 6.52) | 1.52 (0.47, 4.92) | — | — | 0.93 (0.13, 6.53) | 1.28 (0.42, 3.90) | 1.51 (0.52, 4.41) | 1.17 (0.39, 3.45) |
| SC2 | — | — | 0.90 (0.22, 3.75) | 0.26 (0.03, 2.02) | **0.05 (0.01, 0.15)** | 1.10 (0.53, 2.27) | — | 0.65 (0.09, 4.64) | 1.45 (0.60, 3.49) | 0.54 (0.17, 1.76) | 0.23 (0.04, 1.44) | 0.88 (0.43, 1.78) |
| FLU | — | — | 0.66 (0.09, 4.78) | 1.09 (0.37, 3.23) | 1.66 (0.78, 3.53) | 0.63 (0.22, 1.83) | 1.47 (0.15, 14.72) | — | **0.14 (0.04, 0.57)** | 1.08 (0.40, 2.93) | 1.64 (0.81, 3.33) | 0.55 (0.19, 1.60) |
| ADV | — | — | 1.94 (0.15, 24.49) | 3.37 (0.58, 19.77) | 3.26 (0.84, 12.66) | **4.37 (1.68, 11.36)** | — | — | 2.82 (0.23, 34.13) | 4.10 (0.68, 24.66) | 3.39 (0.89, 12.95) | **4.63 (1.77, 12.09)** |
| EV | — | — | 1.47 (0.19, 11.44) | 2.10 (0.77, 5.73) | 2.16 (0.60, 7.76) | 1.48 (0.65, 3.34) | — | — | 1.38 (0.17, 10.88) | 1.94 (0.69, 5.46) | 2.21 (0.62, 7.81) | 1.50 (0.66, 3.43) |
| RV | 0.38 (0.05, 2.78) | — | 1.27 (0.58, 2.78) | **2.26 (1.35, 3.79)** | 1.05 (0.50, 2.21) | 1.13 (0.72, 1.78) | 0.37 (0.05, 2.73) | — | 1.27 (0.58, 2.79) | **2.24 (1.33, 3.77)** | 1.03 (0.48, 2.23) | 1.12 (0.72, 1.76) |
| HMPV | — | — | 2.55 (0.50, 13.15) | 1.95 (0.38, 9.86) | 4.45 (0.88, 22.45) | — | — | — | 2.27 (0.40, 12.78) | 1.79 (0.33, 9.67) | 4.47 (0.89, 22.53) | — |
| HPIV | — | — | 2.89 (0.61, 13.63) | 1.82 (0.55, 5.98) | — | 0.38 (0.05, 2.74) | — | — | 2.99 (0.68, 13.14) | 2.00 (0.64, 6.28) | — | 0.38 (0.05, 2.72) |
| **Outcome: HMPV** | | | | | | | | | | | | |
| RSV | — | — | **4.16 (1.09, 15.87)** | — | 1.77 (0.22, 14.33) | 1.88 (0.37, 9.49) | — | — | 3.47 (0.84, 14.34) | — | 1.37 (0.17, 10.90) | 1.36 (0.28, 6.70) |
| SC2 | — | — | **4.35 (1.51, 12.54)** | **5.53 (2.26, 13.51)** | 1.24 (0.17, 9.21) | 1.60 (0.50, 5.17) | — | — | **3.21 (1.08, 9.54)** | **4.22 (1.79, 9.95)** | 0.77 (0.10, 5.60) | 1.14 (0.37, 3.50) |
| FLU | — | — | — | 1.90 (0.25, 14.70) | 1.00 (0.14, 7.07) | **3.19 (1.40, 7.26)** | — | — | — | 1.64 (0.22, 12.14) | 0.70 (0.10, 5.18) | **2.69 (1.14, 6.32)** |
| ADV | — | — | — | — | — | 1.14 (0.25, 5.17) | — | — | — | — | — | 1.45 (0.34, 6.15) |
| EV | — | — | 1.98 (0.17, 23.17) | 1.65 (0.23, 11.62) | 2.42 (0.34, 17.29) | 0.75 (0.09, 6.08) | — | — | 1.98 (0.19, 20.61) | 1.67 (0.23, 12.22) | 2.30 (0.32, 16.59) | 0.75 (0.09, 6.07) |
| RV | — | — | 1.21 (0.41, 3.62) | 1.00 (0.32, 3.15) | 1.53 (0.58, 4.04) | 0.97 (0.45, 2.09) | — | — | 1.23 (0.41, 3.68) | 1.08 (0.34, 3.37) | 1.57 (0.57, 4.35) | 1.01 (0.48, 2.13) |
| HCOV | — | — | **3.51 (1.15, 10.74)** | 0.52 (0.06, 4.70) | 1.66 (0.44, 6.32) | 1.57 (0.21, 11.80) | — | — | **3.62 (1.15, 11.37)** | 0.50 (0.05, 4.67) | 1.64 (0.44, 6.15) | 1.52 (0.20, 11.50) |
| HPIV | — | — | **8.93 (2.22, 35.94)** | 2.43 (0.45, 13.02) | — | **3.80 (1.08, 13.31)** | — | — | **8.99 (2.20, 36.73)** | 2.69 (0.50, 14.39) | — | **4.21 (1.31, 13.50)** |
| **Outcome: HPIV** | | | | | | | | | | | | |
| RSV | — | — | 6.55 (0.75, 57.24) | — | 3.52 (0.43, 28.67) | 1.38 (0.35, 5.46) | — | — | 4.81 (0.58, 40.19) | — | 3.24 (0.42, 25.25) | 2.46 (0.74, 8.21) |
| SC2 | — | — | — | **3.43 (1.34, 8.78)** | 1.67 (0.40, 7.01) | 0.82 (0.23, 2.98) | — | 1.90 (0.20, 17.54) | **0.17 (0.05, 0.66)** | **2.87 (1.17, 7.06)** | 1.09 (0.26, 4.62) | 0.59 (0.19, 1.87) |
| FLU | — | — | 3.53 (0.47, 26.44) | — | 1.45 (0.19, 10.91) | 1.67 (0.46, 6.01) | — | — | 3.17 (0.47, 21.36) | — | 1.18 (0.16, 8.89) | 1.39 (0.38, 5.03) |
| ADV | — | — | **6.34 (1.64, 24.53)** | 8.05 (0.94, 68.76) | — | 2.78 (0.33, 23.71) | — | — | **6.65 (2.14, 20.67)** | **8.50 (1.06, 67.97)** | — | 2.46 (0.26, 22.80) |
| EV | — | — | — | — | — | 0.63 (0.08, 4.84) | — | — | — | — | — | 0.63 (0.08, 4.84) |
| RV | 0.70 (0.13, 3.73) | — | **0.03 (0.01, 0.11)** | 1.44 (0.68, 3.05) | 0.46 (0.05, 4.23) | 1.38 (0.66, 2.91) | 0.66 (0.13, 3.25) | — | **0.04 (0.01, 0.11)** | 1.46 (0.70, 3.08) | 0.46 (0.05, 4.37) | 1.37 (0.65, 2.89) |
| HCOV | — | — | — | 2.04 (0.60, 6.97) | **0.13 (0.03, 0.51)** | 0.82 (0.19, 3.61) | — | — | — | 1.92 (0.57, 6.52) | **0.11 (0.03, 0.42)** | 0.78 (0.19, 3.14) |
| HMPV | — | — | 2.75 (0.89, 8.48) | — | — | 1.40 (0.18, 10.84) | — | — | 2.61 (0.99, 6.86) | — | — | 1.40 (0.19, 10.56) |

Columns within each block give exposure windows in days. Cell shading reflects the hazard ratio: navy below 1, white at 1, red above 1, clamped at 4. Bold indicates $p < 0.05$ ; em dashes denote cells where the hazard ratio was not estimable. Blank cells were not fitted in that model.

#### Supplemental Table 6: Age-specific HRs and 95% CI estimates for adults 18–50 years old.

Adjusted hazard ratios (aHRs) and 95% confidence intervals are from Cox proportional hazards models with time-varying indicator covariates for each pathogen exposure window (1–7, 8–14, 15–30, 31–60, 61–90, and 91–180 days after first positive swab) and an interaction term between exposure and age category, where the reference category is adults aged 18–50 years old. Note that the age range is inclusive of 50 year olds because 49 year olds were allowed to enroll in the study. Each row represents a distinct exposure pathogen and each outcome pathogen is shown in a separate block. The left panel includes symptomatic infections only; the right panel additionally includes asymptomatic infections for SC2, FLU, and RSV. Cell shading reflects the magnitude of the aHR: navy indicates values below 1, white indicates 1, and red indicates values above 1 (clamped at 4). Bold indicates p<0.05; em dashes denote cells with complete separation of the exposure and outcome data, where the aHR was not estimable. Abbreviations: SC2, SARS-CoV-2; FLU, influenza; RSV, respiratory syncytial virus; ADV, adenovirus; EV, enterovirus; RV, rhinovirus; HCOV, seasonal coronavirus; HMPV, human metapneumovirus; HPIV, human parainfluenza virus.

#### Age-specific hazard ratios: 18-49 yr (1 of 2)

Hazard of symptomatic infection with a second pathogen

| | Symptomatic infections only | | | | | | Asymptomatic infections included (SC2, FLU, RSV) | | | | | |
|---|---|---|---|---|---|---|---|---|---|---|---|---|
| **Exposure** | **1–7** | **8–14** | **15–30** | **31–60** | **61–90** | **91–180** | **1–7** | **8–14** | **15–30** | **31–60** | **61–90** | **91–180** |
| **Outcome: RSV** | | | | | | | | | | | | |
| SC2 | — | 1.87 (0.44, 7.89) | 0.44 (0.07, 2.92) | 1.02 (0.37, 2.81) | 0.30 (0.06, 1.66) | 0.93 (0.36, 2.40) | 0.55 (0.08, 4.06) | 1.83 (0.59, 5.69) | 0.82 (0.27, 2.53) | 1.60 (0.74, 3.43) | **0.12 (0.02, 0.66)** | 1.01 (0.54, 1.92) |
| FLU | — | — | 3.40 (0.84, 13.69) | — | 3.79 (0.50, 28.94) | **0.07 (0.01, 0.34)** | — | — | **6.48 (2.82, 14.87)** | 1.63 (0.40, 6.65) | 2.43 (0.34, 17.25) | **0.06 (0.02, 0.19)** |
| ADV | — | — | — | 2.59 (0.60, 11.08) | **3.72 (1.10, 12.58)** | **0.31 (0.14, 0.71)** | — | — | — | 1.97 (0.49, 7.86) | **3.68 (1.22, 11.09)** | 0.67 (0.21, 2.08) |
| EV | **4.69 (1.15, 19.14)** | — | 0.66 (0.08, 5.67) | 1.44 (0.45, 4.63) | 0.65 (0.15, 2.89) | 1.62 (0.51, 5.10) | 3.48 (0.79, 15.25) | 6.14 (0.81, 46.31) | — | 1.56 (0.54, 4.48) | 1.11 (0.39, 3.16) | 1.82 (0.75, 4.39) |
| RV | 0.32 (0.08, 1.29) | — | 1.16 (0.48, 2.78) | 0.85 (0.40, 1.81) | **2.26 (1.18, 4.32)** | 1.05 (0.47, 2.36) | 0.29 (0.07, 1.22) | — | 1.58 (0.79, 3.19) | 0.74 (0.35, 1.55) | 1.68 (0.90, 3.13) | 0.97 (0.49, 1.92) |
| HCOV | — | — | — | **4.95 (1.28, 19.16)** | 2.73 (0.37, 20.24) | 2.24 (0.60, 8.34) | — | 2.03 (0.25, 16.28) | **0.09 (0.02, 0.40)** | 3.26 (0.74, 14.32) | 3.67 (0.87, 15.51) | 1.44 (0.39, 5.28) |
| HMPV | — | — | — | **7.86 (1.70, 36.22)** | — | — | — | — | — | **13.58 (4.67, 39.52)** | — | — |
| HPIV | 3.97 (0.55, 28.89) | — | 2.05 (0.31, 13.45) | 2.17 (0.46, 10.30) | **5.15 (1.61, 16.55)** | 0.80 (0.11, 5.75) | 2.97 (0.42, 20.99) | — | 1.37 (0.21, 9.14) | 1.55 (0.37, 6.47) | **4.94 (1.73, 14.12)** | 0.96 (0.23, 3.95) |
| **Outcome: SC2** | | | | | | | | | | | | |
| RSV | — | 1.09 (0.16, 7.21) | 1.27 (0.41, 3.98) | 1.42 (0.63, 3.21) | 0.52 (0.13, 2.12) | 0.71 (0.30, 1.71) | 0.72 (0.10, 4.99) | 1.40 (0.35, 5.67) | 0.88 (0.29, 2.69) | 1.62 (0.77, 3.42) | 0.33 (0.08, 1.36) | 0.81 (0.41, 1.63) |
| FLU | — | — | 1.02 (0.15, 7.12) | 1.69 (0.67, 4.27) | 0.37 (0.05, 2.66) | 1.97 (0.96, 4.05) | 0.82 (0.11, 5.95) | **0.03 (0.00, 0.15)** | 0.65 (0.09, 4.67) | 1.48 (0.68, 3.23) | 0.22 (0.03, 1.60) | **2.19 (1.23, 3.91)** |
| ADV | 1.27 (0.17, 9.32) | 1.30 (0.19, 9.00) | 1.57 (0.58, 4.26) | 0.41 (0.14, 1.17) | 0.80 (0.30, 2.15) | 1.14 (0.63, 2.07) | 1.07 (0.15, 7.79) | 2.00 (0.50, 8.07) | 1.45 (0.56, 3.79) | 0.52 (0.19, 1.42) | 0.74 (0.30, 1.83) | 1.31 (0.77, 2.22) |
| EV | 1.28 (0.16, 10.32) | — | 2.11 (0.96, 4.67) | 0.71 (0.29, 1.77) | 1.53 (0.75, 3.11) | 1.26 (0.75, 2.11) | 1.16 (0.15, 9.06) | — | 1.82 (0.82, 4.03) | 0.87 (0.40, 1.90) | 1.52 (0.78, 2.95) | 1.08 (0.64, 1.82) |
| RV | **0.12 (0.01, 0.93)** | 0.29 (0.07, 1.13) | 0.57 (0.31, 1.05) | 1.14 (0.75, 1.72) | 1.26 (0.84, 1.89) | 1.09 (0.83, 1.43) | **0.11 (0.01, 0.81)** | 0.75 (0.34, 1.68) | 0.59 (0.33, 1.03) | 1.17 (0.80, 1.70) | 1.25 (0.87, 1.79) | 1.10 (0.85, 1.41) |
| HCOV | 0.32 (0.05, 2.33) | 0.34 (0.05, 2.34) | 0.94 (0.42, 2.11) | 1.29 (0.71, 2.34) | 0.97 (0.41, 2.30) | 1.22 (0.81, 1.83) | 0.57 (0.14, 2.32) | 0.29 (0.04, 1.96) | 0.99 (0.47, 2.09) | 1.34 (0.77, 2.32) | 0.96 (0.44, 2.11) | 1.17 (0.80, 1.72) |
| HMPV | — | — | 1.61 (0.40, 6.50) | 1.10 (0.27, 4.46) | 1.17 (0.29, 4.80) | 0.60 (0.22, 1.64) | — | — | 2.18 (0.70, 6.80) | 0.98 (0.24, 4.00) | 1.56 (0.48, 5.03) | 0.68 (0.28, 1.65) |
| HPIV | 0.98 (0.15, 6.21) | — | 1.75 (0.53, 5.73) | 1.43 (0.64, 3.20) | 0.68 (0.22, 2.09) | 1.52 (0.89, 2.60) | 0.87 (0.13, 5.58) | — | 1.53 (0.46, 5.04) | 1.64 (0.82, 3.31) | 0.98 (0.41, 2.35) | 1.53 (0.94, 2.48) |
| **Outcome: FLU** | | | | | | | | | | | | |
| RSV | — | — | — | 0.53 (0.07, 4.01) | — | — | — | 1.18 (0.17, 8.21) | 0.58 (0.08, 4.29) | — | — | — |
| SC2 | 1.07 (0.15, 7.61) | — | 2.32 (0.80, 6.67) | 1.69 (0.68, 4.20) | 0.69 (0.18, 2.68) | 0.95 (0.38, 2.40) | 0.75 (0.11, 5.35) | 1.45 (0.35, 5.96) | 1.54 (0.54, 4.40) | 1.43 (0.62, 3.25) | 1.20 (0.43, 3.33) | 0.65 (0.27, 1.58) |
| ADV | — | — | — | 3.06 (0.86, 10.83) | 1.87 (0.23, 15.07) | 1.08 (0.20, 5.73) | — | — | — | 2.64 (0.76, 9.21) | 2.53 (0.54, 11.78) | 1.64 (0.48, 5.66) |
| EV | — | — | **3.97 (1.23, 12.84)** | 2.02 (0.81, 5.00) | 0.38 (0.06, 2.20) | 1.09 (0.22, 5.50) | — | 1.26 (0.16, 9.65) | 2.95 (0.91, 9.56) | 1.73 (0.70, 4.26) | 0.26 (0.05, 1.19) | 1.43 (0.41, 5.02) |
| RV | — | 0.60 (0.08, 4.46) | 0.62 (0.22, 1.77) | 1.11 (0.59, 2.08) | 1.33 (0.57, 3.10) | 0.67 (0.30, 1.47) | — | **2.49 (1.04, 5.97)** | 0.79 (0.35, 1.79) | 1.07 (0.60, 1.89) | 1.59 (0.81, 3.11) | 0.48 (0.22, 1.03) |
| HCOV | — | — | — | — | 3.76 (0.83, 17.14) | 2.80 (0.79, 9.98) | 1.66 (0.23, 11.97) | — | — | — | 2.90 (0.68, 12.30) | **4.08 (1.60, 10.43)** |
| HMPV | — | — | — | — | — | 5.78 (0.97, 34.58) | — | — | — | — | — | 3.58 (0.55, 23.21) |
| HPIV | — | — | 1.94 (0.27, 14.00) | 1.11 (0.15, 8.30) | 3.89 (0.95, 15.89) | — | — | **6.90 (1.68, 28.42)** | 1.78 (0.29, 10.73) | 0.81 (0.10, 6.37) | 4.15 (0.95, 18.16) | — |
| **Outcome: ADV** | | | | | | | | | | | | |
| RSV | — | — | **4.44 (1.16, 16.97)** | **3.52 (1.06, 11.71)** | **0.14 (0.04, 0.56)** | 1.20 (0.29, 4.99) | — | — | 3.20 (0.75, 13.68) | 3.05 (0.93, 10.04) | **0.09 (0.02, 0.37)** | 0.98 (0.24, 4.07) |
| SC2 | **5.78 (2.09, 16.01)** | **5.28 (1.90, 14.65)** | **3.00 (1.33, 6.77)** | 0.32 (0.05, 1.90) | 0.59 (0.15, 2.38) | 0.67 (0.28, 1.63) | **4.88 (2.00, 11.89)** | **5.19 (2.09, 12.87)** | **2.57 (1.14, 5.80)** | 0.27 (0.05, 1.49) | 0.48 (0.12, 1.94) | 0.47 (0.18, 1.23) |
| FLU | — | — | — | 1.44 (0.21, 9.77) | — | 0.78 (0.11, 5.73) | — | — | — | 0.86 (0.10, 7.27) | — | 0.55 (0.07, 4.07) |
| EV | 6.71 (0.92, 48.96) | — | — | 1.33 (0.39, 4.53) | 0.62 (0.09, 4.49) | 1.09 (0.36, 3.32) | 6.75 (0.93, 49.16) | — | — | 1.35 (0.39, 4.64) | 0.63 (0.09, 4.53) | 1.10 (0.36, 3.39) |
| RV | — | — | 2.02 (0.87, 4.71) | 1.19 (0.56, 2.53) | 1.40 (0.68, 2.89) | 1.59 (0.91, 2.78) | — | — | 2.03 (0.87, 4.73) | 1.18 (0.56, 2.52) | 1.40 (0.68, 2.89) | 1.61 (0.92, 2.83) |
| HCOV | — | 1.56 (0.20, 12.13) | 1.43 (0.34, 6.04) | 1.43 (0.45, 4.55) | 1.57 (0.50, 4.94) | 1.04 (0.44, 2.50) | — | 1.63 (0.21, 12.44) | 1.46 (0.35, 6.15) | 1.44 (0.46, 4.58) | 1.57 (0.50, 4.95) | 1.05 (0.44, 2.53) |
| HMPV | — | — | — | — | 3.17 (0.81, 12.43) | 0.87 (0.12, 6.29) | — | — | — | — | 3.12 (0.80, 12.22) | 0.88 (0.12, 6.34) |
| HPIV | **8.67 (2.00, 37.55)** | — | **0.15 (0.03, 0.70)** | 0.96 (0.15, 6.12) | **4.74 (1.77, 12.68)** | 1.92 (0.68, 5.42) | **8.66 (2.00, 37.45)** | — | **0.16 (0.03, 0.73)** | 0.98 (0.14, 6.80) | **4.68 (1.73, 12.66)** | 2.00 (0.71, 5.63) |

Columns within each block give exposure windows in days. Cell shading reflects the hazard ratio: navy below 1, white at 1, red above 1, clamped at 4. Bold indicates $p < 0.05$ ; em dashes denote cells where the hazard ratio was not estimable. Blank cells were not fitted in that model.

#### Age-specific hazard ratios: 18-49 yr (2 of 2)

Hazard of symptomatic infection with a second pathogen

| Exposure | Symptomatic infections only | | | | | | Asymptomatic infections included (SC2, FLU, RSV) | | | | | |
|---|---|---|---|---|---|---|---|---|---|---|---|---|
| | 1–7 | 8–14 | 15–30 | 31–60 | 61–90 | 91–180 | 1–7 | 8–14 | 15–30 | 31–60 | 61–90 | 91–180 |
| **Outcome: EV** | | | | | | | | | | | | |
| RSV | — | — | 1.39 (0.19, 10.06) | 0.78 (0.11, 5.61) | 2.60 (0.81, 8.38) | 0.83 (0.20, 3.42) | — | — | 2.15 (0.53, 8.77) | 1.69 (0.53, 5.44) | 1.91 (0.60, 6.09) | 0.67 (0.16, 2.72) |
| SC2 | 2.57 (0.91, 7.26) | 1.65 (0.53, 5.16) | 0.96 (0.36, 2.57) | 0.83 (0.37, 1.86) | 0.83 (0.35, 2.00) | 1.22 (0.74, 2.03) | 1.75 (0.52, 5.83) | 2.37 (0.97, 5.75) | 0.80 (0.30, 2.13) | 0.70 (0.32, 1.57) | 1.00 (0.48, 2.08) | 1.19 (0.74, 1.91) |
| FLU | — | — | — | — | 1.12 (0.15, 8.09) | 1.56 (0.47, 5.23) | — | — | 1.53 (0.20, 11.57) | **0.04 (0.01, 0.25)** | 0.78 (0.10, 6.06) | 1.59 (0.56, 4.47) |
| ADV | 3.72 (0.85, 16.37) | — | **0.16 (0.04, 0.67)** | 0.63 (0.08, 4.88) | 0.83 (0.14, 4.99) | 1.09 (0.35, 3.44) | 3.81 (0.87, 16.69) | — | **0.15 (0.04, 0.65)** | 0.63 (0.08, 4.89) | 0.83 (0.14, 5.02) | 1.08 (0.34, 3.45) |
| RV | **2.47 (1.23, 4.96)** | — | 0.73 (0.28, 1.90) | 1.10 (0.59, 2.04) | 1.03 (0.50, 2.11) | 1.24 (0.81, 1.90) | **2.49 (1.24, 5.01)** | — | 0.73 (0.28, 1.90) | 1.10 (0.60, 2.04) | 1.02 (0.49, 2.11) | 1.24 (0.81, 1.90) |
| HCOV | — | — | 1.60 (0.50, 5.12) | **2.61 (1.24, 5.47)** | **0.28 (0.14, 0.56)** | 1.51 (0.76, 3.03) | — | — | 1.60 (0.49, 5.19) | **2.66 (1.26, 5.60)** | **0.29 (0.14, 0.57)** | 1.51 (0.76, 3.03) |
| HMPV | — | — | 2.52 (0.36, 17.78) | | — | 1.52 (0.49, 4.69) | — | — | 2.59 (0.37, 18.18) | | — | 1.54 (0.50, 4.75) |
| HPIV | — | — | 1.21 (0.18, 8.23) | 1.31 (0.33, 5.24) | 0.68 (0.11, 4.20) | 1.92 (0.98, 3.76) | — | — | 1.18 (0.17, 8.08) | 1.34 (0.33, 5.35) | 0.69 (0.11, 4.25) | 1.94 (0.99, 3.81) |
| **Outcome: RV** | | | | | | | | | | | | |
| RSV | — | 0.87 (0.12, 6.23) | 0.39 (0.06, 2.72) | 1.20 (0.52, 2.79) | 0.52 (0.08, 3.40) | 1.06 (0.54, 2.05) | — | 0.65 (0.09, 4.66) | 1.22 (0.45, 3.26) | 1.13 (0.50, 2.54) | 0.63 (0.15, 2.60) | 0.97 (0.52, 1.82) |
| SC2 | 1.50 (0.75, 2.98) | 0.90 (0.38, 2.18) | 1.06 (0.61, 1.84) | 0.66 (0.39, 1.12) | 1.19 (0.76, 1.84) | 1.07 (0.77, 1.49) | 0.96 (0.43, 2.15) | 1.24 (0.62, 2.46) | 1.04 (0.62, 1.75) | **0.57 (0.34, 0.96)** | 1.16 (0.77, 1.75) | 1.06 (0.77, 1.45) |
| FLU | — | — | — | 0.83 (0.26, 2.60) | 0.88 (0.21, 3.62) | 0.70 (0.29, 1.65) | — | — | — | 0.66 (0.22, 2.04) | 0.70 (0.17, 2.93) | 0.75 (0.36, 1.58) |
| ADV | 1.01 (0.14, 7.17) | — | 1.64 (0.53, 5.07) | **2.55 (1.23, 5.28)** | 2.11 (0.95, 4.68) | **2.07 (1.12, 3.83)** | 1.03 (0.15, 7.26) | — | 1.58 (0.51, 4.91) | **2.57 (1.24, 5.33)** | 2.09 (0.95, 4.62) | **2.07 (1.12, 3.83)** |
| EV | 2.63 (0.66, 10.55) | — | — | 1.64 (0.59, 4.54) | 1.05 (0.30, 3.63) | 1.05 (0.39, 2.84) | 2.59 (0.65, 10.43) | — | — | 1.65 (0.59, 4.60) | 1.05 (0.30, 3.62) | 1.06 (0.39, 2.86) |
| HCOV | 0.28 (0.04, 2.02) | — | 0.59 (0.22, 1.58) | 1.51 (0.93, 2.44) | 1.62 (0.95, 2.74) | 1.01 (0.63, 1.60) | 0.28 (0.04, 2.04) | — | 0.60 (0.22, 1.60) | 1.53 (0.95, 2.48) | 1.63 (0.96, 2.77) | 1.02 (0.64, 1.62) |
| HMPV | | — | 1.73 (0.59, 5.08) | 0.66 (0.16, 2.66) | 1.03 (0.33, 3.24) | 0.77 (0.29, 2.06) | | — | 1.75 (0.60, 5.10) | 0.66 (0.16, 2.69) | 1.04 (0.33, 3.28) | 0.77 (0.29, 2.06) |
| HPIV | 0.67 (0.10, 4.76) | — | 2.00 (0.91, 4.39) | 1.34 (0.62, 2.94) | 0.79 (0.28, 2.23) | **1.96 (1.23, 3.14)** | 0.66 (0.09, 4.68) | — | 2.00 (0.91, 4.39) | 1.35 (0.62, 2.95) | 0.79 (0.28, 2.24) | **1.97 (1.23, 3.15)** |
| **Outcome: HCOV** | | | | | | | | | | | | |
| RSV | — | — | 2.12 (0.81, 5.53) | 1.62 (0.78, 3.37) | 1.83 (0.98, 3.40) | 1.28 (0.66, 2.47) | — | — | 1.23 (0.41, 3.67) | 1.65 (0.85, 3.19) | **2.09 (1.22, 3.56)** | 1.18 (0.64, 2.19) |
| SC2 | 1.00 (0.32, 3.15) | 0.96 (0.31, 3.00) | 1.51 (0.85, 2.70) | 1.00 (0.59, 1.70) | 0.84 (0.45, 1.55) | 1.09 (0.73, 1.62) | 0.85 (0.27, 2.65) | 0.80 (0.25, 2.49) | 1.31 (0.74, 2.34) | 0.92 (0.55, 1.53) | 0.72 (0.40, 1.31) | 1.13 (0.79, 1.63) |
| FLU | 1.65 (0.23, 11.89) | **0.04 (0.00, 0.36)** | 0.49 (0.07, 3.46) | 1.40 (0.70, 2.82) | 0.99 (0.41, 2.43) | 0.96 (0.39, 2.35) | 1.12 (0.15, 8.59) | **0.08 (0.02, 0.35)** | 1.20 (0.39, 3.71) | 1.68 (0.94, 2.99) | 0.74 (0.30, 1.82) | 1.18 (0.57, 2.42) |
| ADV | — | — | 1.46 (0.35, 6.07) | 1.66 (0.60, 4.57) | 1.01 (0.28, 3.65) | 1.21 (0.52, 2.83) | — | — | 1.50 (0.36, 6.18) | 1.73 (0.65, 4.65) | 1.02 (0.29, 3.59) | 1.19 (0.51, 2.76) |
| EV | — | — | 0.42 (0.06, 2.98) | 1.54 (0.70, 3.42) | 0.83 (0.33, 2.08) | 1.00 (0.58, 1.72) | — | — | 0.42 (0.06, 3.00) | 1.52 (0.68, 3.39) | 0.84 (0.33, 2.11) | 1.01 (0.58, 1.75) |
| RV | 0.43 (0.11, 1.73) | 0.70 (0.22, 2.19) | 1.20 (0.68, 2.11) | 1.03 (0.63, 1.69) | 1.46 (0.99, 2.15) | 1.26 (0.92, 1.71) | 0.43 (0.11, 1.75) | 0.70 (0.23, 2.21) | 1.21 (0.69, 2.12) | 1.03 (0.63, 1.69) | 1.45 (0.98, 2.14) | 1.25 (0.92, 1.71) |
| HMPV | 1.44 (0.20, 10.12) | — | 0.71 (0.10, 5.14) | **3.30 (1.47, 7.42)** | 0.84 (0.11, 6.27) | 0.56 (0.08, 3.92) | 1.46 (0.20, 10.45) | — | 0.72 (0.10, 5.18) | **3.41 (1.50, 7.74)** | 0.86 (0.11, 6.42) | 0.55 (0.08, 3.74) |
| HPIV | 1.44 (0.22, 9.51) | — | 2.04 (0.68, 6.12) | 0.67 (0.17, 2.69) | 1.85 (0.78, 4.37) | 0.93 (0.40, 2.19) | 1.47 (0.22, 9.70) | — | 2.04 (0.67, 6.17) | 0.67 (0.17, 2.68) | 1.90 (0.80, 4.54) | 0.95 (0.41, 2.21) |
| **Outcome: HMPV** | | | | | | | | | | | | |
| RSV | — | — | — | — | 0.92 (0.13, 6.74) | 2.28 (0.95, 5.44) | — | — | — | — | 0.77 (0.11, 5.57) | 1.93 (0.81, 4.57) |
| SC2 | 1.44 (0.21, 9.80) | — | 0.60 (0.08, 4.38) | 0.60 (0.15, 2.36) | 0.33 (0.06, 1.90) | 0.80 (0.33, 1.93) | 2.45 (0.61, 9.81) | — | 0.53 (0.07, 3.79) | 0.55 (0.14, 2.16) | 0.56 (0.14, 2.21) | 0.72 (0.30, 1.73) |
| FLU | — | — | — | — | 1.01 (0.14, 7.48) | 1.77 (0.57, 5.47) | — | — | — | — | 0.76 (0.10, 5.59) | 1.24 (0.39, 3.89) |
| ADV | — | — | — | — | 1.66 (0.19, 14.25) | **0.11 (0.03, 0.38)** | — | — | — | — | 1.65 (0.19, 14.28) | **0.13 (0.04, 0.43)** |
| EV | — | — | — | — | 1.19 (0.17, 8.39) | 1.27 (0.41, 3.89) | — | — | — | — | 1.14 (0.16, 8.08) | 1.27 (0.41, 3.88) |
| RV | 1.94 (0.28, 13.56) | — | 0.81 (0.20, 3.21) | 0.77 (0.26, 2.31) | 1.20 (0.41, 3.50) | 1.27 (0.68, 2.38) | 1.93 (0.27, 13.59) | — | 0.81 (0.20, 3.19) | 0.77 (0.26, 2.31) | 1.21 (0.42, 3.51) | 1.28 (0.68, 2.40) |
| HCOV | — | — | 0.48 (0.06, 3.57) | **2.17 (1.00, 4.71)** | 1.01 (0.27, 3.78) | 1.29 (0.31, 5.43) | — | — | 0.49 (0.07, 3.62) | **2.19 (1.01, 4.77)** | 1.03 (0.27, 3.88) | 1.28 (0.30, 5.35) |
| HPIV | — | — | 2.62 (0.34, 20.24) | 1.45 (0.22, 9.72) | 1.76 (0.24, 12.69) | 1.45 (0.35, 5.97) | — | — | 2.54 (0.33, 19.64) | 1.48 (0.22, 10.04) | 1.79 (0.25, 12.88) | 1.50 (0.37, 6.14) |
| **Outcome: HPIV** | | | | | | | | | | | | |
| RSV | — | — | 3.40 (0.81, 14.35) | 2.22 (0.53, 9.27) | 0.97 (0.15, 6.36) | 1.82 (0.73, 4.56) | 2.98 (0.42, 20.99) | — | 1.41 (0.19, 10.20) | 2.58 (0.77, 8.61) | 0.82 (0.13, 5.32) | 1.87 (0.80, 4.39) |
| SC2 | 1.82 (0.45, 7.44) | 2.05 (0.52, 8.11) | 1.08 (0.33, 3.55) | 0.83 (0.30, 2.25) | 0.91 (0.36, 2.30) | 0.80 (0.39, 1.66) | 1.50 (0.37, 6.12) | 0.82 (0.12, 5.78) | 1.26 (0.45, 3.51) | 0.72 (0.27, 1.93) | 0.79 (0.31, 2.00) | 0.70 (0.34, 1.43) |
| FLU | — | — | 2.63 (0.24, 28.53) | 0.32 (0.08, 1.39) | 3.14 (0.94, 10.52) | **2.56 (1.04, 6.33)** | — | — | 1.99 (0.19, 20.45) | 0.23 (0.05, 1.02) | 2.21 (0.64, 7.64) | 1.80 (0.72, 4.48) |
| ADV | 3.05 (0.40, 23.45) | — | 1.32 (0.21, 8.27) | 2.03 (0.55, 7.46) | 1.08 (0.19, 6.25) | 1.86 (0.69, 5.07) | 3.15 (0.41, 24.27) | — | 1.41 (0.23, 8.70) | 2.00 (0.54, 7.38) | 1.08 (0.18, 6.29) | 1.89 (0.69, 5.14) |
| EV | — | — | 0.67 (0.09, 4.79) | 1.09 (0.24, 4.88) | 2.40 (0.78, 7.39) | 0.61 (0.19, 1.98) | — | — | 0.67 (0.09, 4.83) | 1.09 (0.24, 4.91) | 2.43 (0.79, 7.47) | 0.60 (0.18, 1.97) |
| RV | — | — | 1.48 (0.71, 3.09) | 1.00 (0.48, 2.08) | 1.26 (0.64, 2.47) | 1.01 (0.59, 1.73) | — | — | 1.49 (0.71, 3.11) | 0.99 (0.47, 2.07) | 1.24 (0.63, 2.45) | 1.02 (0.60, 1.75) |
| HCOV | — | 1.33 (0.19, 9.38) | 2.03 (0.70, 5.86) | 1.31 (0.42, 4.06) | **2.29 (1.01, 5.18)** | 1.84 (0.90, 3.75) | — | 1.32 (0.19, 9.32) | 2.06 (0.71, 5.96) | 1.29 (0.42, 3.93) | **2.28 (1.01, 5.14)** | 1.82 (0.89, 3.73) |
| HMPV | — | — | — | — | — | 0.72 (0.11, 4.74) | — | — | — | — | — | 0.71 (0.11, 4.72) |

Columns within each block give exposure windows in days. Cell shading reflects the hazard ratio: navy below 1, white at 1, red above 1, clamped at 4. Bold indicates $p < 0.05$ ; em dashes denote cells where the hazard ratio was not estimable. Blank cells were not fitted in that model.

### Supplemental Figures

#### Supplemental Figure 1: Consort diagram

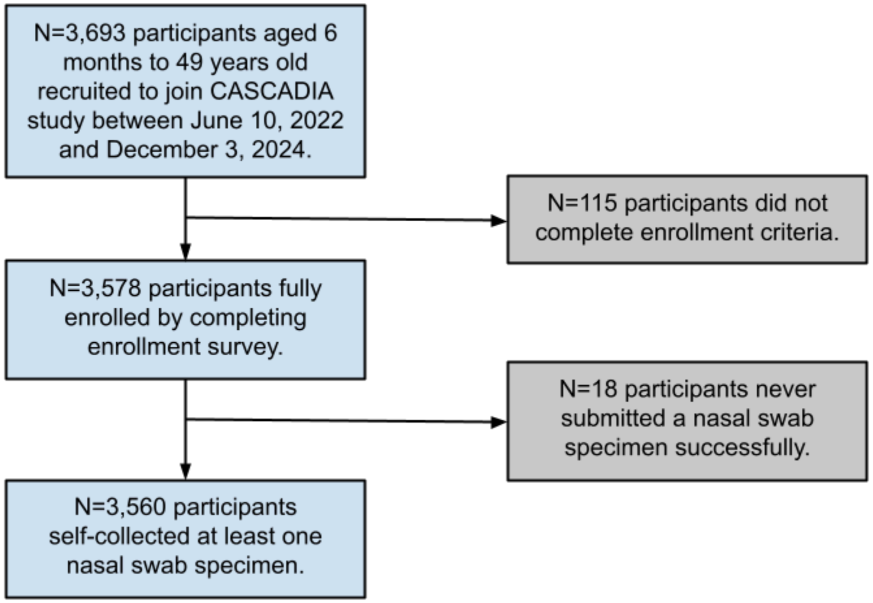

#### Supplemental Figure 2: Distribution of symptomatic swabs collected after each illness episode.

Distribution of the number of multipathogen (Open Array, OA) swabs associated with symptoms collected per participant in the 1-7, 8-14, and 15-30 days following each illness episode, for each virus category. The green dots show the distribution of the number of swabs after the initial positive swab of each illness episode. Each dot represents one illness episode, and the height of the dot corresponds to the number of swabs collected for that illness episode, as shown on the y-axis. The box plots show the median, 25th, and 75th percentile. The diamonds show the mean of each days and virus distribution.

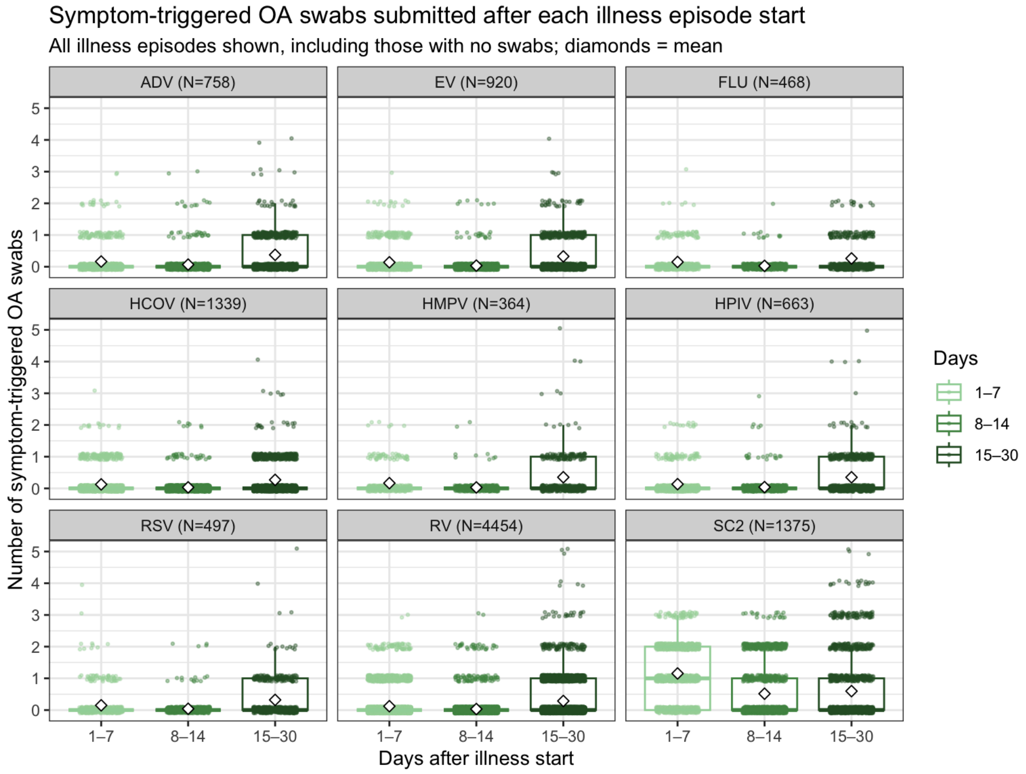

### Supplemental Figure 3: Sensitivity analyses with longer case-free windows.

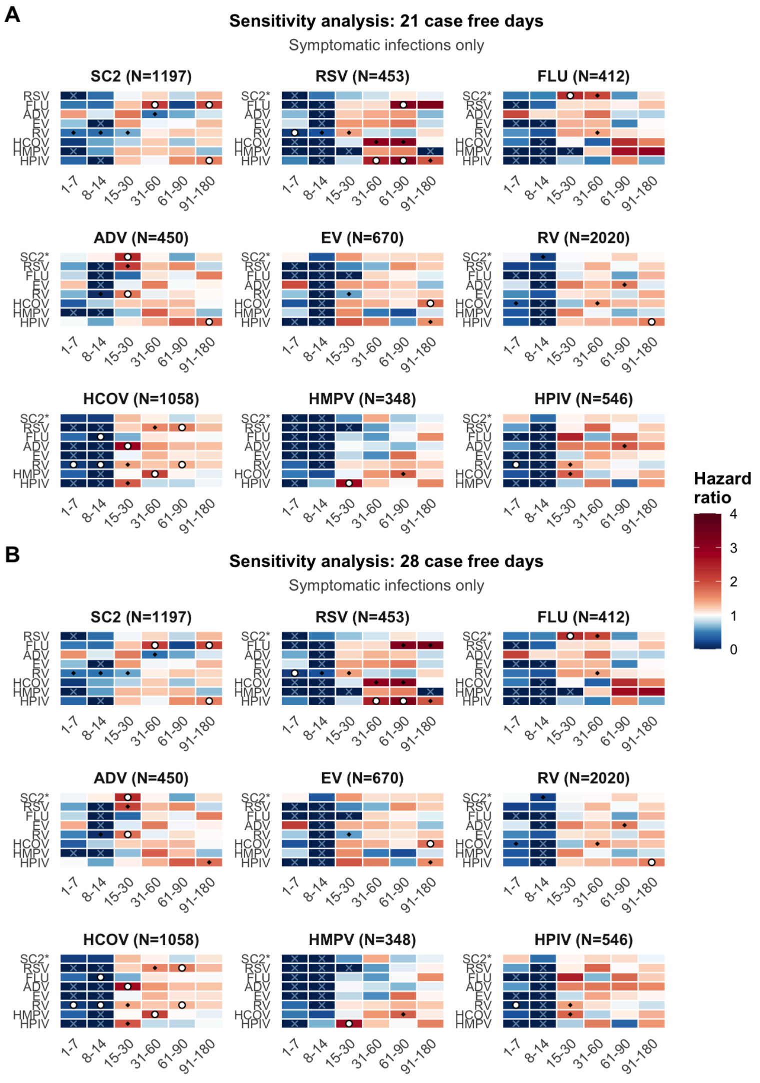


White circles denote a confidence interval excluding 1. Blue X marks denote complete separation of outcome and exposure data.

### Supplemental Figure 4: Sensitivity analyses with a 180 day washout period.

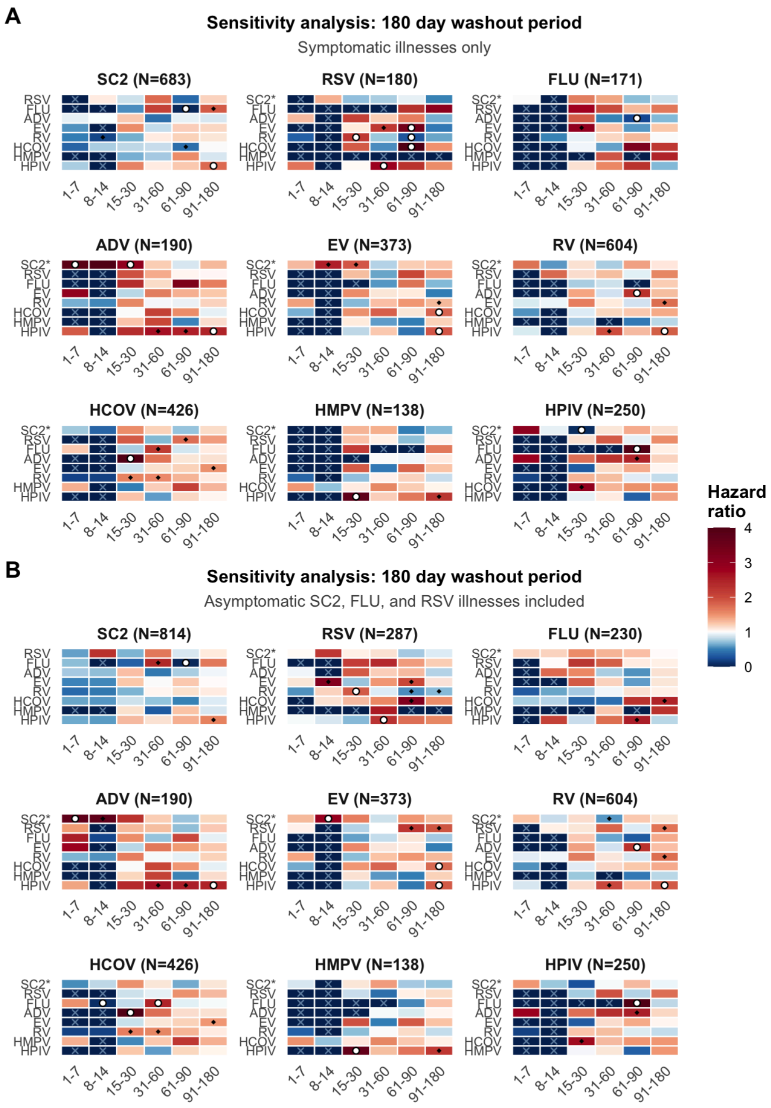


White circles denote a confidence interval excluding 1. Blue X marks denote complete separation of outcome and exposure data.

### Supplemental Methods

#### Laboratory methods

Swabs were stored dry, with no preservative or media (34286830), and nucleic acids were eluted with 300 µL Tris-EDTA from RHINOstic nasal swabs. Fifty µL of eluent was treated with proteinase K and heated at 37C for 15 min then 95C for 15 min 37C for 15 min then 95C for 15 min for direct RT-qPCR (Swab-Express RT-qPCR) as previously described (I.e. CRI) (PMCID 34286830) or 200 µL of eluent underwent total nucleic acid purification (Magna Pure 96 System, Roche Diagnostics, MagNA Pure 96 DNA and Viral NA Small Volume Kit; #06543588001) prior to RT-qPCR (ie CRIMP).

The RT-qPCR assay was arranged in two multiplexed reactions run in duplicate for each specimen. Both multiplexed reactions incorporate human RNase P as human tissue control. The pathogen targets were arranged as RSV A, Influenza B, and SARS-CoV-2 S-Gene in the first mix, and RSV B, Influenza A, and SARS- CoV-2 Orf1b in the second. The assay employs custom probe sets for the pathogens; RSV targets were identified by Gunson et al., Influenza by the CDC (https://www.cdc.gov/coronavirus/2019-ncov/downloads/lab/multiplex-primers-probes-printer.pdf), and SARS-CoV-2 Orf1b and S-gene designed against the ancestral strain (GenBank: MN908947.3) and are proprietary to Thermo Fisher. For SARS-CoV-2, positive samples had targets detected in three or four reactions. The 157-158 deletion in the Delta variants resulted in S-gene target failure or delay in our assay; thus, positive samples were identified by two Orf1b amplifications for this variant. Other pathogens required detection from both replicate reactions to be identified as positive. The internal RNase P control was detected in at least three of four reactions for the assay result to be considered valid.

Five µl of specimen was added to each well of a 384-well plate (Applied Biosystems, 4309849 city country) and each reaction was performed at a final volume of 10 µl. Plates were sealed with optically clear microseal B (Biorad) tape. RT-qPCR was performed on the Applied Biosystems QuantStudio 6 Pro (25°C for 2 minutes, 50°C for 15 minutes, 95°C for 2 minutes, followed by 40 cycles of 95°C for 3 seconds and 60°C for 30 seconds). Cycle threshold (Ct) values were obtained from the QuantStudio Design and Analysis software using predetermined ΔRn thresholds. The delta Rn value is the Rn value of an experimental reaction minus the Rn value of the baseline signal generated by the instrument. Positive controls contain nucleic acid sequences amplified by the Orf1b, S, Flu-A, Flu-B, RSV-A, and RSV-B assays and human RNA. A no-template control (low-TE blank) was included on each plate in a random position.

*Multipathogen testing*

Samples were selected for multipathogen (“open array”) testing if the swab was positive for SARS-CoV-2, influenza A, or RSV A on the multiplex assay, if the participant reported any new symptom within the following 72 hours, or if the participant reported any new symptom at the time the swab was taken. Multipathogen testing provides research-only results for detection of a broader array of pathogens including influenza A/H3N2, influenza A/H1N1, influenza B, parainfluenza, enterovirus, rhinovirus, adenovirus, human coronaviruses, SARS-CoV-2, RSV A/B, human metapneumovirus, and *Streptococcus pneumoniae.* Specimens selected for open array were processed as in prior studies (PMID 36150508, 36150508). Briefly, samples underwent nucleic acid purification and extracted nucleic acids underwent a pre-amplification RT-PCR prior to a re-array and qPCR byTaqMan real-time PCR (RT-PCR) on the OpenArray platform (Thermo) (PMID: 33033018).
